# Brain Age Prediction in Type II GM1 Gangliosidosis

**DOI:** 10.1101/2025.04.23.25326206

**Authors:** Connor J. Lewis, Selby I. Chipman, Precilla D’Souza, Jean M. Johnston, Muhammad H. Yousef, William A. Gahl, Cynthia J. Tifft, Maria T. Acosta

**Author notes:** Corresponding Author: Maria T. Acosta.

## Abstract

GM1 gangliosidosis is an inherited, progressive, and fatal neurodegenerative lysosomal storage disorder with no approved treatment. We calculated a predicted brain ages and Brain Structures Age Gap Estimation (BSAGE) for 81 MRI scans from 41 Type II GM1 gangliosidosis patients and 897 MRI scans from 556 neurotypical controls (NC) utilizing *BrainStructuresAges*, a machine learning MRI analysis pipeline. NC showed whole brain aging at a rate of 0.83 per chronological year compared with 1.57 in juvenile GM1 patients and 12.25 in late-infantile GM1 patients, accurately reflecting the clinical trajectories of the two disease subtypes. Accelerated and distinct brain aging was also observed throughout midbrain structures including the thalamus and caudate nucleus, hindbrain structures including the cerebellum and brainstem, and the ventricles in juvenile and late-infantile GM1 patients compared to NC. Predicted brain age and BSAGE both correlated with cross-sectional and longitudinal clinical assessments, indicating their importance as a surrogate neuroimaging outcome measures for clinical trials in GM1 gangliosidosis.

## Introduction

GM1 gangliosidosis is a fatal, ultra rare, neurodegenerative lysosomal storage disorder caused by biallelic mutations in GLB1 encoding β-galactosidase [1], which cleaves the terminal β-1,4 galactose of GM1 gangliosides, producing GM2 gangliosides for further catabolism [2]. Dysfunction of β-galactosidase causes the accumulation of toxic GM1 gangliosides primarily in the central nervous system (CNS), which contains an abundance of gangliosides [3].

GM1 gangliosidosis affects one in every 100,000 to 200,000 live births [4]. Three clinical sub-types of GM1 gangliosidosis exist, based on the age of symptom onset, which is inversely correlated with the amount of residual enzyme activity [5,6]. Type I (Infantile) GM1 gangliosidosis begins prior to the first year of life with symptoms including a macular cherry red spot, CNS dysfunction, and regression involving blindness, deafness, feeding difficulties, seizures, and oral secretions [4]. Type II GM1 gangliosidosis patients can be further divided into late-infantile (LI) and juvenile classifications based on the age of symptom onset [7]. LI GM1 patients have symptom onset at approximately 1 year of age; juvenile GM1 gangliosidosis patients have onset between the ages of 3 and 5 years old [7]. The adult or chronic form of GM1 gangliosidosis is the least severe with symptom onset typically in the second or third decade and characterized by prominent parkinsonism features [7,8].

Type II GM1 gangliosidosis patients typically present with gait disturbances, falling, and slurring of speech [9]. Ataxia, dystonia, dysarthria, skeletal changes, elevated liver function tests (especially aspartate aminotransferase), strabismus, decreased mobility, abnormal electroencephalography (EEG), and swallowing difficulties are all common in the Type II form of the disease [7]. LI GM1 gangliosidosis patients typically survive into the second decade; juvenile patients can survive into the fourth decade [7,9].

Numerous neuroimaging findings are associated with Type II GM1 gangliosidosis. Volumetric studies have shown decreased volumes of the whole brain, cerebellum, lentiform nucleus, thalamus, corpus callosum, and caudate nucleus, with consequent enlargement of the lateral ventricles [10–13]. T1-weighted signal abnormalities of the bilateral globus pallidum [14], increased signal attenuation in the thalamus, and decreased signal attenuation of the basal ganglia have been documented, as well as decreased T2-weighted signal intensity in the thalamus, globus pallidus, and substantia nigra and hyperintensities of the putamen [14–17]. Magnetic resonance spectroscopy (MRS) shows diminished N-acetyl-aspartate (NAA) in the thalamus and centrum semi-ovale and increased myo-inositol in the centrum semi-ovale [7,17].

There are no treatments for GM1 gangliosidosis [18], although gene therapy clinical trials are currently underway for GM1 [19]. As these trials move to the latter phases, outcome measures demonstrating the efficacy of the therapeutic intervention will be required. Due to the CNS pathology of GM1, analysis of anatomical magnetic resonance imaging could serve as an objective outcome measure [20,21]. In this study, we present the first use of MRI-based predicted brain age in GM1 gangliosidosis aimed at assessing the neuronal degeneration in this cohort. These findings could serve as imaging outcome parameters for demonstrating therapeutic efficacy attendant to interventions such as gene therapy.

## 2 Methods

### 2.1 Type 2 GM1 Gangliosidosis Participants

Forty-one GM1 patients from the “Natural History of Glycosphingolipid Storage Disorders and Glycoprotein Disorders” protocol (ClinicalTrials.gov ID: NCT00029965) were included in this study [22]. As described in D’Souza et al [7], a GM1 diagnosis was based upon β-galactosidase enzyme deficiency or biallelic pathogenic variants in GLB1. Participants were classified into juvenile or LI forms of GM1 based on the age of symptom onset. 61 MRI scans from 26 juvenile (baseline age: 9.6 ± 4.7 years) patients were included in this study alongside 20 MRI scans from 15 late-infantile patients (baseline scan age: 4.6 ± 1.8 years, Section A of the Supplementary Materials).

### 2.2 Neurotypical Control Participants

To account for the nearly 25-year age range of GM1 gangliosidosis patients. Neurotypical control MRI scans were gathered from three open-source data sets. First, 97 participants (279 scans) from the Calgary Preschool MRI dataset were included consisting of participants between the ages of 2 and 8 years (baseline scan age: 3.84 ± 0.88 years old) [23,24]. Second, 159 participants (318 scans) from the Queensland Twin Adolescent Brain (QTAB) were included in this study, consisting of participants between the ages of 9 and 16 years (baseline scan age: 10.93 ± 1.36 years old) [25,26]. Lastly, 300 participants (300 scans) from the Queensland Twin Imaging (QTIM) were included in this study, consisting of participants between the ages of 18 and 30 years (baseline scan age: 21.72 ± 3.71 years old) [27,28].

### 2.3 T1-Weighted MRI Acquisition – GM1 Patients

All GM1 patients’ MRI scans were performed under propofol sedation and were performed on a Philips 3T system (Achieva, Philips Healthcare, Best, The Netherlands) with an 8-channel SENSE head coil. Images were acquired using a 3D T1-weighted protocol with the following parameters: 1 mm isotropic resolution, TR of 11 ms, TE of 7 ms, flip angle = 6 degrees, FOV = 220 mm [11]. Unprocessed digital imaging and communications in medicine (DICOM) images were converted to NIfTI using *dcm2niix* [29].

### 2.4 T1-Weighted MRI Acquisition – Calgary Preschool MRI

As described in Reynolds et al. [23,24], Calgary preschool neurotypical controls were scanned without sedation on a General Electric 3T system (MR750w, GE Healthcare, Chicago, IL, USA) using a 32-channel head coil. T1-weighted imaging from preschool neurotypical controls was acquired with TR/TE = 8.23/3.76 ms, flip angle = 12 degress, and resolution of 0.9 mm × 0.9 mm × 0.9 mm (resampled to 0.45 mm × 0.45 mm × 0.9 mm), and 210 slices.

### 2.5 T1-Weighted MRI Acquisition – Queensland Twin Adolescent Brain (QTAB) MRI

As described in Strike et al. [25,26], participants from QTAB were scanned on a Siemens 3T Prisma system with a 64-channel head coil at the Centre for Advanced Imaging at the University of Queensland, Australia. Images were acquired using a 3D T1-weighted Magnetization Prepared with 2 Rapid Gradient Echoes (MP2RAGE) imaging protocol with the following parameters: 0.8 mm isotropic voxels, TR/TE = 4000/2.99 ms, flip angle of 6 or 7 degrees, FOV = 256 × 240 mm, and 192 slices.

### 2.6 T1-Weighted MRI Acquisition – Queensland Twin IMaging (QTIM) MRI

As described in Strike et al. [27,28], Participants from QTIM were scanned on a 4T 4T Bruker Medspec (Bruker, Germany) with a transverse electromagnetic (TEM) head coil. Images were acquired using a 3D T1-weighted protocol with the following parameters: 0.9 mm isotropic voxels, TR/TE=1500/3.35 ms, FOV = 230 mm, and 256 or 240 slices.

### 2.7 MRI Processing

All MRI scans from GM1 patients and NC were sent through volBrain’s *BrainStructuresAges* pipeline [30–32]. There were no excluded MRI scans or scans which failed to process. *BrainStructuresAges* is a deep learning MRI analysis pipeline trained with 2887 MRI scans from various sources to predict brain age [30]. The pipeline was then shown to be more accurate in terms of mean absolute error and R^2^ than other state-of-the-art models in analyzing more than 30,000 MRI scans. We used *BrainStructuresAges* to predict ages for the whole brain, white matter, 3^rd^ ventricle, 4^th^ ventricle, lateral ventricles, inferior lateral ventricles, cerebellar gray matter, cerebellar white matter, lobules I-V of the cerebellum, lobules VI and VII of the cerebellum, lobules VIII-X of the cerebellum, the caudate nucleus, thalamus, brainstem, hippocampus, nucleus accumbens, amygdala, ventral diencephalon (DC), external cerebrospinal fluid (CSF), basal forebrain, putamen, and globus pallidus in all participants. Brain structures age gap estimation (BSAGE) was calculated as the predicted brain age minus the patient’s chronological age as described in Nguyen et al. [30].

### 2.8 Clinical Global Impressions (CGI) Scores

To determine the clinical relevance of predicted brain age in GM1 gangliosidosis participants, we correlated these metrics with clinical global impression (CGI) scores. CGI scores a clinician rated outcome measure assessing the clinical severity of a patient [33,34]. Clinical Global Impressions were scored by consensus after a retrospective chart review as described in Lewis et al. [35], baseline CGI Severity (CGI-S) scores were assigned 1 = ‘normal’ to 7 = “among the most extremely” at the patients’ baseline evaluation.

### 2.9 Vineland Adaptive Behavior Composite (ABC)

The Vineland Adaptive Behavioral Composite (ABC) Standard Scores are a semi-structured clinical interview with the patient and their caregiver [36,37]. The ABC domain of the Vineland is functional impairment measure with a standard score that has a population mean of 100 and standard deviation of 15. Vineland-3 scores were used when available, and Vineland-II scores were used when the third edition was not available (neurodevelopmental assessment was conducted before the 3^rd^ edition’s release) [35]. As described in D’Souza et al. [7], GM1 gangliosidosis patients demonstrated functional impairments assessed by the Vineland scale including the ABC and fine motor, gross motor, socialization, daily living, and communication subdomains in both LI and juvenile GM1 patients.

### 2.10 Statistical Analysis

Linear mixed effects modeling (LMEM) in this study was performed in R (The R Foundation, v4.3.1), with the lme4 package [38]. The effect of participant sex (fixed effect) was modeled as a covariate of no interest and a subject level random intercept was used to account for repeated T1 scans conducted for each participant [39,40]. Males were assigned a value of 1 and females were assigned a value of 0 to remove the effect of sex from LMEM. Between group effects were assessed as the interaction between participants’ chronological age and their group. For comparisons between GM1 gangliosidosis patients and neurotypical controls, NC were assigned a value of 0 and GM1 gangliosidosis patients were assigned a value of 1 to test the effects of GM1 gangliosidosis on the age predictions. For comparisons between late-infantile (LI) GM1 patients and neurotypical controls, NC were assigned a value of 0 and LI GM1 patients were assigned a value of 1 to test the effects of the late-infantile disease on the age predictions. For comparisons between juvenile GM1 patients and neurotypical controls, NC were assigned a value of 0 and Juv GM1 patients were assigned a value of 1 to test the effects of the juvenile disease on the age predictions. For comparisons between Juv GM1 patients and LI GM1 patients, Juv patients were assigned a value of 0 and LI GM1 patients were assigned a value of 1 to test the effects of the disease subtype on the age predictions. The interaction between participant age and the cohort comparison variable was evaluated using a likelihood-ratio test as shown in section C of the supplementary materials. P-values < 0.05 were considered significant after a Bonferri correction. An ordinary one-way analysis of variation (ANOVA) was performed using Graphpad Prism to compare for differences in BSAGE between groups with a post-hoc Tukey test (version 10.1.0, GraphPad Software, Boston, MA). Spearman Correlations were performed between predicted brain age and clinical metrics and were also performed in GraphPad Prism.

## 3. Results

### 3.1 GM1 vs NC

In the comparisons between NC and the entire GM1 cohort, neurotypical controls showed an average increase in predicted whole brain age of 0.83 years per one year increase in biological age with no statistical effect of sex (𝜒^2^(1) = 1.76, *p* = 0.19). Predicted whole brain age was affected in two ways in GM1 gangliosidosis patients (Figure 1). First, there was a constant effect, with GM1 patients having a mean 18.57 ± 1.73-year higher predicted whole brain age compared to controls (Supplement Table G1). Second, the interaction between biological age and GM1 showed further divergence with an average increase in predicted whole brain age of 0.74 ± 0.14 per year, resulting in an average predicted whole brain increase of 1.57 years per one year increase in biological age compared to 0.83 in the neurotypical control cohort (𝜒^2^(1) = 27.78, *p* < 0.0001, Table 1, Supplement Figure F1). Figure 2 illustrates brain aging in GM1 gangliosidosis patients; at age 10 the GM1 patients demonstrate significant aging outside of the scalar range.

**Figure 1.**
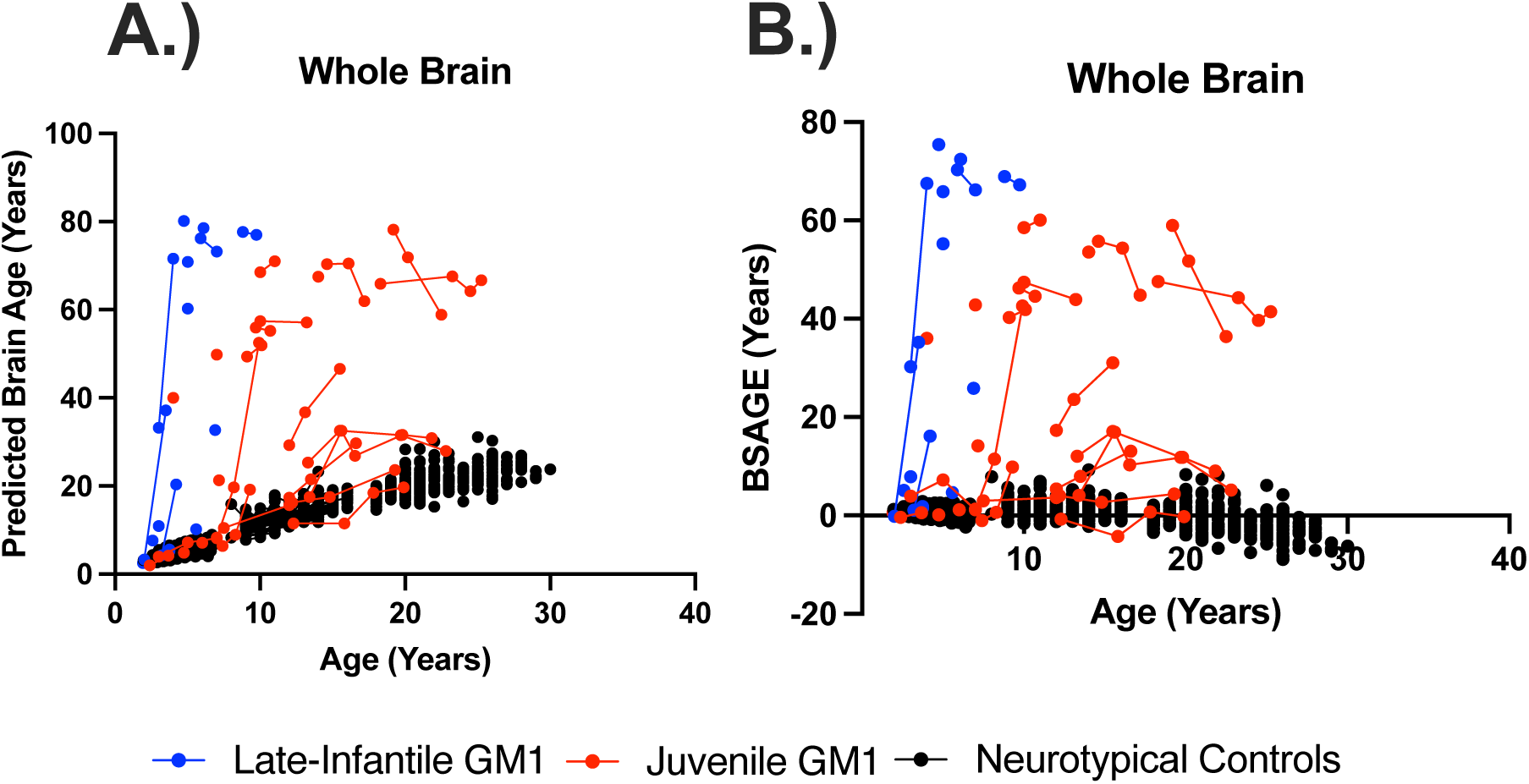
The longitudinal relationship between (A) predicted whole brain age and (B) whole brain, brain structures age gap estimation (BSAGE) with chronological age. Late-infantile (n = 15, 20 total MRI scans) GM1 gangliosidosis patients are shown in blue, juvenile (n = 26, 61 total MRI scans) GM1 gangliosidosis patients are shown in red, and neurotypical controls (n = 556, 897 total MRI scans) participants are shown in black. Connecting lines indicate repeated scans on the same participant. Statistical analysis was performed using a linear mixed effects model to test the interactions between chronological age and cohort on predicted brain age. The whole GM1 cohort (𝜒^2^(1) = 27.78, *p* < 0.0001), juvenile GM1 patients (𝜒^2^(1) = 67.32, *p* < 0.0001), and late-infantile GM1 patients (𝜒^2^(1) = 383.80, *p* < 0.0001) were found to have a statistically significant increasing effect on predicted whole brain age compared to neurotypical controls. Neurotypical controls showed a rate of brain aging at 0.83 years per chronological year compared to 1.57 years per chronological year in the GM1 cohort, 1.79 years per chronological year in the juvenile GM1 cohort, and 12.20 years per chronological year in the late-infantile cohort. There was also a significant interaction effect of the late-infantile disease (𝜒^2^(1) = 15.91, *p* < 0.0001) compared to the juvenile disease where juvenile patients showed a rate of whole brain aging of 1.65 years per chronological year compared to 12.35 years per chronological year in late-infantile patients. The estimates and standard errors for the linear mixed effects modeling can be found in Supplementary Table G1. Longitudinal relationships between BSAGE and chronological age can be found in Section I of the Supplementary Materials.

**Figure 2.**
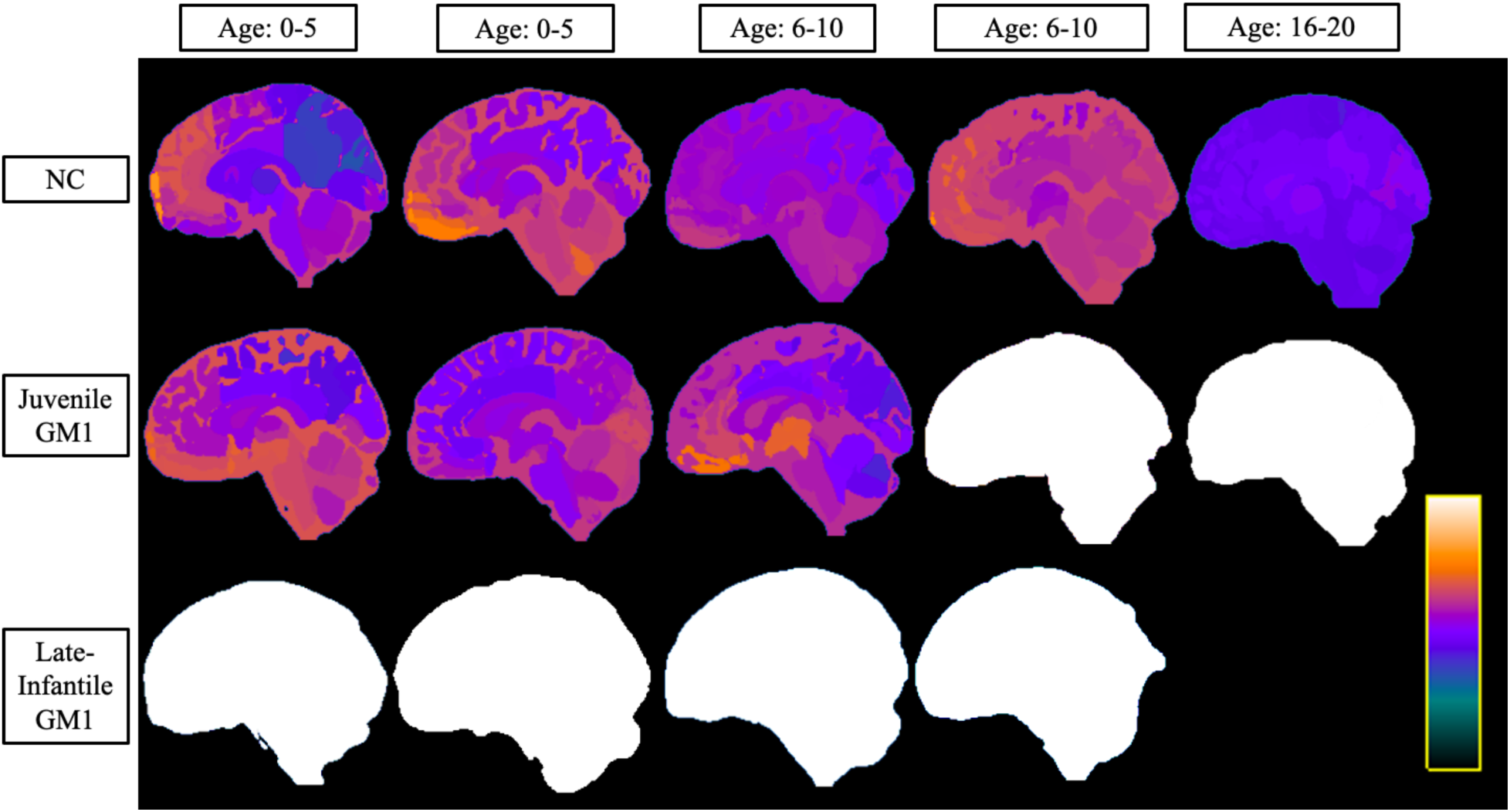
Sagittal View of Brain Structures Ages of the Three Cohorts at Varying Ages. Neurotypical controls (NC) are shown on the top row, juvenile GM1 gangliosidosis patients are shown in the middle row, and late-infantile GM1 gangliosidosis patients are shown in the bottom row. Colors were assigned according to the predicted age of that brain structure and were assigned between very dark blue (lowest predicted age) and white (highest predicted age) as shown in the scalar bar in the bottom right portion. The scalar ranges were adjusted for each column with min/max values of 0-8 years for the first column (0-5 years old), 0-10 years for the second column (0-5 years old), 0-14 years for the third column (6-10 years old), 0-20 years for the fourth column (6-10 years old), and 0-40 years for the fifth column (16-20 years old). No 20-year-old MRI scan was available for late-infantile patients as the oldest late-infantile patient in this study was 6-10 years old at the time of the MRI evaluation. All MRI images were registered to the MNI coordinate space and slices were taken at x = 1.0 (± 1) mm for each participant. A coronal, axial, and a matching anatomical version of this image is available in section D of the Supplementary Material. Specific ages were redacted per Medrxiv requirements.

**Table 1.**
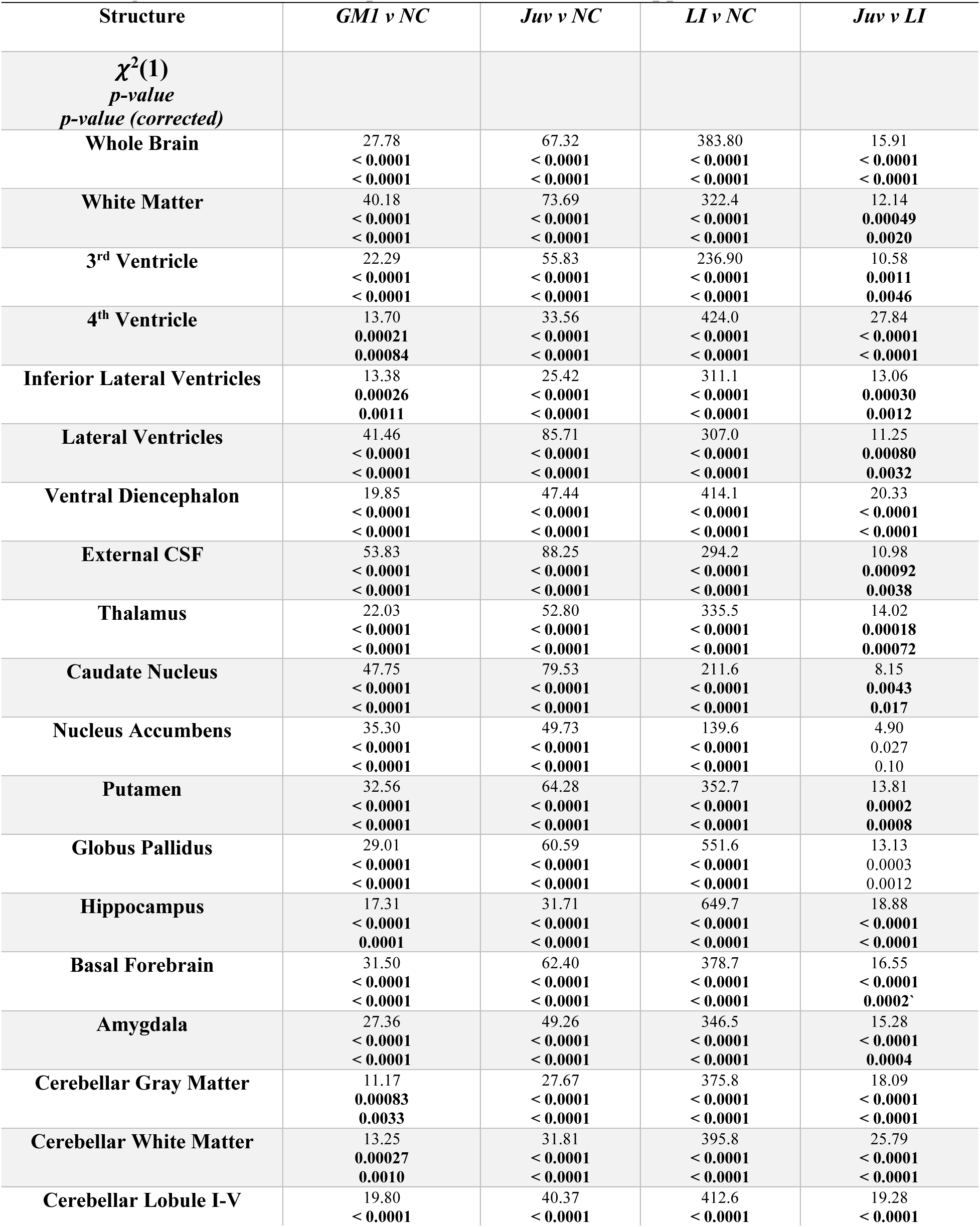
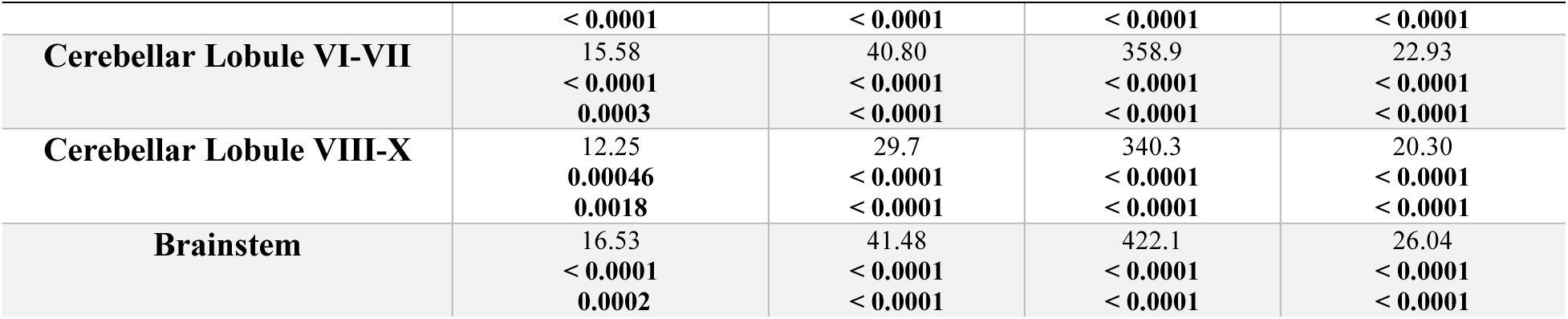
Brain Age Predictions Results from Linear Mixed Effects Modeling. Estimates and Standard Errors from the LMEM for the interaction between participant age and the cohort comparison can be found in Supplement Table G1.

| Structure | *GMI v NC* | *Juv v NC* | *LI v NC* | *Juv v LI* |
| --- | --- | --- | --- | --- |
| $\chi^2(1)$<br>*p-value*<br>*p-value (corrected)* | | | | |
| **Whole Brain** | 27.78<br>< 0.0001<br>< 0.0001 | 67.32<br>< 0.0001<br>< 0.0001 | 383.80<br>< 0.0001<br>< 0.0001 | 15.91<br>< 0.0001<br>< 0.0001 |
| **White Matter** | 40.18<br>< 0.0001<br>< 0.0001 | 73.69<br>< 0.0001<br>< 0.0001 | 322.4<br>< 0.0001<br>< 0.0001 | 12.14<br>0.00049<br>0.0020 |
| **3<sup>rd</sup> Ventricle** | 22.29<br>< 0.0001<br>< 0.0001 | 55.83<br>< 0.0001<br>< 0.0001 | 236.90<br>< 0.0001<br>< 0.0001 | 10.58<br>0.0011<br>0.0046 |
| **4<sup>th</sup> Ventricle** | 13.70<br>0.00021<br>0.00084 | 33.56<br>< 0.0001<br>< 0.0001 | 424.0<br>< 0.0001<br>< 0.0001 | 27.84<br>< 0.0001<br>< 0.0001 |
| **Inferior Lateral Ventricles** | 13.38<br>0.00026<br>0.0011 | 25.42<br>< 0.0001<br>< 0.0001 | 311.1<br>< 0.0001<br>< 0.0001 | 13.06<br>0.00030<br>0.0012 |
| **Lateral Ventricles** | 41.46<br>< 0.0001<br>< 0.0001 | 85.71<br>< 0.0001<br>< 0.0001 | 307.0<br>< 0.0001<br>< 0.0001 | 11.25<br>0.00080<br>0.0032 |
| **Ventral Diencephalon** | 19.85<br>< 0.0001<br>< 0.0001 | 47.44<br>< 0.0001<br>< 0.0001 | 414.1<br>< 0.0001<br>< 0.0001 | 20.33<br>< 0.0001<br>< 0.0001 |
| **External CSF** | 53.83<br>< 0.0001<br>< 0.0001 | 88.25<br>< 0.0001<br>< 0.0001 | 294.2<br>< 0.0001<br>< 0.0001 | 10.98<br>0.00092<br>0.0038 |
| **Thalamus** | 22.03<br>< 0.0001<br>< 0.0001 | 52.80<br>< 0.0001<br>< 0.0001 | 335.5<br>< 0.0001<br>< 0.0001 | 14.02<br>0.00018<br>0.00072 |
| **Caudate Nucleus** | 47.75<br>< 0.0001<br>< 0.0001 | 79.53<br>< 0.0001<br>< 0.0001 | 211.6<br>< 0.0001<br>< 0.0001 | 8.15<br>0.0043<br>0.017 |
| **Nucleus Accumbens** | 35.30<br>< 0.0001<br>< 0.0001 | 49.73<br>< 0.0001<br>< 0.0001 | 139.6<br>< 0.0001<br>< 0.0001 | 4.90<br>0.027<br>0.10 |
| **Putamen** | 32.56<br>< 0.0001<br>< 0.0001 | 64.28<br>< 0.0001<br>< 0.0001 | 352.7<br>< 0.0001<br>< 0.0001 | 13.81<br>0.0002<br>0.0008 |
| **Globus Pallidus** | 29.01<br>< 0.0001<br>< 0.0001 | 60.59<br>< 0.0001<br>< 0.0001 | 551.6<br>< 0.0001<br>< 0.0001 | 13.13<br>0.0003<br>0.0012 |
| **Hippocampus** | 17.31<br>< 0.0001<br>0.0001 | 31.71<br>< 0.0001<br>< 0.0001 | 649.7<br>< 0.0001<br>< 0.0001 | 18.88<br>< 0.0001<br>< 0.0001 |
| **Basal Forebrain** | 31.50<br>< 0.0001<br>< 0.0001 | 62.40<br>< 0.0001<br>< 0.0001 | 378.7<br>< 0.0001<br>< 0.0001 | 16.55<br>< 0.0001<br>0.0002 |
| **Amygdala** | 27.36<br>< 0.0001<br>< 0.0001 | 49.26<br>< 0.0001<br>< 0.0001 | 346.5<br>< 0.0001<br>< 0.0001 | 15.28<br>< 0.0001<br>0.0004 |
| **Cerebellar Gray Matter** | 11.17<br>0.00083<br>0.0033 | 27.67<br>< 0.0001<br>< 0.0001 | 375.8<br>< 0.0001<br>< 0.0001 | 18.09<br>< 0.0001<br>< 0.0001 |
| **Cerebellar White Matter** | 13.25<br>0.00027<br>0.0010 | 31.81<br>< 0.0001<br>< 0.0001 | 395.8<br>< 0.0001<br>< 0.0001 | 25.79<br>< 0.0001<br>< 0.0001 |
| **Cerebellar Lobule I-V** | 19.80<br>< 0.0001 | 40.37<br>< 0.0001 | 412.6<br>< 0.0001 | 19.28<br>< 0.0001 |
|  | < 0.0001 | < 0.0001 | < 0.0001 | < 0.0001 |
| **Cerebellar Lobule VI-VII** | 15.58 | 40.80 | 358.9 | 22.93 |
|  | < 0.0001 | < 0.0001 | < 0.0001 | < 0.0001 |
|  | 0.0003 | < 0.0001 | < 0.0001 | < 0.0001 |
| **Cerebellar Lobule VIII-X** | 12.25 | 29.7 | 340.3 | 20.30 |
|  | 0.00046 | < 0.0001 | < 0.0001 | < 0.0001 |
|  | 0.0018 | < 0.0001 | < 0.0001 | < 0.0001 |
| **Brainstem** | 16.53 | 41.48 | 422.1 | 26.04 |
|  | < 0.0001 | < 0.0001 | < 0.0001 | < 0.0001 |
|  | 0.0002 | < 0.0001 | < 0.0001 | < 0.0001 |

Similar differences between NC and GM1 patients in predicted whole brain age were observed for the predicted ages of the white matter, 3^rd^ and 4^th^ ventricles, inferior lateral ventricles, lateral ventricles (Figure 3), external CSF, ventral diencephalon, thalamus, caudate nucleus, nucleus accumbens, putamen (Figure 4), globus pallidus, hippocampus, basal forebrain, amygdala, cerebellar gray and white matter (Figure 5), lobules I-V of the cerebellum, lobules VI and VII of the cerebellum, lobules VIII-X of the cerebellum, and the brainstem, as summarized in Table 1.

**Figure 3.**
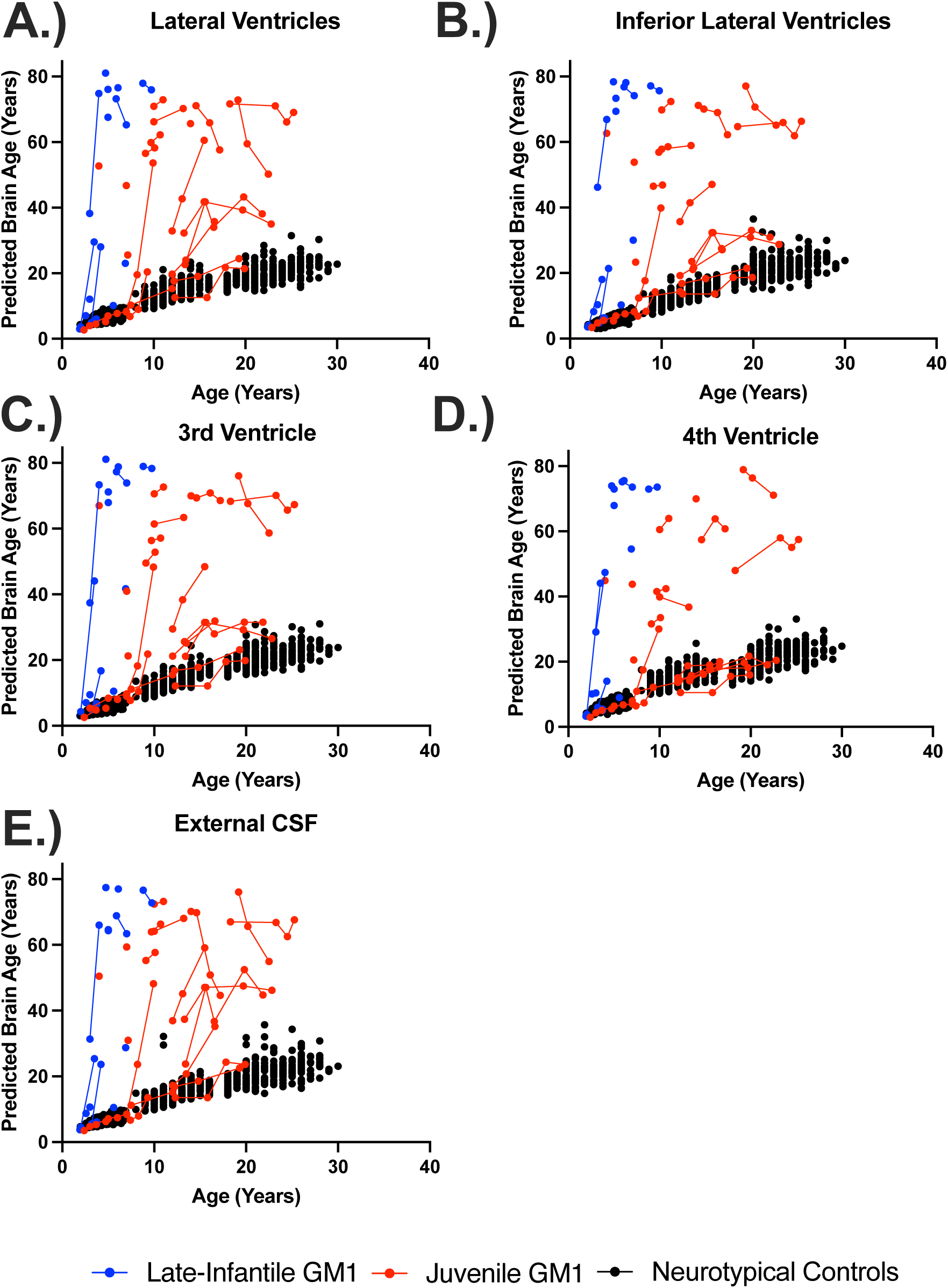
The relationship between predicted brain age and biological age in ventricle and cerebrospinal structures. Predicted brain ages for the (A) lateral ventricles, (B) inferior lateral ventricles, (C) 3^rd^ ventricles, (D) 4^th^ ventricle, and (E) external cerebrospinal fluid (CSF) are plotted against chronological age. Late-infantile (n = 15, 20 total MRI scans) GM1 gangliosidosis patients are shown in blue, juvenile (n = 26, 61 total MRI scans) GM1 gangliosidosis patients are shown in red, and neurotypical controls (n = 556, 897 total MRI scans) participants are shown in black. Connecting lines indicate repeated scans on the same participant. Statistical analysis was performed using a linear mixed effects model to test the interactions between chronological age and cohort on predicted brain age and the results are summarized in Table I, with the estimated and standard errors shown in Table G1 of the Supplementary Materials.

**Figure 4.**
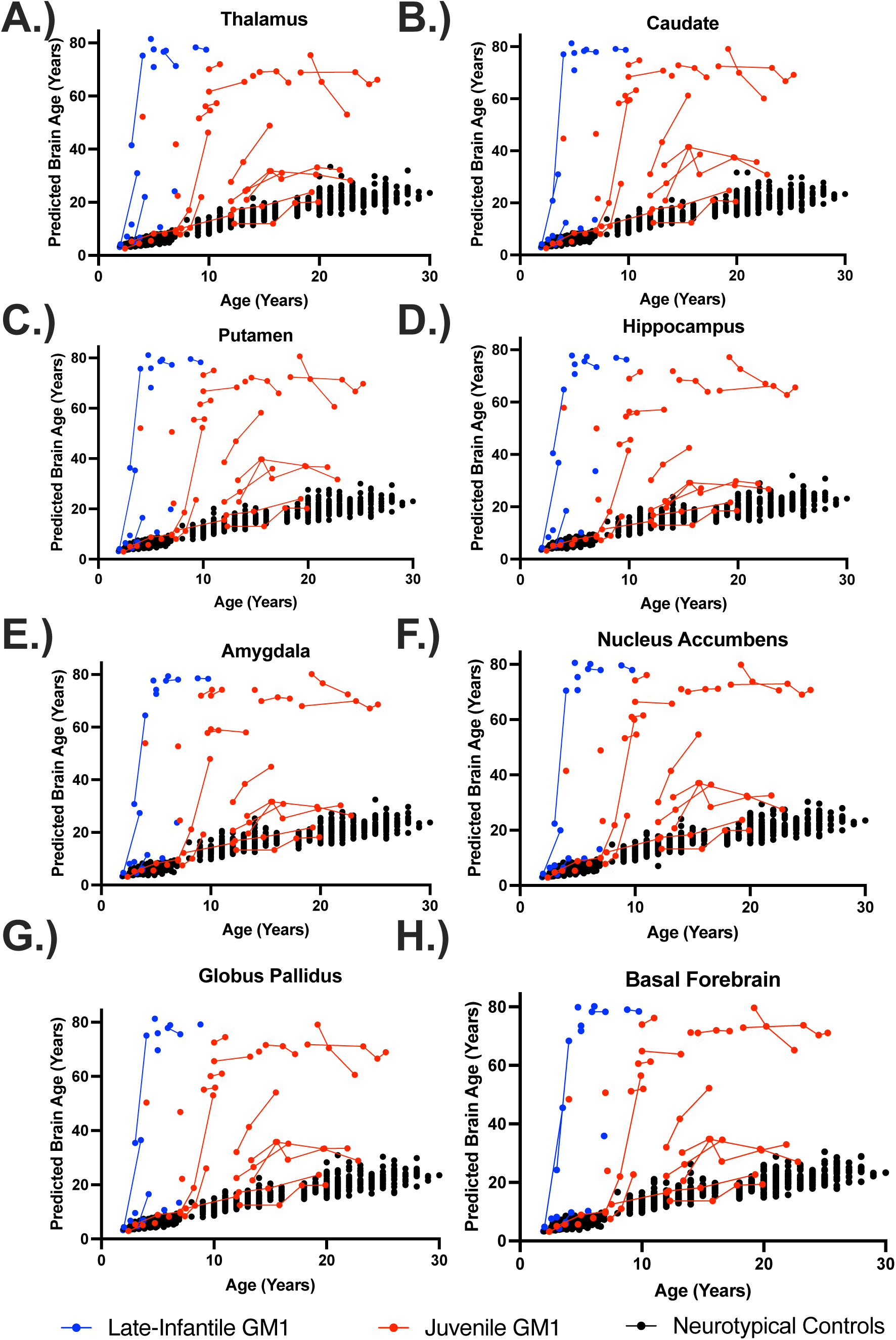
The relationship between predicted brain age and biological age in midbrain structures. Predicted brain ages for the (A) thalamus, (B) caudate nucleus, (C) putamen, (D) hippocampus, (E) amygdala, (F) nucleus accumbens, (G) globus pallidus, and (H) the basal forebrain are plotted against chronological age. Late-infantile (n = 15, 20 total MRI scans) GM1 gangliosidosis patients are shown in blue, juvenile (n = 26, 61 total MRI scans) GM1 gangliosidosis patients are shown in red, and neurotypical controls (n = 556, 897 total MRI scans) participants are shown in black. Connecting lines indicate repeated scans on the same participant. Statistical analysis was performed using a linear mixed effects model to test the interactions between chronological age and cohort on predicted brain age and the results are summarized in Table I, with the estimated and standard errors shown in Table G1 of the Supplementary Materials.

**Figure 5.**
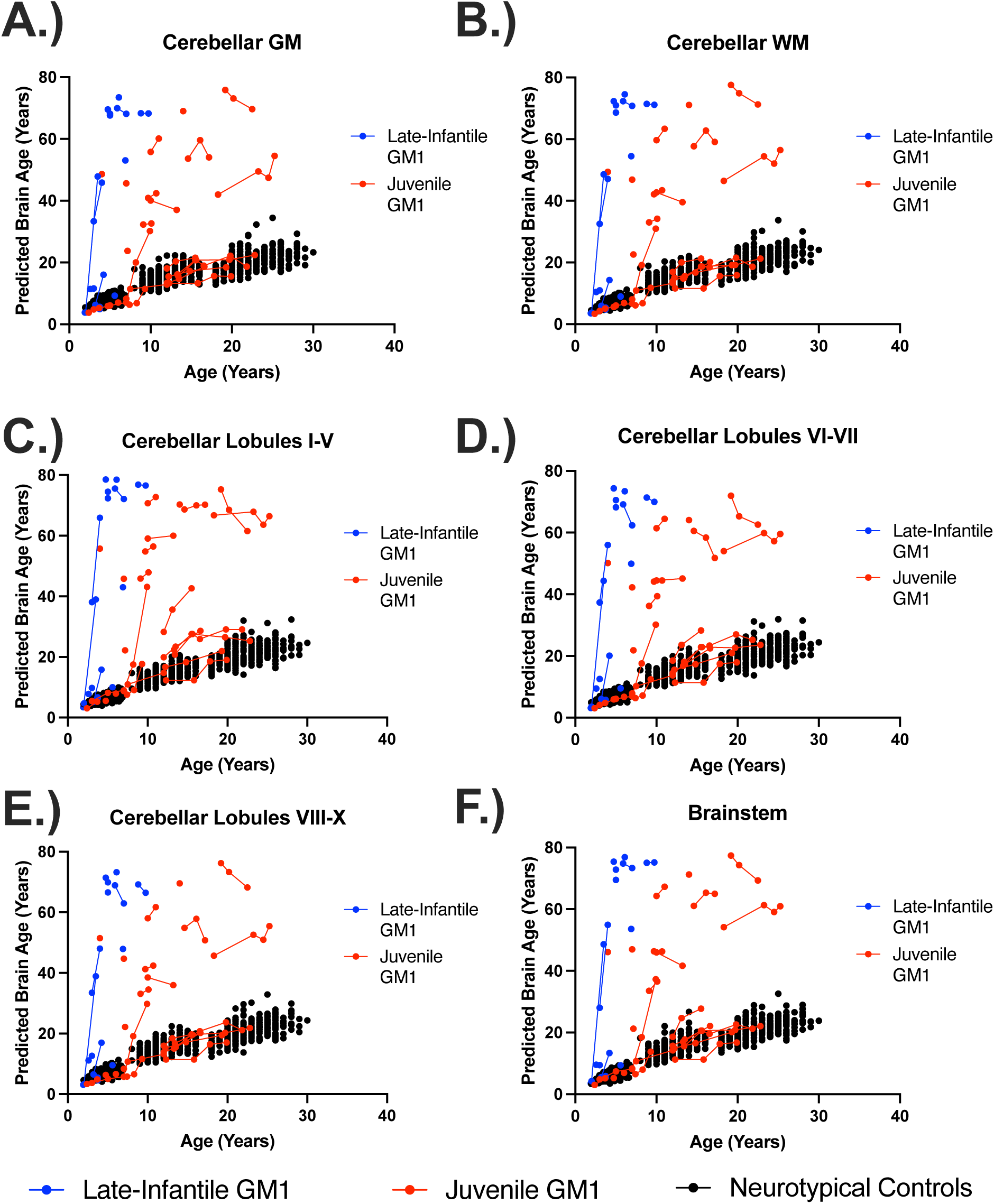
The relationship between predicted brain age and biological age in hindbrain structures. Predicted brain ages for the (A) cerebellar gray matter (GM), (B) cerebellar white matter (WM), (C) cerebellar lobules I-V, (D) cerebellar lobules VI-VII, (E) cerebellar lobules VIII-X, and (F) the brainstem are plotted against chronological age. Late-infantile (n = 15, 20 total MRI scans) GM1 gangliosidosis patients are shown in blue, juvenile (n = 26, 61 total MRI scans) GM1 gangliosidosis patients are shown in red, and neurotypical controls (n = 556, 897 total MRI scans) participants are shown in black. Connecting lines indicate repeated scans on the same participant. Statistical analysis was performed using a linear mixed effects model to test the interactions between chronological age and cohort on predicted brain age and the results are summarized in Table I, with the estimated and standard errors shown in Table G1 of the Supplementary Materials.

### 3.2 Juv GM1 vs NC

In the comparisons between NC and the juvenile GM1 cohorts, predicted whole brain age was also affected in two ways. First, there was a constant effect by which juvenile GM1 patients had a 10.31 ± 1.61-year higher predicted whole brain age on average compared to controls (Supplement Table G1). Second, the interaction between biological age and juvenile GM1 showed further divergence with an average increase in predicted whole brain age of 0.96 ± 0.11 per year resulting in an average predicted whole brain increase of 1.79 years per one year increase in biological age compared to 0.83 in the neurotypical control cohort (𝜒^2^(1) = 67.32, *p* < 0.0001, Table 1, Supplement Figure F2). Figure 1 also illustrates brain aging in the juvenile GM1 gangliosidosis patients; at the younger ages (3-7 years old), the juvenile patients appeared similar to the NC, but at the older ages (10 years old and above) the juvenile patients showed significant aging outside of the scalar range.

Similar relationships between NC and juvenile GM1 patients in predicted whole brain age were observed for the predicted ages of the white matter, 3^rd^ and 4^th^ ventricles, inferior lateral ventricles, lateral ventricles (Figure 3), external CSF, ventral diencephalon, thalamus, caudate nucleus (Figure 4), nucleus accumbens, putamen, globus pallidus, hippocampus, basal forebrain, amygdala, cerebellar gray and white matter, lobules I-V of the cerebellum, lobules VI and VII of the cerebellum (Figure 5), lobules VIII-X of the cerebellum, and the brainstem, as summarized in Table 1.

### 3.3 LI GM1 vs NC

In the comparisons between NC and LI GM1 cohorts, there was a constant effect whereby LI GM1 patients had a −21.29 ± 2.76-year lower predicted brain age on average compared to controls. The interaction between biological age and late-infantile GM1 showed substantial divergence with an average increase in predicted whole brain age of 11.37 ± 0.52 per year resulting in an average predicted whole brain increase of 12.20 per one year increase in biological age compared to 0.83 in the neurotypical control cohort (𝜒^2^(1) = 383.30, *p* < 0.0001, Table 1, Supplement Figure F3). Figure 1 also illustrates brain aging in the late-infantile GM1 gangliosidosis patients; at all ages the late-infantile patient show significant aging outside of the scalar range. Similar relationships between NC and LI GM1 patients in predicted whole brain age were observed for the predicted ages of the white matter, 3^rd^ and 4^th^ ventricles, inferior lateral ventricles, lateral ventricles, external CSF, ventral diencephalon, thalamus (Figure 3), caudate nucleus, nucleus accumbens, putamen, globus pallidus, hippocampus (Figure 4), basal forebrain, amygdala, cerebellar gray and white matter, lobules I-V of the cerebellum, lobules VI and VII of the cerebellum, lobules VIII-X of the cerebellum (Figure 5), and the brainstem, as summarized in Table 1.

### 3.4 Juv GM1 vs LI GM1

In the comparisons between Juv GM1 and LI GM1 cohorts, there was a constant effect whereby LI GM1 patients had a −34.12 ± 15.04-year lower predicted brain age on average compared to Juv GM1 patients. The interaction between biological age and late-infantile GM1 (compared to Juv GM1) showed substantial divergence with an average increase in predicted whole brain age of 11.70 ± 2.63 per year resulting in an average predicted whole brain increase of 12.35 per one year increase in biological age compared to 1.65 in the juvenile GM1 cohort (𝜒^2^(1) = 15.91, *p* < 0.0001, Table 1, Supplement Figure F4). Similar relationships between juvenile and late-infantile GM1 patients in predicted whole brain age were observed for the predicted ages of the white matter, 3^rd^ and 4^th^ ventricles (Figure 3), inferior lateral ventricles, lateral ventricles, external CSF, ventral diencephalon, thalamus, caudate nucleus (Figure 4), nucleus accumbens, putamen, globus pallidus, hippocampus, basal forebrain, amygdala, cerebellar gray and white matter, lobules I-V of the cerebellum, lobules VI and VII of the cerebellum, lobules VIII-X of the cerebellum (Figure 5), and the brainstem, as summarized in Table 1.

### 3.5 BSAGE

Late-infantile GM1 patients had an average cross-sectional whole brain BSAGE = 35.50, juvenile GM1 patients had an average BSAGE = 21.19, and NC participants had an average BSAGE = 0.03 (Figure 6) at the time of most recent MRI scan. An analysis of variance (ANOVA) showed statistically significant (F(2,594) = 671.5, *p* < 0.0001) differences in BSAGE between the cohorts. A post-hoc Tukey test showed that the late-infantile cohort had a statistically significantly higher BSAGE compared to both the neurotypical controls (*p* < 0.0001) and juvenile GM1 patients (*p* < 0.0001). The juvenile GM1 cohort also had a significantly higher BSAGE compared to the neurotypical controls (*p* < 0.0001).

**Figure 6.**
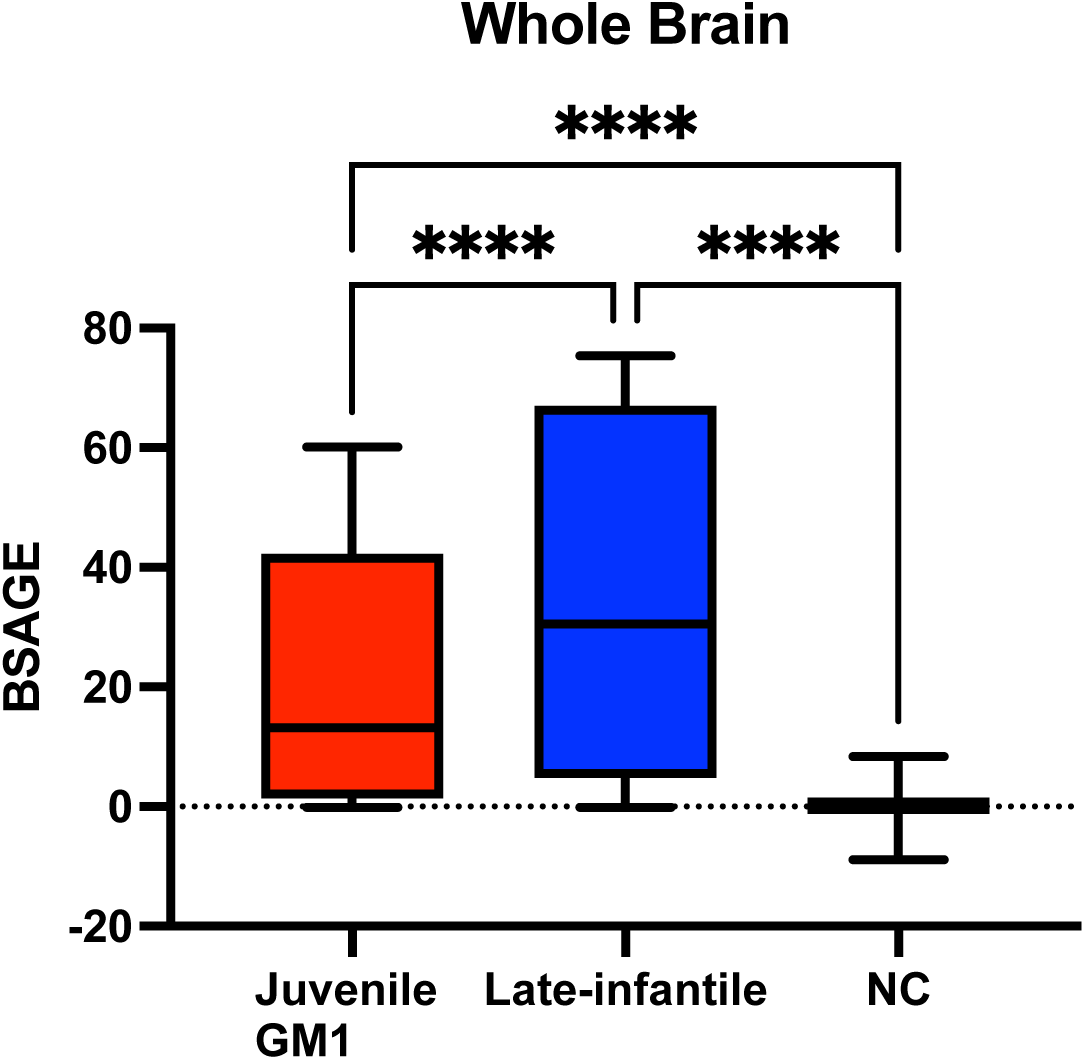
Comparison of Whole Brain BSAGE between Juvenile GM1 patients (n = 26), late- infantile GM1 patients (n = 15), and neurotypical controls (n = 556). Late-infantile GM1 patients had an average BSAGE = 35.50, juvenile GM1 patients had an average BSAGE = 21.19, and NC participants had an average BSAGE = 0.03. An analysis of variance (ANOVA) showed statistically significant (F(2, 595) = 671.5, *p* < 0.0001) differences in BSAGE between the cohorts. A post-hoc Tukey test showed the late-infantile cohort had a statistically significantly higher BSAGE compared to both the neurotypical controls (*p* < 0.0001) and juvenile GM1 patients (*p* < 0.0001). The juvenile GM1 cohort also had a significantly higher BSAGE compared to the neurotypical controls (*p* < 0.0001). Cohort comparisons between LI GM1, juvenile GM1, and NC for BSAGE for the other brain structures can be found in section J of the Supplementary Materials.

Similar relationships between NC, juvenile GM1 patients, and late-infantile patients in whole brain BSAGE were observed for the BSAGE of the white matter, 3^rd^, 4^th^ ventricle, inferior lateral, and lateral ventricles, external CSF, ventral diencephalon, thalamus, caudate nucleus, nucleus accumbens, putamen, globus pallidus, hippocampus, basal forebrain, amygdala, cerebellar gray and white matter, lobules I-V of the cerebellum, lobules VI and VII of the cerebellum, lobules VIII-X of the cerebellum, and the brainstem, as described in Section J of the Supplementary Material.

Longitudinal evaluations of BSAGE (Figure 2B) showed that NC control BSAGE remained close to the x-axis or a 0 value for BSAGE despite increases in biological age. Juvenile GM1 patients, however, demonstrated BSAGE increases commensurate with an increase in chronological age. Furthermore, late-infantile GM1 patients demonstrated an increased slope in the relationship between BSAGE and biological age compared to both NC and juvenile GM1 gangliosidosis patients (Figure 2B).

Comparison of Whole Brain BSAGE among the three NC participants (Supplement Figure E2), showed that the Calgary participants had an average BSAGE = 0.63, QTAB participants had an average BSAGE = 1.48, and QTIM participants had an average BSAGE = −0.93. An analysis of variance (ANOVA) showed statistically significant (F(2,553) = 43.53, *p* < 0.0001) differences in BSAGE between the three NC cohorts. A post-hoc Tukey test showed that the QTAB cohort had a statistically significantly higher BSAGE compared to both Calgary (*p* = 0.009) and QTIM (*p* < 0.0001). The Calgary cohort also had a significantly higher BSAGE compared to QTIM (*p* < 0.0001).

### 3.6 Clinical Correlations

Both the predicted whole brain (ρ(10) = 0.63, *p =* 0.03) and brain structures age gap estimation (BSAGE, ρ(10) = 0.64, *p =* 0.03) positively correlated with baseline CGI-S in LI GM1 gangliosidosis patients (Figure 7). Predicted whole brain (ρ(23) = 0.76, *p <* 0.01) and brain structures age gap estimation (BSAGE, ρ(23) = 0.74, *p <* 0.01) also both positively correlated with baseline CGI-S in juvenile GM1 gangliosidosis patients (Figure 7A and 7B).

**Figure 7.**
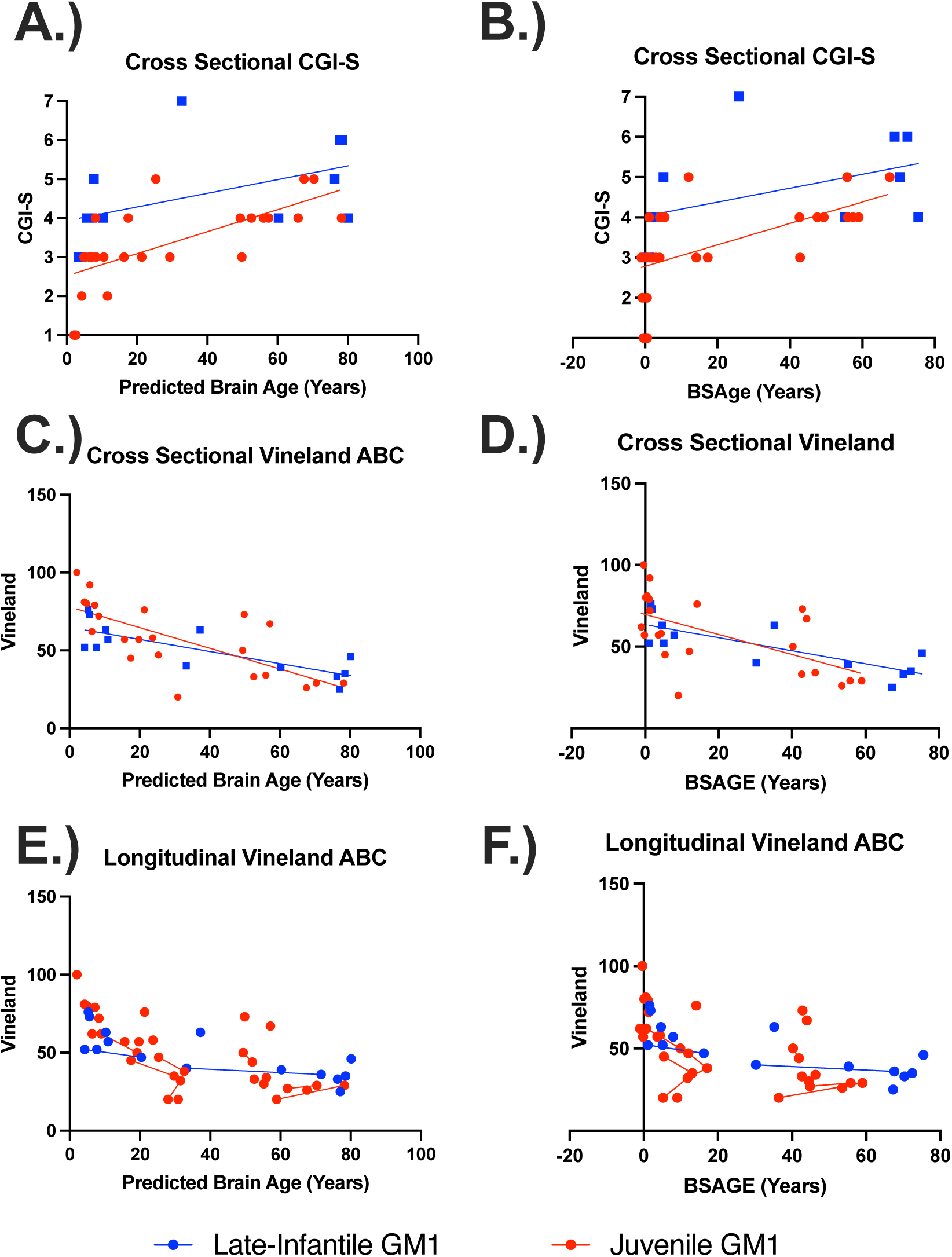
The relationship between clinical outcome assessments and both predicted whole brain age and whole brain BSAGE for late-infantile (blue) and juvenile (red) GM1 patients. BSAGE is equivalent to the patient’s predicted brain age minus their chronological age. (A) Predicted brain age correlated with CGI-S in both the late-infantile (ρ(10) = 0.63; 95% CI [0.06, 0.89], *p =* 0.03) and juvenile (ρ(23) = 0.76; 95% CI [0.51, 0.89], *p <* 0.01) GM1 patients. (B) Brain structures age gap estimation (BSAGE) also correlated with CGI-S in both the late-infantile (ρ(10) = 0.64, *p =* 0.03) and juvenile (ρ(23) = 0.74, *p <* 0.01) GM1 patients. (C) Predicted brain age correlated with cross-sectional Vineland scores in both the late-infantile (ρ(11) = −0.72 95% CI [−0.91, −0.27], *p <* 0.01) and juvenile (ρ(20) = −0.78; 95% CI [−0.90, −0.52], *p <* 0.01) GM1 patients. (D) BSAGE also correlated with cross-sectional Vineland scores in both the late-infantile (ρ(11) = −0.73; 95% CI [−0.92, −0.29], *p <* 0.01) and juvenile (ρ(20) = −0.66; 95% CI [−0.85, −0.31], *p <* 0.01) GM1 patients. (E) Predicted brain age correlated with longitudinal vineland scores in both the late-infantile (𝜒^2^(1) = 12.71, R^2^ = 0.57, *p* < 0.01) and juvenile (𝜒^2^(1) = 12.11, R^2^ = 0.39, *p* < 0.01) GM1 patients. (F) BSAGE also correlated with longitudinal vineland scores in both the late-infantile (𝜒^2^(1) = 12.15, R^2^ = 0.55, *p* < 0.01) and juvenile (𝜒^2^(1) = 4.22, R^2^ = 0.16, *p* = 0.04) GM1 patients.

Predicted whole brain age (ρ(11) = −0.72, *p <* 0.01) and BSAGE (ρ(11) = −0.73, *p <* 0.01) both negatively correlated with cross-sectional Vineland adaptive behavioral composite standard scores in LI GM1 patients. Predicted whole brain age (ρ(20) = −0.78, *p <* 0.01) and BSAGE (ρ(20) = −0.66, *p <* 0.01) also negatively correlated with cross-sectional Vineland adaptive behavioral composite standard scores in juvenile GM1 gangliosidosis patients (Figure 7C and 7D).

Predicted brain age correlated with longitudinal Vineland scores in both the late-infantile (𝜒^2^(1) = 12.71, R^2^ = 0.57, *p* < 0.01) and juvenile (𝜒^2^(1) = 12.11, R^2^ = 0.39, *p* < 0.01) GM1 patients. BSAGE also correlated with longitudinal Vineland scores in both the late-infantile (𝜒^2^(1) = 12.15, R^2^ = 0.55, *p* < 0.01) and juvenile (𝜒^2^(1) = 4.22, R^2^ = 0.16, *p* = 0.04) GM1 patients (Figure 7E and 7F).

### 3.7 Correlations with Brain Volumetrics

Predicted whole brain age was correlated with whole brain volume (cm^3^) in both the juvenile (R^2^ = 0.57) and late-infantile (R^2^ = 0.46) GM1 gangliosidosis patients (Supplement Figure K1). Predicted whole brain age was weakly correlated with whole brain volume (cm^3^) in the NC (R^2^ = 0.02, Supplement Figure K5). Predicted whole brain age was also correlated with whole brain volume controlled for ICV in both the juvenile (R^2^ = 0.77) and late-infantile (R^2^ = 0.51) GM1 gangliosidosis patients (Supplement Figure K2). Predicted whole brain age was also weakly correlated with whole brain volume controlled for ICV in the NC (R^2^ = 0.07, Supplement Figure K6).

Whole brain BSAGE was correlated with whole brain volume (cm^3^) in both the juvenile (R^2^ = 0.49) and late-infantile (R^2^ = 0.43) GM1 gangliosidosis patients (Supplement Figure K3). Whole brain BSAGE was not correlated with whole brain volume (cm^3^) in NC participants (Supplement Figure K7). Whole brain BSAGE was correlated with whole brain volume controlled for ICV in both the juvenile (R^2^ = 0.69) and late-infantile (R^2^ = 0.47) GM1 gangliosidosis patients (Supplement Figure K4). Whole brain BSAGE was weakly correlated with whole brain volume controlled for ICV in the NC (R^2^ = 0.08, Supplement Figure K8).

## Discussion

In this study, we quantified brain aging in the largest cohort of type II GM1 gangliosidosis patients (n = 41, 81 total T1-weighted scans) alongside 556 neurotypical controls (897 T1-weighted scans) of the same age range. We found that GM1 patients and NC have distinct brain aging trajectories; GM1 patients on average showed whole brain aging at approximately twice the rate of their unaffected peers (1.57 compared to 0.83). Furthermore, when examining the type II GM1 gangliosidosis subtypes, we found that the juvenile GM1 patients’ whole brain aged at a rate similar to that of the whole GM1 cohort, but the late-infantile patients aged at nearly eight times that rate (12.35, Figure 1). This relationship accurately depicts the neurologic clinical picture of the two subtypes of GM1 gangliosidosis; late-infantile patients have seizures, speech difficulties, and mobility difficulties earlier and more frequently than their juvenile GM1 counterparts. Similar relationships to that of the whole brain were observed for cerebellar structures including cerebellar gray and white matter, cerebellar lobules I-V, cerebellar lobules VI and VII, cerebellar lobules VIII-X, and the brainstem (Figure 5), midbrain structures including the thalamus, caudate nucleus, putamen, and globus pallidus (Figure 4), and cerebrospinal fluid (CSF) structures including the 3^rd^ ventricle, 4^th^ ventricle, lateral ventricle, inferior lateral ventricle, and external CSF (Figure 3), reflecting the widespread neurodegeneration in Type II GM1 patients. This is consistent with previous volumetric and MRI studies in Type II GM1 gangliosidosis patients that showed atrophy of the cerebrum, cerebellum, corpus callosum, thalamus, caudate nucleus, lentiform nucleus (including the globus pallidus and putamen), associated ventricle enlargement, as well as white matter effects [10–13,18].

Previous investigations into brain aging through brain age predications have demonstrated various neurotypical development findings, including increased brain development during puberty and the capability of predicting mortality [41,42]. Regarding evaluations of the brain age gap or BSAGE as referred to in this study, previous studies have shown that a positive brain age gap is common in neurodegenerative diseases, indicating diagnostic potential [41]. Furthermore, Nguyen et al. [30] utilizing the same *BrainStructuresAges* pipeline used in the present study, found the hippocampus to be the most accelerated aging region in Alzheimer’s patients, the temporal and frontal lobes to have accelerated aging in frontotemporal dementia patients, the thalamus and cerebral gray matter in multiple sclerosis, the prefrontal and medial temporal lobe in schizophrenia patients, and a mostly neurotypical brain in Parkinson’s patients. These results highlight the capability and specificity of the model.

Deviations from neurotypical development in either direction (either underdeveloped or premature development) have been related to worse outcomes. For instance, in toddlers increased BSAGE has been shown to be associated with worse self-regulation, and decreases in BSAGE have been associated with worse cognifition [43]. Furthermore, in a study leveraging data from the adolescent brain cognitive development (ABCD) study, they found increases in T1-weighted derived BSAGE to be associated with socioeconomic disadvantage and neighborhood safety [44]. Decreased BSAGE has also connected to attention-deficit/hyperactivity disorder (ADHD) and depression in adolescents [45]. Cross-sectional BSAGE evaluations from the most recent MRI scan also revealed significant differences between late-infantile and juvenile GM1 gangliosidosis patients. Despite the late-infantile patients being younger than the juvenile patients on average, they showed significantly more aging (Figure 6). Cross-sectional BSAGE evaluations also demonstrated whole brain aging differences among the three NC cohorts (Supplement Figure E2). However, all three cohorts were within two years of true chronological age which was minor compared to either the juvenile (21.19) or late-infantile (35.50) GM1 patients (Figure 6).

Correlations between whole brain volumetrics and predicted whole brain age revealed noteworthy findings (Section K of the Supplementary Materials). First, the predicted whole brain age and BSAGE in NC were at most weakly correlated with whole brain volume. This finding suggests that *BrainStructuresAges* is not solely a univariate function of volume and is potentially a more robust neuroimaging derived aging parameter. Consideration of localized volumes, white and gray matter density, or cortical thickness in combination with brain volume may be more indicative of brain aging [46–48]. Second, in both juvenile and late-infantile patients, predicted whole brain age and BSAGE were correlated with brain volume. This finding indicates that accelerated brain aging is a product of volumetric loss or atrophy in GM1 patients. However, since these findings were not perfectly correlated, brain aging in GM1 gangliosidosis was also not observed to be a univariate function of volume, further suggesting that brain age predictions are more robust than one dimensional volumetric measurements.

Correlations with clinical outcome assessments (Figure 7) including the Vineland Adaptive Behavioral Composite (ABC) and clinical global impression (CGI) scale, further cement the importance of brain aging in this cohort. Increased brain aging was associated with worsening scores in both categories, reflecting both patients’ global clinical presentation and adaptive behavior. The evaluation of brain aging may be an important neuroimaging surrogate outcome assessment for our ongoing gene therapy clinical trial (NCT03952637) [49].

Limitations of this study need to be considered before this study is used in clinical practice, clinical trials, and other research projects assessing neurodegeneration. First, there were variations in scanning protocols between GM1 patients and each of the neurotypical control datasets. GM1 patients, Calgary, and QTAB participants were all scanned on a 3T system, however of different manufacturers (Phillips, General Electrics, and Siemens, respectively). QTIM participants were scanned on 4T Bruker system. Furthermore, the GM1 patients were sedated during their lengthly MRI acquisition protocol, including magnetic resonance spectroscopy (MRS), diffusion tensor imaging (DTI), and functional magnetic resonance imaging (fMRI) collection. Since the present study was focused solely on structural MRI, propofol related effects were likely negligible, but there likely was increased motion in the NC cohorts comparatively. Ultimately, the resolution for GM1 patients (1 mm^3^), Calgary NC (resampled to 0.182 mm^3^), QTAB NC (0.512 mm^3^), and QTIM NC (0.729 mm^3^) were all 1 mm^3^ or better which is sufficient for volumetric analysis [50]. Second, while the whole GM1 cohort was of sufficient sample size (n = 41), there was not an even distribution of late-infantile (n = 15) and juvenile (n = 26) patients. Moreover, longitudinal MRI evaluations showed an even further skew as there were a total of 20 MRI scans in the late-infantile cohort and 61 in the juvenile cohort. This ultimately reflects both the rarity of this disease and the short window between diagnosis which and death which occurs early in the second decade of life in late-infantile patients [7,9]. Despite this challenge, future studies on Type II GM1 gangliosidosis should focus on gathering data from more patients or employ alternative techniques of boosting the sample size to achieve results that increasingly reflect the disease. Future studies investigating brain aging in GM1 gangliosidosis should explore multimodal MRI data including fMRI and DTI, and potentially MRS in evaluations of brain aging [48].

In summary, we quantified brain aging in Type II GM1 gangliosidosis patients in relation to neurotypical controls of the same age range. Brain aging was quantified using a machine learning analysis pipeline capable of predicting brain age in numerous structures throughout the brain. We found widespread accelerated brain aging in GM1 patients compared to neurotypical controls in structures throughout the brain. Late-infantile patients were also found to have accelerated aging compared to juvenile GM1 patients, accurately depicting the clinical phenotypes of these patients. We also correlated brain aging with clinical outcome assessments to bolster predicted age as a neuroimaging surrogate clinical outcome assessment.

## Data Availability

The data described in this manuscript are available from the corresponding author upon reasonable request. Neuroimaging data for Calgary Preschool neurotypical control group are publicly available here: https://osf.io/axz5r/ [23,24]. Neuroimaging data for the Queensland Twin Adolescent Brain (QTAB) is publicly available here: https://openneuro.org/datasets/ds004146/versions/1.0.4 [25,26]. Neuroimaging data for the Queensland Twin IMaging (QTIM) dataset is publicly available here: https://openneuro.org/datasets/ds004169/versions/1.0.6 [27,28].

## Acknowledgements

We thank the participants and their families for the generosity of their time and efforts.

## Ethics Declaration

The NIH Institutional Review Board approved this protocol (02-HG-0107). Informed consent was completed with parents or legal guardians of the patients. All participants were assessed for their ability to provide assent; none were deemed capable.

## Authors’ Contributions

Conceptualization: CJL, CJT, and MTA; Methodology: CJL, PD, JMJ, SIC, MHY, WAG, CJT, and MTA. Data Collection: CJL, PD, JMJ, SIC, MHY, CJT, and MTA; Formal Analysis: CJL, SIC, WAG, CJT, MTA; Investigation: CJL, SIC, WAG, CJT, MTA; Writing (original draft): CJL, WAG, CJT, MTA; Writing (reviewing and editing): CJL, PD, JMJ, SIC, MHY, WAG, CJT, and MTA; Supervision: WAG, CJT, and MTA; Funding: WAG and CJT. All authors have read and agreed to the published version of the manuscript.

## Consent for Publication

Not applicable.

## Supplementary Methods

### Supplement A: Natural History Study Participant Characteristics

Patients were enrolled in the National Human Genome Research Institute (NHGRI) study of the “Natural History of Glycosphingolipid Storage Disorders and Glycoprotein Disorders” (ClinicalTrials.gov ID: NCT00029965).^1^ Demographic information for each of the 41 GM1 patients is listed in Table A1, and the age of the entire cohort is shown in Figure A1.

**Table A1.**
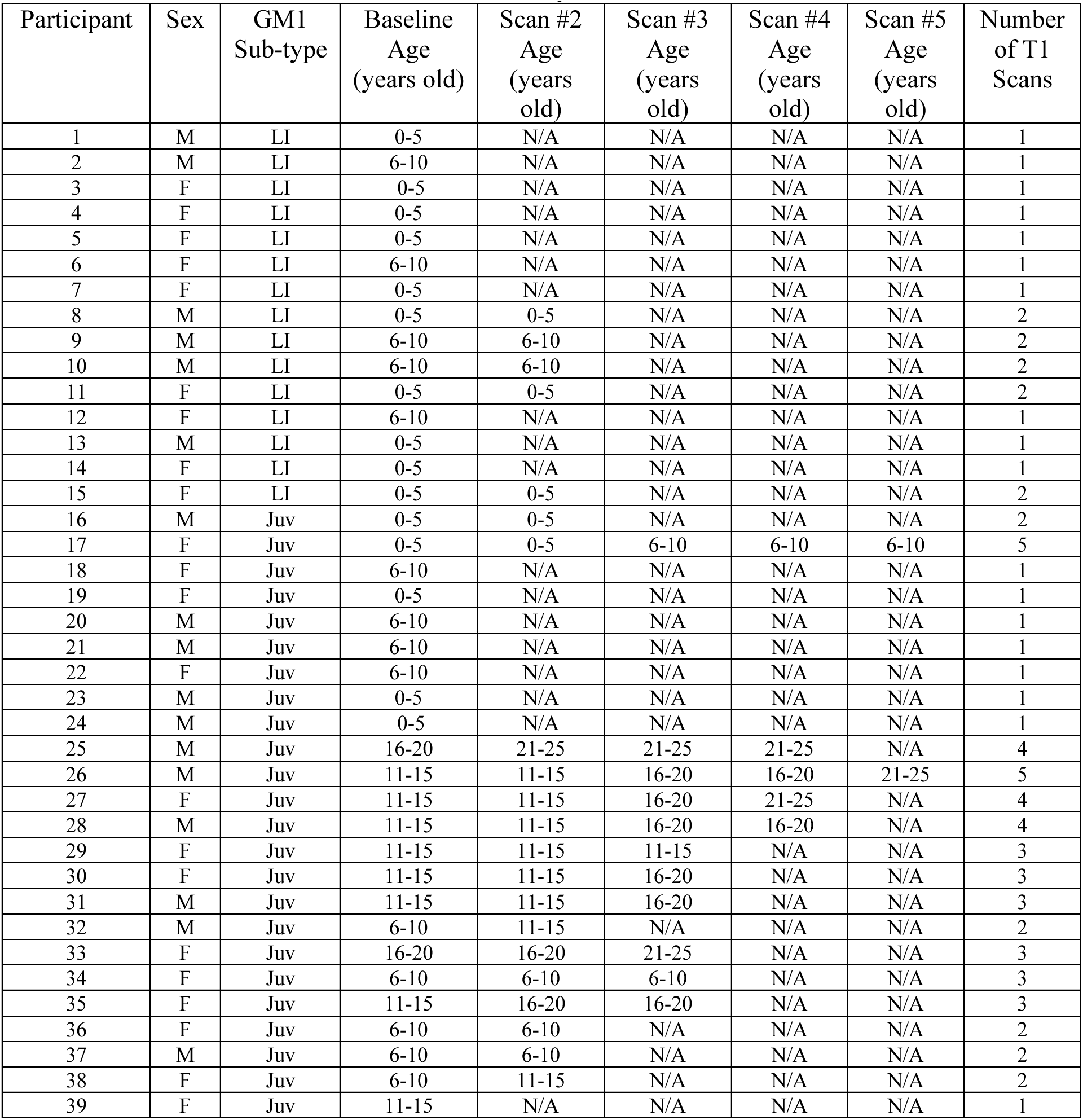
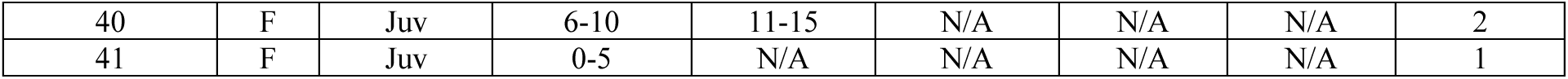
Natural History Study T1-Weighted MRI Scan Age (n = 41). N/A are designated when a participant did not have the above referenced MRI scan. Specific ages were redacted per Medrxiv requirements.

| Participant | Sex | GM1 Sub-type | Baseline Age (years old) | Scan #2 Age (years old) | Scan #3 Age (years old) | Scan #4 Age (years old) | Scan #5 Age (years old) | Number of T1 Scans |
| --- | --- | --- | --- | --- | --- | --- | --- | --- |
| 1 | M | LI | 0-5 | N/A | N/A | N/A | N/A | 1 |
| 2 | M | LI | 6-10 | N/A | N/A | N/A | N/A | 1 |
| 3 | F | LI | 0-5 | N/A | N/A | N/A | N/A | 1 |
| 4 | F | LI | 0-5 | N/A | N/A | N/A | N/A | 1 |
| 5 | F | LI | 0-5 | N/A | N/A | N/A | N/A | 1 |
| 6 | F | LI | 6-10 | N/A | N/A | N/A | N/A | 1 |
| 7 | F | LI | 0-5 | N/A | N/A | N/A | N/A | 1 |
| 8 | M | LI | 0-5 | 0-5 | N/A | N/A | N/A | 2 |
| 9 | M | LI | 6-10 | 6-10 | N/A | N/A | N/A | 2 |
| 10 | M | LI | 6-10 | 6-10 | N/A | N/A | N/A | 2 |
| 11 | F | LI | 0-5 | 0-5 | N/A | N/A | N/A | 2 |
| 12 | F | LI | 6-10 | N/A | N/A | N/A | N/A | 1 |
| 13 | M | LI | 0-5 | N/A | N/A | N/A | N/A | 1 |
| 14 | F | LI | 0-5 | N/A | N/A | N/A | N/A | 1 |
| 15 | F | LI | 0-5 | 0-5 | N/A | N/A | N/A | 2 |
| 16 | M | Juv | 0-5 | 0-5 | N/A | N/A | N/A | 2 |
| 17 | F | Juv | 0-5 | 0-5 | 6-10 | 6-10 | 6-10 | 5 |
| 18 | F | Juv | 6-10 | N/A | N/A | N/A | N/A | 1 |
| 19 | F | Juv | 0-5 | N/A | N/A | N/A | N/A | 1 |
| 20 | M | Juv | 6-10 | N/A | N/A | N/A | N/A | 1 |
| 21 | M | Juv | 6-10 | N/A | N/A | N/A | N/A | 1 |
| 22 | F | Juv | 6-10 | N/A | N/A | N/A | N/A | 1 |
| 23 | M | Juv | 0-5 | N/A | N/A | N/A | N/A | 1 |
| 24 | M | Juv | 0-5 | N/A | N/A | N/A | N/A | 1 |
| 25 | M | Juv | 16-20 | 21-25 | 21-25 | 21-25 | N/A | 4 |
| 26 | M | Juv | 11-15 | 11-15 | 16-20 | 16-20 | 21-25 | 5 |
| 27 | F | Juv | 11-15 | 11-15 | 16-20 | 21-25 | N/A | 4 |
| 28 | M | Juv | 11-15 | 11-15 | 16-20 | 16-20 | N/A | 4 |
| 29 | F | Juv | 11-15 | 11-15 | 11-15 | N/A | N/A | 3 |
| 30 | F | Juv | 11-15 | 11-15 | 16-20 | N/A | N/A | 3 |
| 31 | M | Juv | 11-15 | 11-15 | 16-20 | N/A | N/A | 3 |
| 32 | M | Juv | 6-10 | 11-15 | N/A | N/A | N/A | 2 |
| 33 | F | Juv | 16-20 | 16-20 | 21-25 | N/A | N/A | 3 |
| 34 | F | Juv | 6-10 | 6-10 | 6-10 | N/A | N/A | 3 |
| 35 | F | Juv | 11-15 | 16-20 | 16-20 | N/A | N/A | 3 |
| 36 | F | Juv | 6-10 | 6-10 | N/A | N/A | N/A | 2 |
| 37 | M | Juv | 6-10 | 6-10 | N/A | N/A | N/A | 2 |
| 38 | F | Juv | 6-10 | 11-15 | N/A | N/A | N/A | 2 |
| 39 | F | Juv | 11-15 | N/A | N/A | N/A | N/A | 1 |
| 40 | F | Juv | 6-10 | 11-15 | N/A | N/A | N/A | 2 |
| 41 | F | Juv | 0-5 | N/A | N/A | N/A | N/A | 1 |

**Figure A1.**
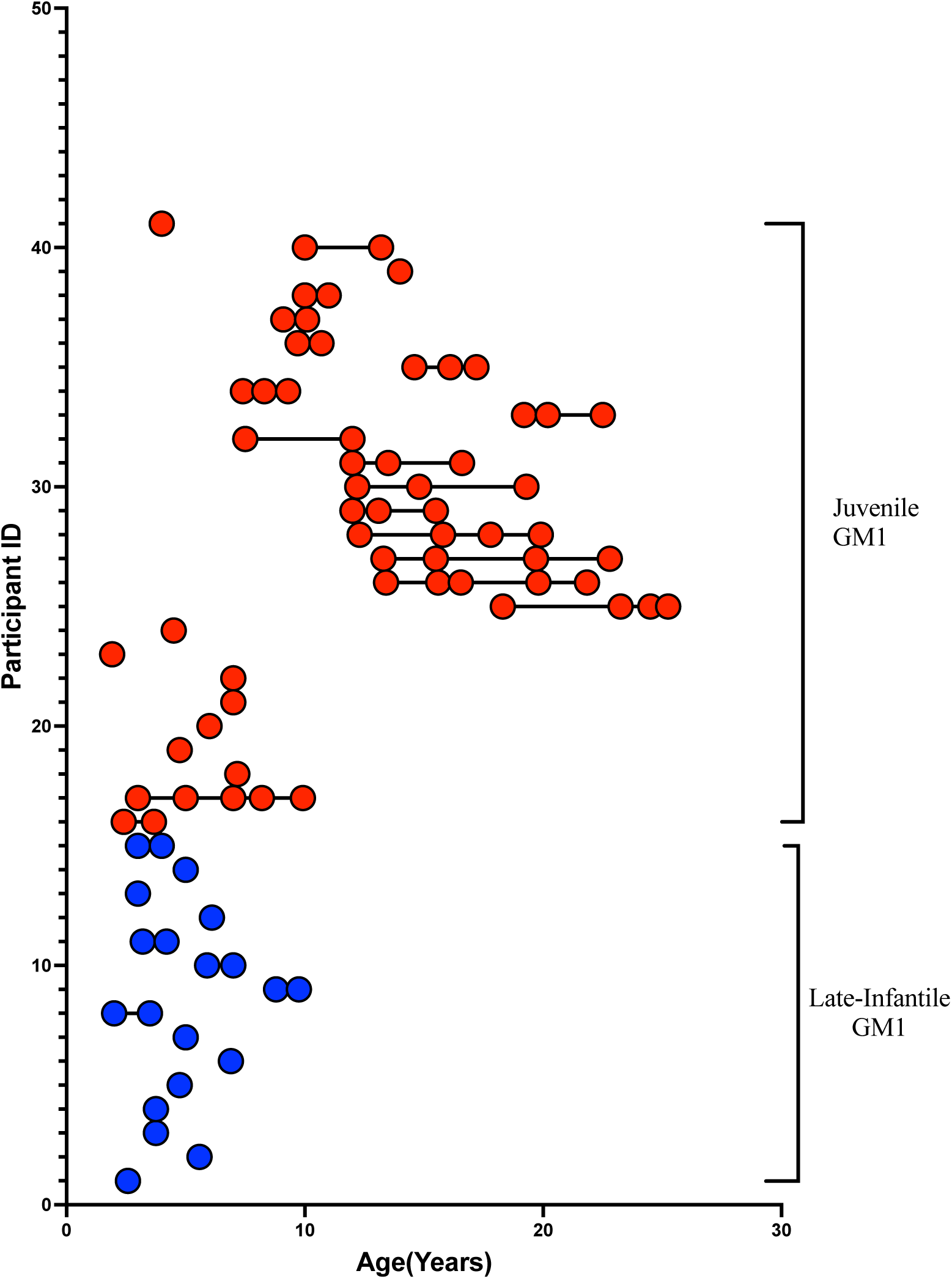
GM1 patient age at each T1-weighted MRI Scan. Late-Infantile (LI, n=15, 20 total MRI scans) GM1 patients are shown in blue and juvenile (n = 26, 61 total MRI scans) GM1 patients are red. Each T1-weighted scan is represented as a circle for all 65 scans; connecting lines indicate that a repeated scan was collected on the same patient. Each patient is shown on a separate row.

### Supplement B: Neurotypical Control Participants

**Table BI.**
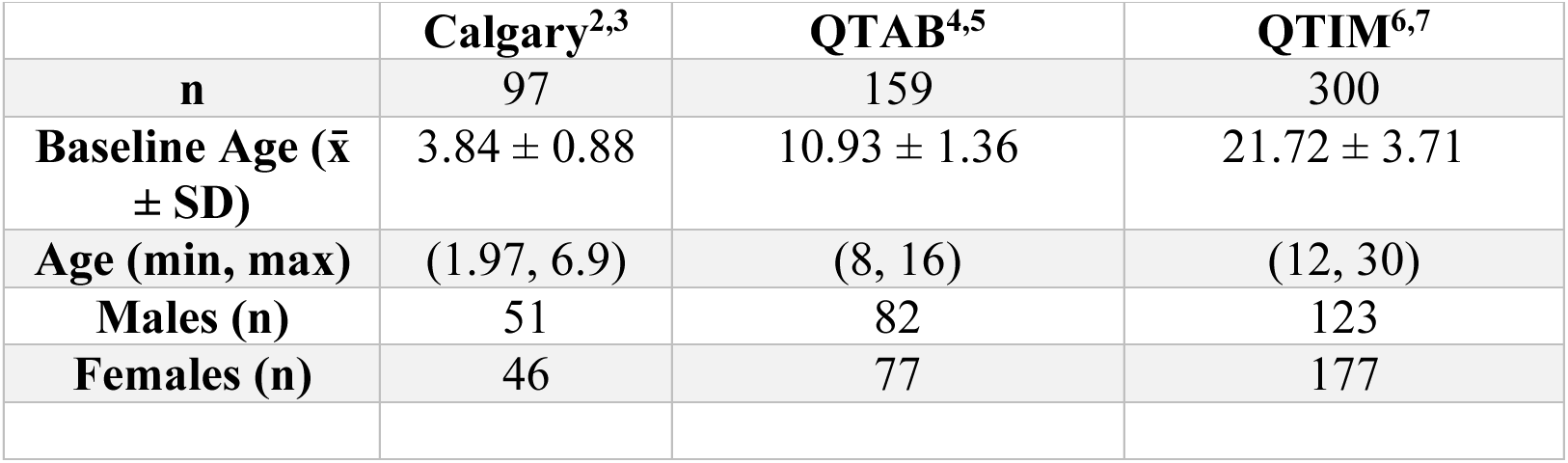
Summary of Neurotypical Control Data Sets.

First, 97 participants (279 scans) from the Calgary Preschool MRI dataset were included consisting of participants between the ages of 2 and 7 years old.^2,3^ Second, 159 participants (318 scans) from the Queensland Twin Adolescent Brain (QTAB) were included in this study, consisting of participants between the ages of 8 and 16 years old.^4,5^ Lastly, 300 participants (300 scans) from the Queensland Twin IMaging (QTIM) were included in this study, consisting of participants between the ages of 12 and 30.^6,7^

**Figure B1.**
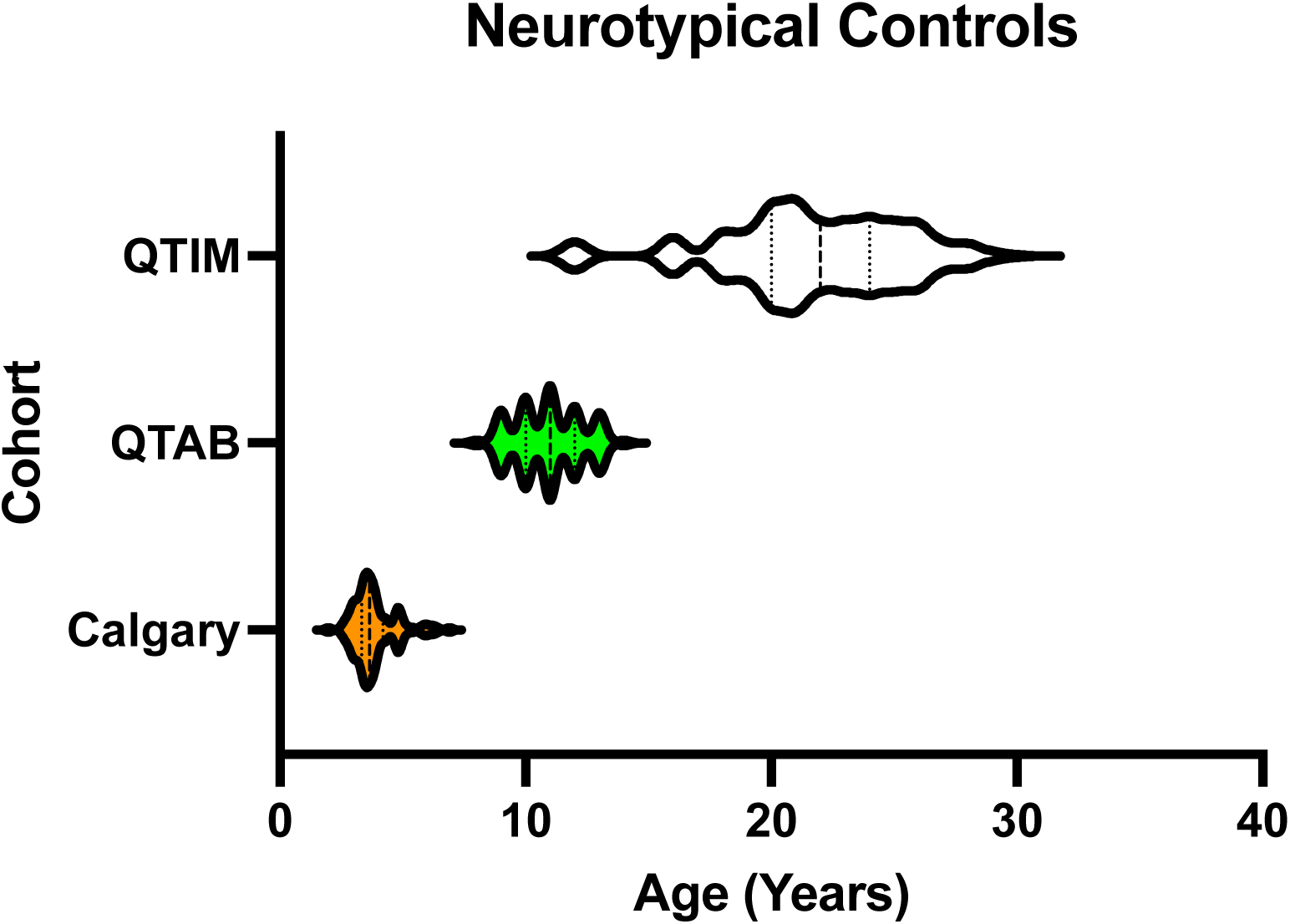
Violin Plot of Neurotypical Control Age. Baseline Scans were used for the Calgary and QTAB Datasets.

### Supplement C: Linear Mixed Effects Modeling

#### Testing the interaction of Cohort and Age on predicted brain age

Linear mixed effects modeling was built in R^8^ with the LME4^9-11^ package. The interaction between the cohort and biological age was tested using the sample code below:

Model 1:

model1 <- lmer(Predicted Brain Age ∼ Age * Cohort + Sex + (1|Participant), dataset) summary(lmer)

Model 2:

model2 <- lmer(Predicted Brain Age ∼ Age + Cohort + Sex + (1|Participant), dataset) summary(lmer)

Likelihood-ratio test:

fm.anova <- anova(model1,model2) summary(fm.anova)

Predicted Brain Age – Corresponds to the predicted brain age determined by

*BrainStructuresAges* for the structure being analyzed

Cohort – Corresponds to the comparison between cohorts as presented in Table 1. Sex – Corresponds to the participants’ biological sex

Age – Corresponds to the participants’ chronological age

Participant – Each participant was given a distinct number to account for repeated measures (subject level random intercept).

## Supplementary Results

### Supplement D: Axial, Coronal, and Anatomical Slices of Brain Aging

**Figure D1.**
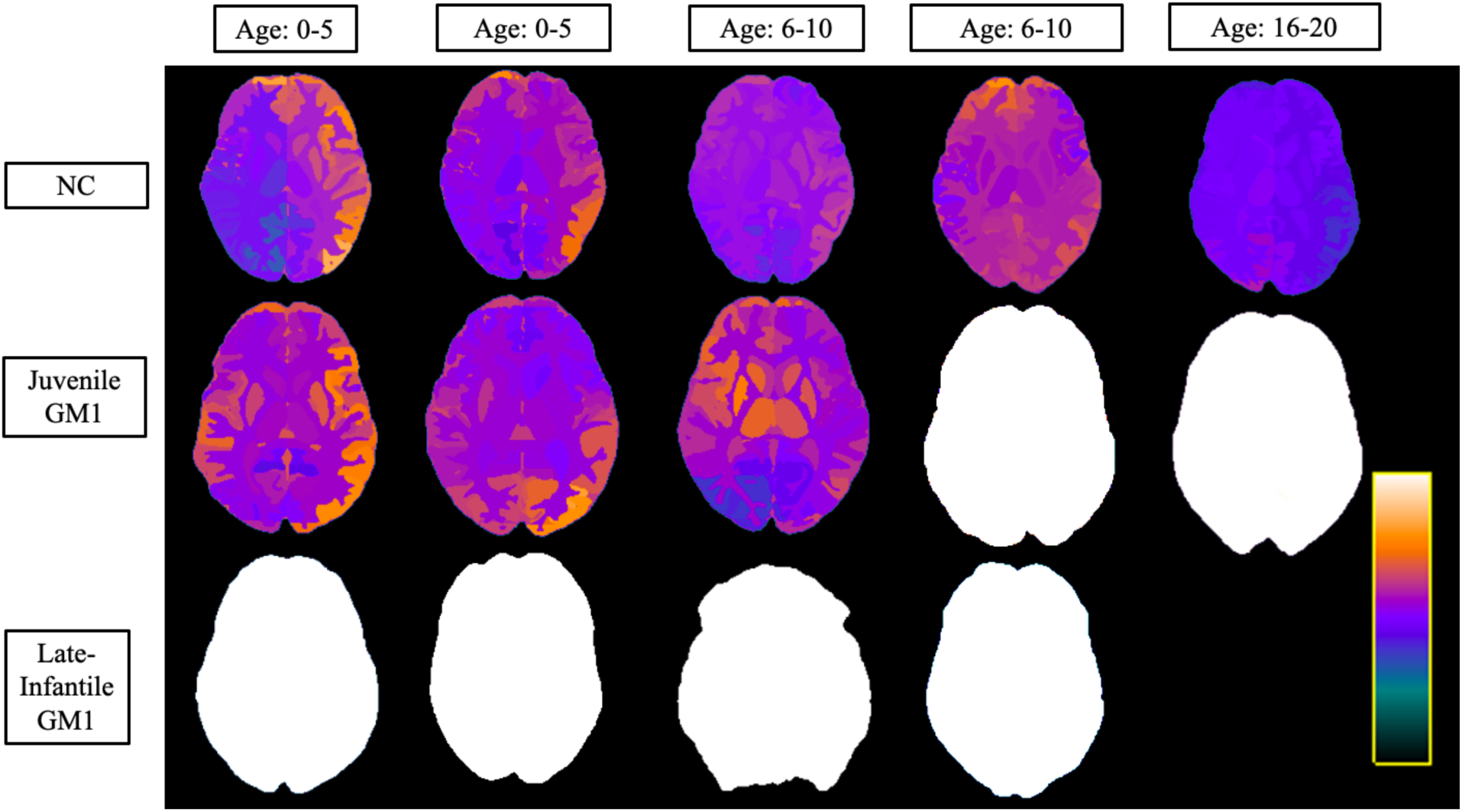
Axial View of Brain Structures Ages of the Three Cohorts at Varying Ages. Neurotypical controls (NC) are shown on the top row, juvenile GM1 gangliosidosis patients are shown in the middle row, and late-infantile GM1 gangliosidosis patients are shown in the bottom row. Colors were assigned according to the predicted age of that brain structure and were assigned between very dark blue (lowest predicted age) and white (highest predicted age) as shown in the scalar bar in the bottom right portion. The scalar ranges were adjusted for each column with min/max values of 0-8 years for the first column (0-5 years old), 0-10 years for the second column (0-5 years old), 0-14 years for the third column (6-10 years old), 0-20 years for the fourth column (6-10 years old), and 0-40 years for the fifth column (16-20 years old). No 20- year-old MRI scan was available for late-infantile patients as the oldest late-infantile patient in this study was 6-10 years old at the time of the MRI evaluation. All MRI images were registered to the MNI coordinate space and slices were taken at z = 6.4 (± 1) mm for each participant. A matching anatomical version of this image is available (Supplement Figure D4). Specific ages were redacted per Medrxiv requirements.

**Figure D2.**
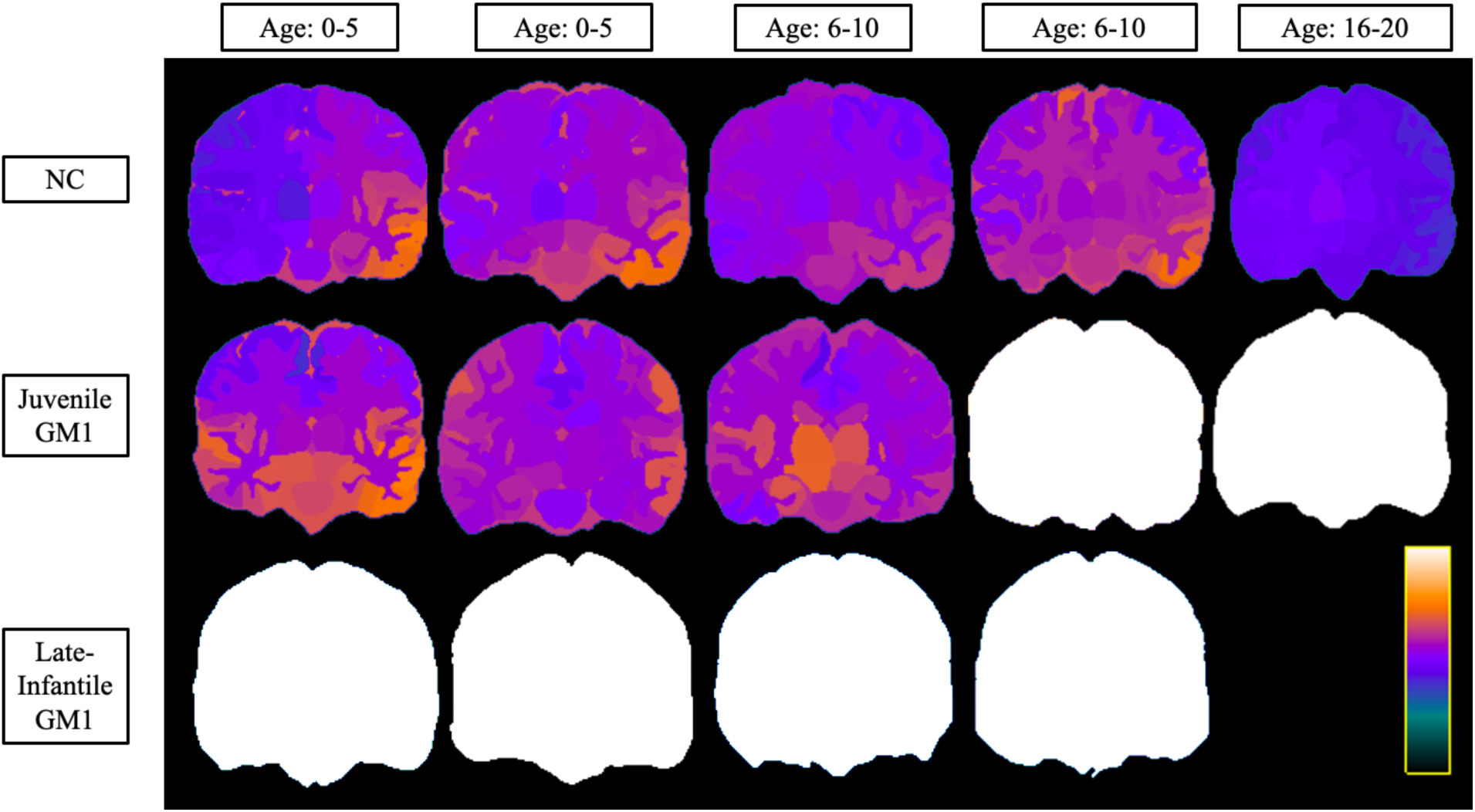
Coronal View of Brain Structures Ages of the Three Cohorts at Varying Ages. Neurotypical controls (NC) are shown on the top row, juvenile GM1 gangliosidosis patients are shown in the middle row, and late-infantile GM1 gangliosidosis patients are shown in the bottom row. Colors were assigned according to the predicted age of that brain structure and were assigned between very dark blue (lowest predicted age) and white (highest predicted age) as shown in the scalar bar in the bottom right portion. The scalar ranges were adjusted for each column with min/max values of 0-8 years for the first column (0-5 years old), 0-10 years for the second column (0-5 years old), 0-14 years for the third column (6-10 years old), 0-20 years for the fourth column (6-10 years old), and 0-40 years for the fifth column (16-20 years old). No 20- year-old MRI scan was available for late-infantile patients as the oldest late-infantile patient in this study was 6-10 years old at the time of the MRI evaluation. All MRI images were registered to the MNI coordinate space and slices were taken at y = −17.6 (± 1) mm for each participant. A matching anatomical version of this image is available (Supplement Figure D5). Specific ages were redacted per Medrxiv requirements.

**Figure D3.**
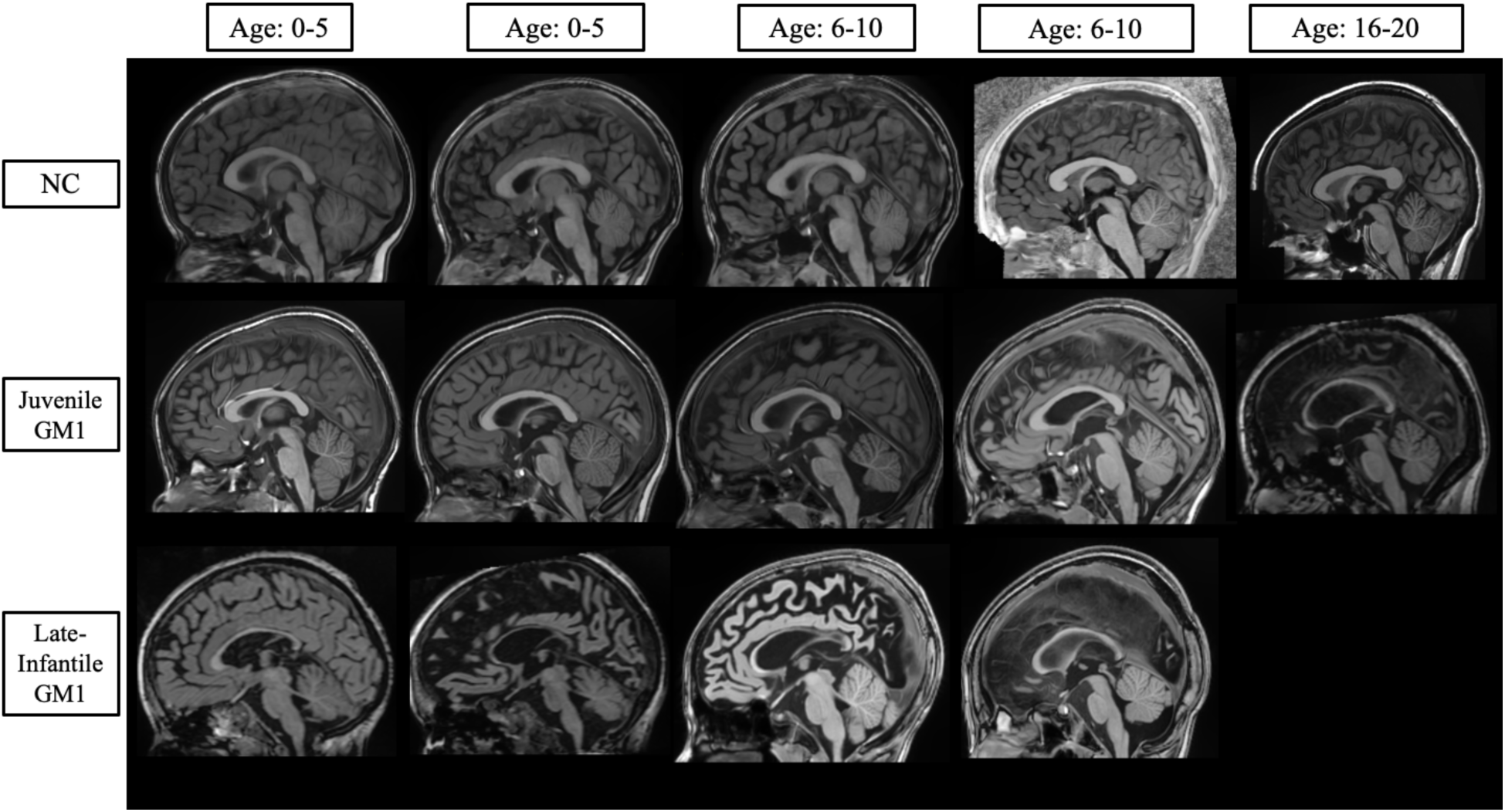
T1-Weighted Sagittal Anatomical Slice of Figure 1. Neurotypical controls (NC) are shown on the top row, juvenile GM1 gangliosidosis patients are shown in the middle row, and late-infantile GM1 gangliosidosis patients are shown in the bottom row. All MRI images were registered to the MNI coordinate space and slices were taken at x = 1.0 (± 1) mm (identical to Figure 2) for each participant. Specific ages were redacted per Medrxiv requirements.

**Figure D4.**
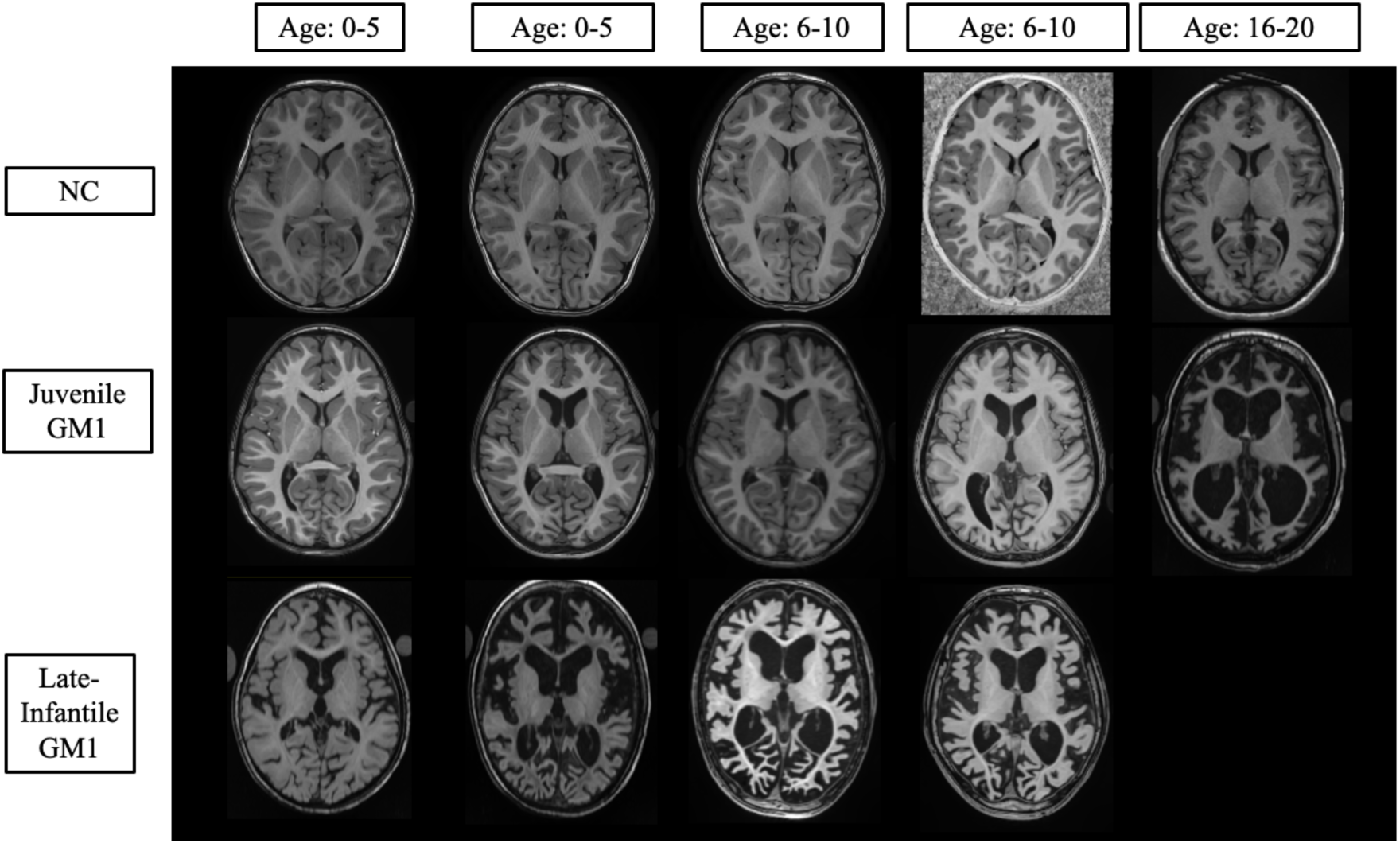
T1-Weighted Axial Anatomical Slice of Figure D1. Neurotypical controls (NC) are shown on the top row, juvenile GM1 gangliosidosis patients are shown in the middle row, and late-infantile GM1 gangliosidosis patients are shown in the bottom row. All MRI images were registered to the MNI coordinate space and slices were taken at z = 6.4 (± 1) mm (identical to Figure D1) for each participant. Specific ages were redacted per Medrxiv requirements.

**Figure D5.**
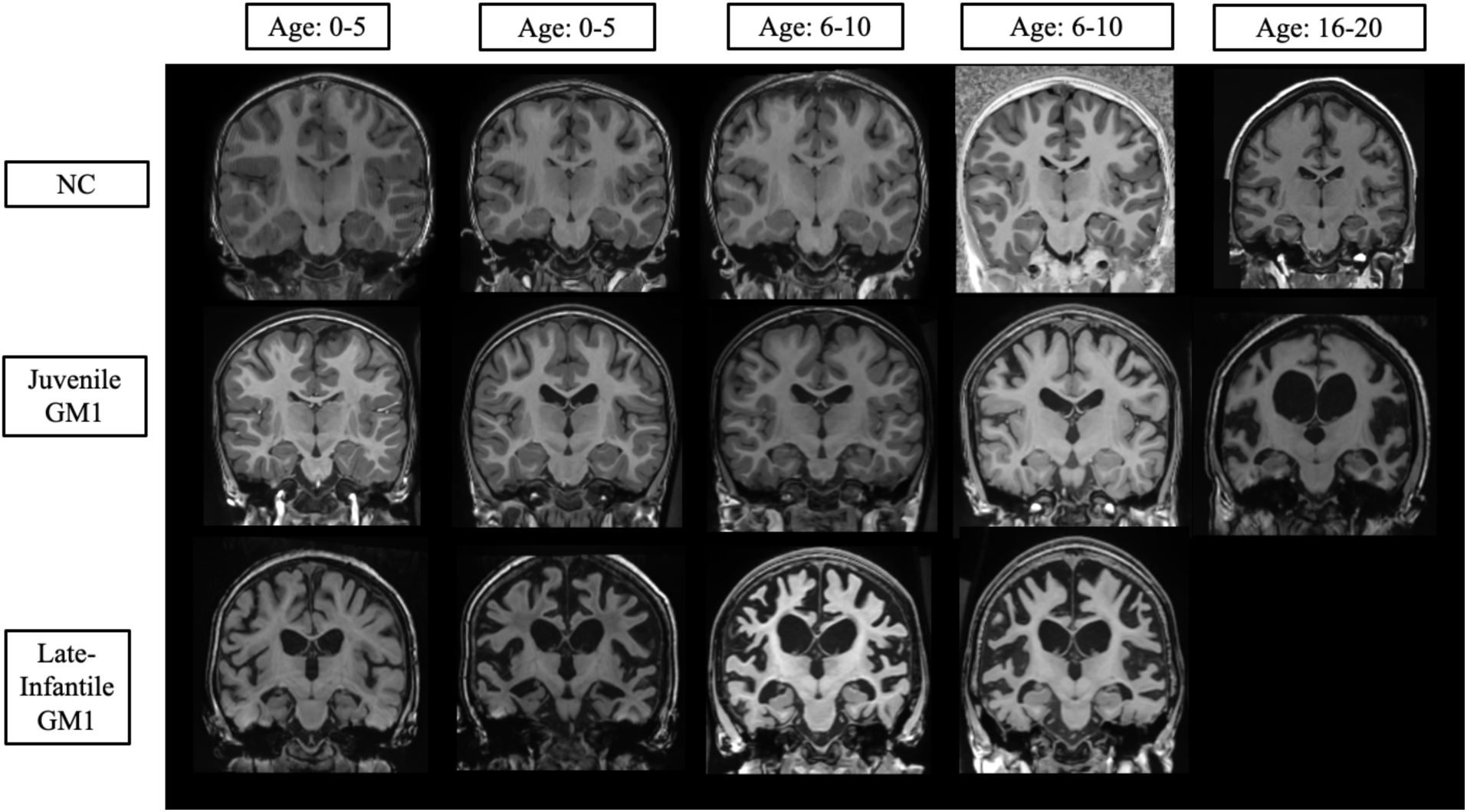
T1-Weighted Coronal Anatomical Slice of Figure D2. Neurotypical controls (NC) are shown on the top row, juvenile GM1 gangliosidosis patients are shown in the middle row, and late-infantile GM1 gangliosidosis patients are shown in the bottom row. All MRI images were registered to the MNI coordinate space and slices were taken at y = −17.6 (± 1) mm (identical to Figure D2) for each participant. Specific ages were redacted per Medrxiv requirements.

### Supplement E: Comparisons between Neurotypical Control Datasets

**Figure E1.**
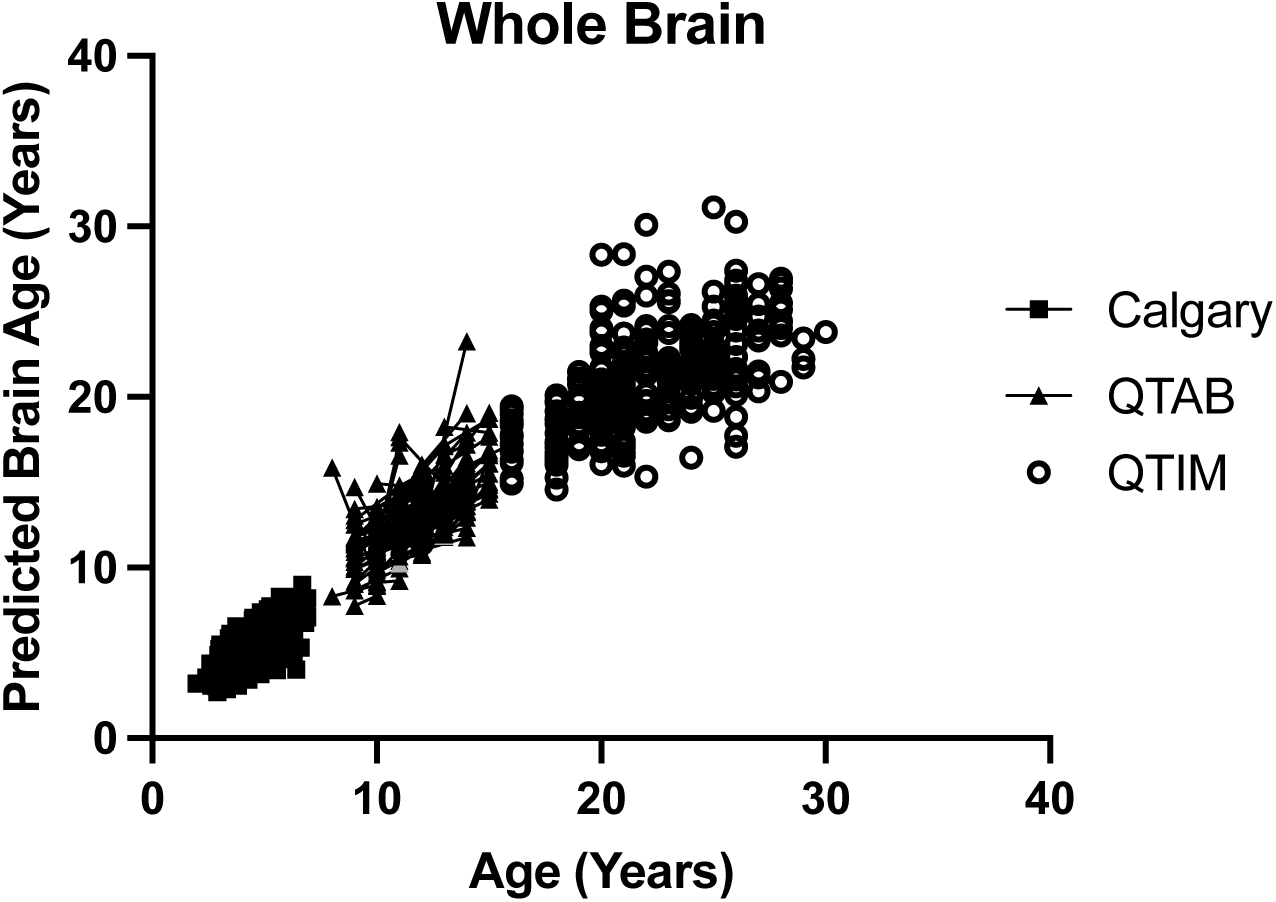
The relationship between predicted brain age and biological age in NC participants (n = 556, 897 total MRI scans). Calgary participants are shown as squares, QTAB participants are shown as triangles, and QTIM participants are shown as open circles. Connecting lines indicate repeated scans on the same participant. Calgary participants showed an average increase of 0.77 (0.03) in predicted brain age per biological year, QTAB participants showed an average increase of 0.81 (0.05) in predicted brain age per biological year, and QTIM participants showed an average increase of 0.65 (0.03) in predicted brain age per biological year

**Figure E2.**
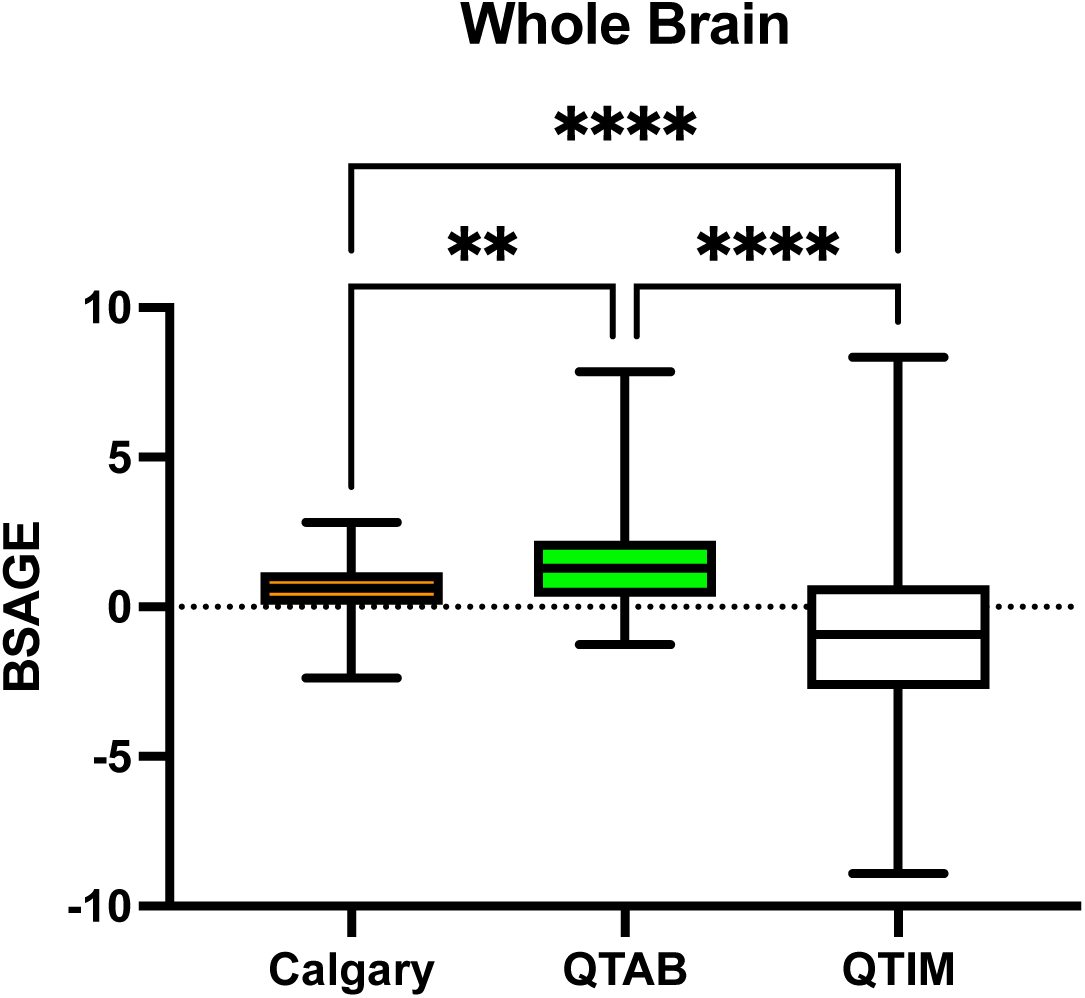
Comparison of Whole Brain BSAGE between NC participants (n = 556). Calgary participants had an average BSAGE = 0.63, QTAB participants had an average BSAGE = 1.48, and QTIM participants had an average BSAGE = −0.93. An analysis of variance (ANOVA) showed statistically significant (F(2, 554) = 43.53, *p* < 0.0001) differences in BSAGE between the three NC cohorts. A post-hoc Tukey test showed the QTAB cohort had a statistically significantly higher BSAGE compared to both Calgary (*p* = 0.009) and QTIM (*p* < 0.0001). The Calgary cohort also had a significantly higher BSAGE compared to QTIM (*p* < 0.0001).

### Supplement F: Linear Mixed Effects Modeling (LMEM) Outputs

**Figure F1.**
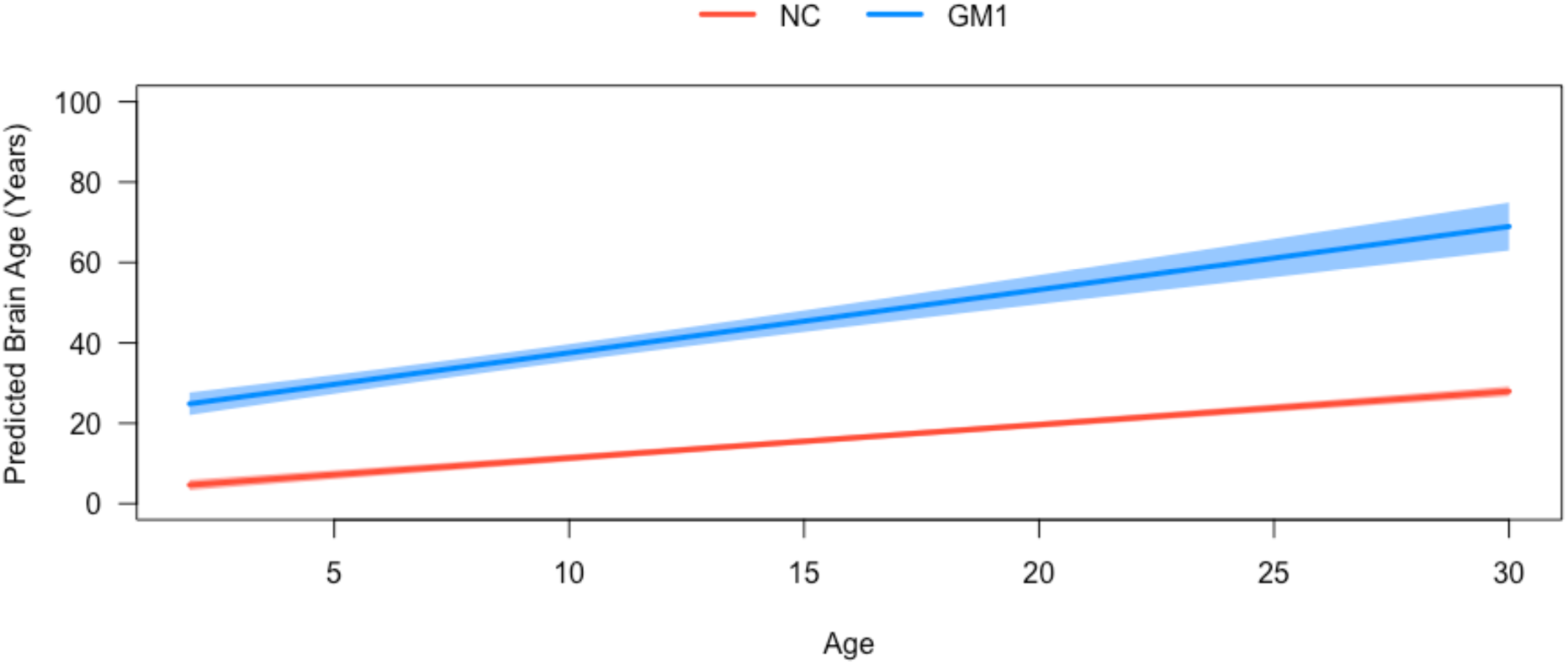
Comparisons of GM1 patients and neurotypical controls (NC) for predicted whole brain age as assessed by LMEM. The GM1 cohort is shown in blue, and the neurotypical controls are shown in red. The shaded regions represent the 95% confidence intervals calculated from the estimates and standards errors defined in Table G1.

**Figure F2.**
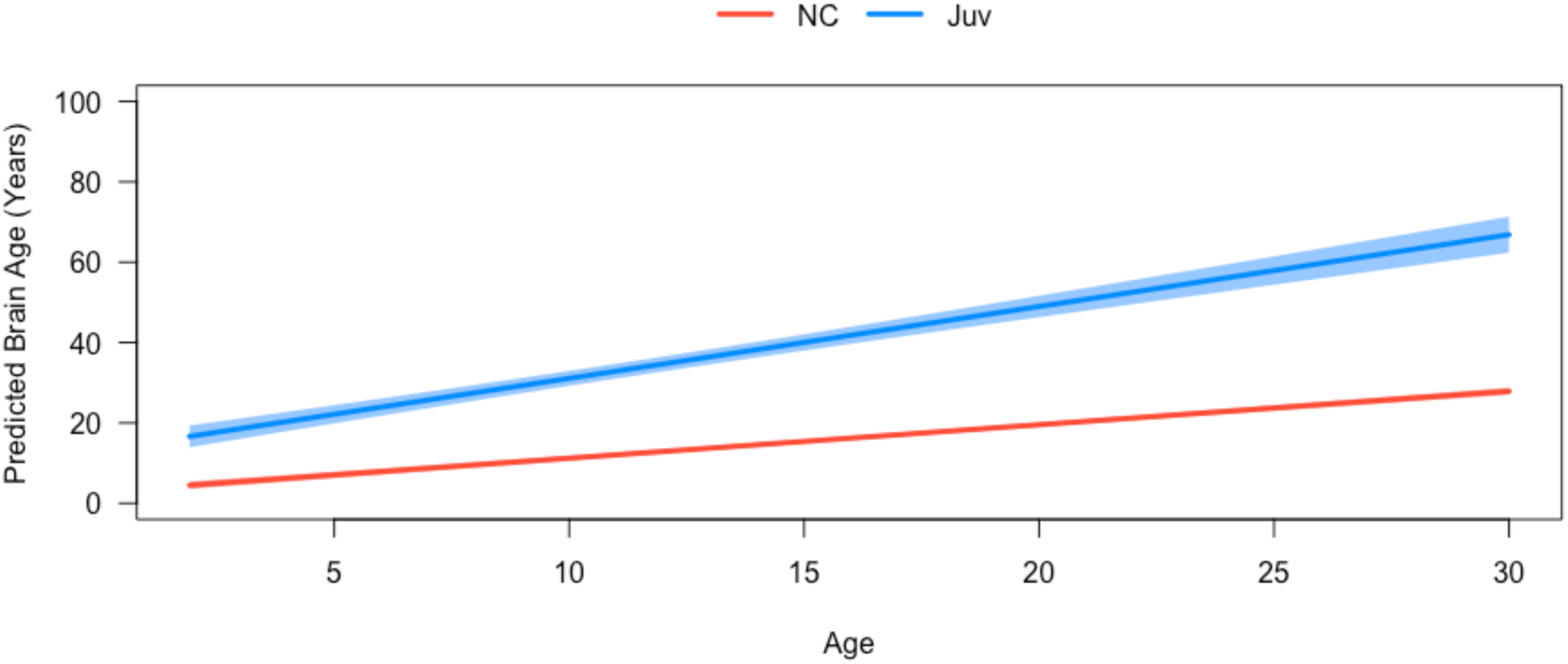
Comparisons of juvenile (Juv) GM1 patients and neurotypical controls (NC) for predicted whole brain age as assessed by LMEM. The Juv GM1 cohort is shown in blue, and the NC are shown in red. The shaded regions represent the 95% confidence intervals calculated from the estimates and standards errors defined in Table G1.

**Figure F3.**
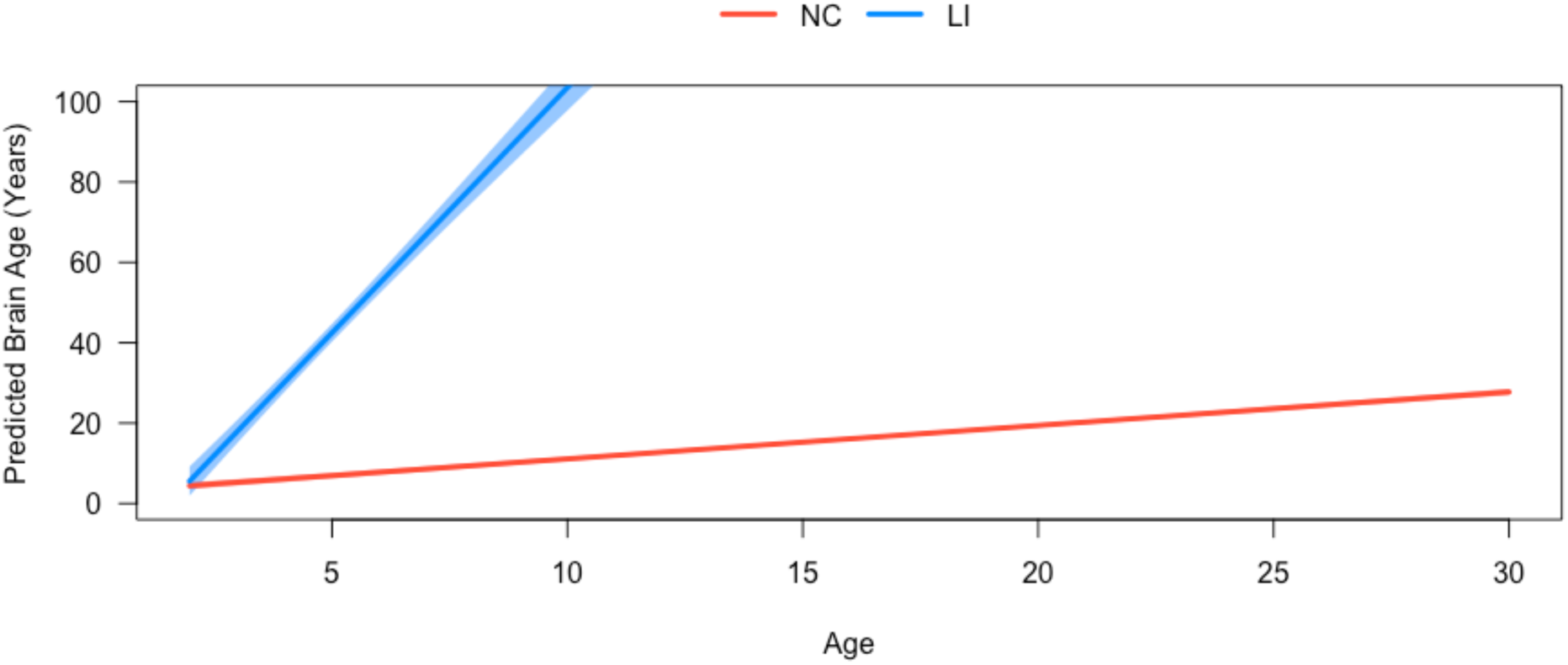
Comparisons of late-infantile (LI) GM1 patients and neurotypical controls for predicted whole brain age as assessed by LMEM. The LI GM1 cohort is shown in blue, and the neurotypical controls are shown in red. The shaded regions represent the 95% confidence intervals calculated from the estimates and standards errors defined in Table G1.

**Figure F4.**
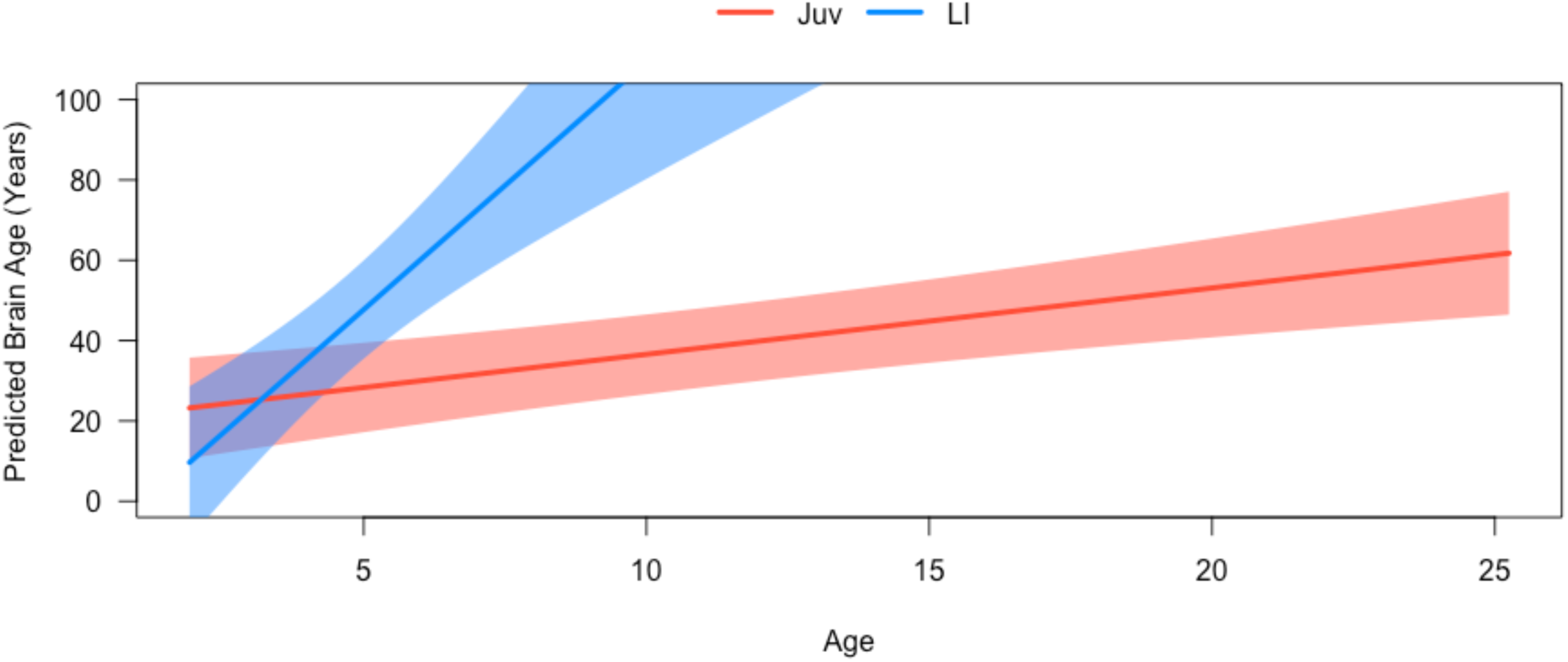
Comparisons of late-infantile (LI) GM1 patients and Juvenile (Juv) GM1 patients for predicted whole brain age as assessed by LMEM. The LI GM1 cohort is shown in blue, and the Juv GM1 patients are shown in red. The shaded regions represent the 95% confidence intervals calculated from the estimates and standards errors defined in Table G1.

### Supplement G: Linear Mixed Effects Modeling (LMEM) Estimates and Standard Errors

**Supplement Table G1.** Estimates and Standard Errors from the LMEM for comparisons of the whole GM1 cohort, Late-Infantile (LI) GM1 patients, and Juvenile (Juv) GM1 patients, and Neurotypical controls (NC) for each of the structures analyzed. The estimates for the intercept, followed by the interaction between the cohort and biological age, biological age alone, the cohort alone, and biological sex are defined in each cell with the standard errors in parenthesis.

| Structure | *GM1 v NC* | *Juv v NC* | *LI v NC* | *Juv v LI* |
| --- | --- | --- | --- | --- |
| Intercept: Estimate (Standard Error) |  |  |  |  |
| Interaction: Estimate (Standard Error) |  |  |  |  |
| Age: Estimate (Standard Error) |  |  |  |  |
| Cohort: Estimate (Standard Error) |  |  |  |  |
| Sex: Estimate (Standard Error) |  |  |  |  |
| **Whole Brain** | 3.06 (0.75)<br>0.74 (0.14)<br>0.83 (0.04)<br>18.77 (1.73)<br>-0.74 (0.57) | 2.90 (0.53)<br>0.96 (0.11)<br>0.83 (0.03)<br>10.31 (1.61)<br>-0.51 (0.40) | 2.79 (0.47)<br>11.37 (0.52)<br>0.83 (0.02)<br>-21.29 (2.76)<br>-0.24 (0.36) | 20.06 (6.93)<br>10.70 (2.63)<br>1.65 (0.42)<br>-34.12 (15.04)<br>-13.06 (6.77) |
| **White Matter** | 3.76 (0.76)<br>0.92 (0.14)<br>0.77 (0.04)<br>18.94 (1.77)<br>-0.76 (0.57) | 3.59 (0.57)<br>1.09 (0.12)<br>0.78 (0.03)<br>12.02 (1.74)<br>-0.47 (0.43) | 3.45 (0.49)<br>10.57 (0.54)<br>0.78 (0.03)<br>-18.32 (2.87)<br>-0.23 (0.38) | 22.81 (7.27)<br>9.67 (2.76)<br>1.73 (0.45)<br>-32.39 (15.78)<br>-13.92 (7.03) |
| **3<sup>rd</sup> Ventricle** | 3.74 (0.77)<br>0.62 (0.13)<br>0.79 (0.04)<br>20.38 (1.70)<br>-0.79 (0.58) | 3.50 (0.53)<br>0.83 (0.11)<br>0.80 (0.03)<br>12.35 (1.57)<br>-0.40 (0.41) | 3.41 (0.49)<br>8.98 (0.53)<br>0.80 (0.02)<br>-9.51 (2.84)<br>-0.22 (0.38) | 23.85 (6.81)<br>8.48 (2.58)<br>1.49 (0.40)<br>-24.57 (14.82)<br>-15.87 (6.87) |
| **4<sup>th</sup> Ventricle** | 5.04 (0.74)<br>0.42 (0.11)<br>0.74 (0.04)<br>17.31 (1.58)<br>-0.73 (0.57) | 4.88 (0.51)<br>0.47 (0.08)<br>0.74 (0.03)<br>9.65 (1.33)<br>-0.46 (0.39) | 4.76 (0.45)<br>11.30 (0.49)<br>0.74 (0.02)<br>-22.22 (2.61)<br>-0.23 (0.35) | 20.68 (5.69)<br>11.40 (2.02)<br>1.15 (0.28)<br>-35.25 (12.03)<br>-13.43 (6.42) |
| **Inferior Lateral Ventricles** | 3.30 (0.76)<br>0.42 (0.11)<br>0.81 (0.04)<br>22.58 (1.61)<br>-0.85 (0.58) | 3.10 (0.56)<br>0.50 (0.10)<br>0.82 (0.03)<br>16.69 (1.53)<br>-0.54 (0.43) | 2.99 (0.48)<br>8.60 (0.45)<br>0.82 (0.02)<br>-7.93 (2.50)<br>-0.25 (0.38) | 27.48 (6.47)<br>8.45 (2.32)<br>1.19 (0.32)<br>-27.43 (13.76)<br>-15.32 (7.22) |
| **Lateral Ventricles** | 5.04 (0.76)<br>0.94 (0.15)<br>0.70 (0.04)<br>17.72 (1.78)<br>-0.70 (0.57) | 4.87 (0.55)<br>1.16 (0.12)<br>0.71 (0.03)<br>9.81 (1.71)<br>-0.40 (0.42) | 4.69 (0.50)<br>10.40 (0.54)<br>0.71 (0.03)<br>-17.86 (2.91)<br>-0.16 (0.38) | 21.95 (7.35)<br>9.36 (2.78)<br>1.74 (0.46)<br>-29.34 (15.90)<br>-14.18 (2.78) |
| **Ventral Diencephalon** | 4.19 (0.75)<br>0.58 (0.13)<br>0.77 (0.04)<br>19.89 (1.68)<br>-0.75 (0.57) | 4.19 (0.75)<br>0.70 (0.10)<br>0.77 (0.04)<br>19.89 (1.68)<br>-0.75 (0.57) | 3.87 (0.46)<br>11.54 (0.50)<br>0.77 (0.02)<br>-21.35 (2.69)<br>-0.21 (0.36) | 23.46 (6.46)<br>11.36 (2.43)<br>1.36 (0.37)<br>-37.08 (14.04)<br>-14.62 (6.73) |
| **External CSF** | 5.14 (0.74)<br>1.04 (0.14)<br>0.73 (0.04)<br>16.45 (1.73)<br>-0.80 (0.56) | 5.14 (0.74)<br>1.20 (0.12)<br>0.73 (0.04)<br>16.45 (1.73)<br>-0.80 (0.56) | 4.85 (0.47)<br>9.76 (0.53)<br>0.74 (0.02)<br>-16.91 (2.80)<br>-0.30 (0.36) | 21.58 (7.03)<br>8.83 (2.67)<br>1.83 (0.44)<br>-29.15 (15.23)<br>-13.29 (6.74) |
| **Thalamus** | 3.34 (0.76) | 3.34 (0.76) | 3.03 (0.50) | 23.50 (6.94) |
|  | 0.63 (0.13)<br>0.80 (0.04)<br>20.65 (1.72)<br>-0.75 (0.58) | 0.81 (0.11)<br>0.80 (0.04)<br>20.65 (1.72)<br>-0.75 (0.58) | 10.57 (0.52)<br>0.80 (0.03)<br>-16.54 (2.82)<br>-0.19 (0.39) | 9.94 (2.62)<br>1.47 (0.40)<br>-31.39 (15.09)<br>-14.55 (7.09) |
| **Caudate Nucleus** | 3.44 (0.78)<br>1.05 (0.15)<br>0.79 (0.04)<br>15.88 (1.84)<br>-0.64 (0.59) | 3.44 (0.78)<br>1.16 (0.13)<br>0.79 (0.04)<br>15.88 (1.84)<br>-0.64 (0.59) | 3.13 (0.53)<br>9.30 (0.60)<br>0.80 (0.03)<br>-17.63 (3.18)<br>-0.07 (0.41) | 21.17 (8.01)<br>8.57 (3.03)<br>1.80 (0.51)<br>-32.17 (17.30)<br>-11.83 (7.58) |
| **Nucleus Accumbens** | 3.82 (0.73)<br>0.93 (0.15)<br>0.79 (0.04)<br>14.63 (1.82)<br>-0.42 (0.56) | 3.82 (0.73)<br>0.98 (0.14)<br>0.79 (0.04)<br>14.63 (1.82)<br>-0.42 (0.56) | 3.50 (0.49)<br>7.37 (0.59)<br>0.79 (0.03)<br>-12.17 (3.11)<br>0.14 (0.38) | 21.01 (8.35)<br>6.68 (3.10)<br>1.65 (0.56)<br>-26.48 (17.68)<br>-9.81 (7.29) |
| **Putamen** | 4.01 (0.78)<br>0.82 (0.14)<br>0.76 (0.04)<br>20.21 (1.78)<br>-0.71 (0.59) | 3.86 (0.55)<br>0.97 (0.12)<br>0.76 (0.03)<br>13.46 (1.68)<br>-0.47 (0.42) | 3.69 (0.50)<br>11.30 (0.54)<br>0.76 (0.03)<br>-21.02 (2.90)<br>-0.12 (0.39) | 25.19 (7.38)<br>10.56 (2.80)<br>1.55 (0.44)<br>-36.99 (16.06)<br>-14.31 (7.39) |
| **Globus Pallidus** | 3.82 (0.78)<br>0.77 (0.14)<br>0.78 (0.04)<br>19.97 (1.78)<br>-0.70 (0.59) | 3.66 (0.55)<br>0.93 (0.42)<br>0.78 (0.03)<br>13.04 (1.66)<br>-0.47 (0.42) | 3.52 (0.51)<br>11.11 (0.55)<br>0.78 (0.03)<br>-20.39 (2.96)<br>-0.12 (0.40) | 24.30 (7.42)<br>10.33 (2.81)<br>1.54 (0.44)<br>-35.49 (16.14)<br>-13.98 (7.43) |
| **Hippocampus** | 4.54 (0.74)<br>0.51 (0.12)<br>0.73 (0.04)<br>21.92 (1.66)<br>-0.52 (0.57) | 4.41 (0.53)<br>0.56 (0.10)<br>0.74 (0.03)<br>16.08 (1.56)<br>-0.28 (0.41) | 4.29 (0.47)<br>10.85 (0.49)<br>0.74 (0.02)<br>-19.08 (2.64)<br>-0.12 (0.37) | 26.36 (6.61)<br>10.67 (2.37)<br>1.22 (0.34)<br>-37.20 (14.07)<br>-13.55 (7.45) |
| **Basal Forebrain** | 4.63 (0.78)<br>0.84 (0.15)<br>0.76 (0.04)<br>19.11 (1.82)<br>-0.62 (0.59) | 4.49 (0.56)<br>0.98 (0.12)<br>0.76 (0.03)<br>12.09 (1.71)<br>-0.39 (0.43) | 4.30 (0.49)<br>12.04 (0.56)<br>0.76 (0.03)<br>-24.73 (2.95)<br>-0.05 (0.38) | 23.93 (7.36)<br>11.63 (2.79)<br>1.60 (0.45)<br>-40.74 (15.97)<br>-14.00 (7.17) |
| **Amygdala** | 4.70 (0.78)<br>0.68 (0.13)<br>0.76 (0.04)<br>19.48 (1.70)<br>-0.66 (0.59) | 4.70 (0.78)<br>0.77 (0.11)<br>0.76 (0.04)<br>19.48 (1.70)<br>-0.66 (0.59) | 4.34 (0.49)<br>10.50 (0.51)<br>0.76 (0.03)<br>-18.77 (2.77)<br>-0.06 (0.38) | 25.70 (6.83)<br>10.07 (2.53)<br>1.37 (0.37)<br>-35.18 (14.77)<br>-14.47 (7.32) |
| **Cerebellar Gray Matter** | 6.36 (0.71)<br>0.38 (0.11)<br>0.65 (0.04)<br>16.61 (1.55)<br>-0.72 (0.54) | 6.36 (0.71)<br>0.44 (0.08)<br>0.65 (0.04)<br>16.61 (1.55)<br>-0.72 (0.54) | 6.09 (0.45)<br>10.61 (0.49)<br>0.65 (0.02)<br>-20.90 (2.63)<br>-0.20 (0.35) | 21.80 (5.60)<br>10.89 (2.05)<br>1.04 (0.29)<br>-34.45 (12.02)<br>-13.46 (6.11) |
| **Cerebellar White Matter** | 5.55 (0.73)<br>0.43 (0.12)<br>0.70 (0.04)<br>17.33 (1.59)<br>-0.77 (0.56) | 5.55 (0.73)<br>0.46 (0.08)<br>0.70 (0.04)<br>17.33 (1.59)<br>-0.77 (0.56) | 5.26 (0.45)<br>11.04 (0.50)<br>0.70 (0.02)<br>-21.60 (2.65)<br>-0.23 (0.35) | 21.66 (5.80)<br>11.32 (2.10)<br>1.12 (0.29)<br>-35.91 (12.38)<br>-13.77 (6.43) |
| **Cerebellar Lobule I-V** | 4.42 (0.76)<br>0.56 (0.13)<br>0.76 (0.04)<br>20.54 (1.67)<br>-0.80 (0.58) | 4.42 (0.76)<br>0.64 (0.10)<br>0.76 (0.04)<br>20.54 (1.67)<br>-0.80 (0.58) | 4.09 (0.47)<br>11.22 (0.49)<br>0.76 (0.02)<br>-20.07 (2.67)<br>-0.23 (0.37) | 25.40 (6.49)<br>10.93 (2.41)<br>1.30 (0.35)<br>-36.45 (14.02)<br>-15.26 (6.94) |
| **Cerebellar Lobule VI-VII** | 5.13 (0.71)<br>0.45 (0.11)<br>0.72 (0.04)<br>17.44 (1.55)<br>-0.70 (0.54) | 5.13 (0.71)<br>0.52 (0.08)<br>0.72 (0.04)<br>17.44 (1.55)<br>-0.70 (0.54) | 4.86 (0.46)<br>10.46 (0.50)<br>0.73 (0.02)<br>-18.56 (2.67)<br>-0.23 (0.35) | 20.95 (5.68)<br>10.52 (2.09)<br>1.21 (0.30)<br>-31.90 (12.23)<br>-13.56 (6.15) |
| **Cerebellar Lobule VIII-X** | 5.24 (0.70)<br>0.38 (0.11) | 5.24 (0.70)<br>0.44 (0.08) | 4.97 (0.45)<br>9.57 (0.47) | 20.94 (5.58)<br>9.42 (2.02) |
|  | 0.72 (0.04)<br>17.00 (1.50)<br>-0.73 (0.54) | 0.72 (0.04)<br>17.00 (1.50)<br>-0.73 (0.54) | 0.72 (0.02)<br>-15.69 (2.54)<br>-0.23 (0.35) | 1.12 (0.28)<br>-27.54 (11.92)<br>-13.40 (6.18) |
| **Brainstem** | 4.80 (0.75)<br>0.51 (0.13)<br>0.75 (0.04)<br>17.99 (1.65)<br>-0.73 (0.57) | 4.63 (0.52)<br>0.58 (0.09)<br>0.75 (0.03)<br>10.25 (1.40)<br>-0.45 (0.40) | 4.50 (0.46)<br>11.83 (0.51)<br>0.75 (0.02)<br>-23.93 (2.72)<br>-0.20 (0.35) | 21.36 (6.07)<br>12.17 (2.24)<br>1.26 (0.32)<br>-39.36 (13.09)<br>-13.89 (6.56) |

### Supplement H: Ventral Diencephalon Predicted Brain Age

**Figure H1.**
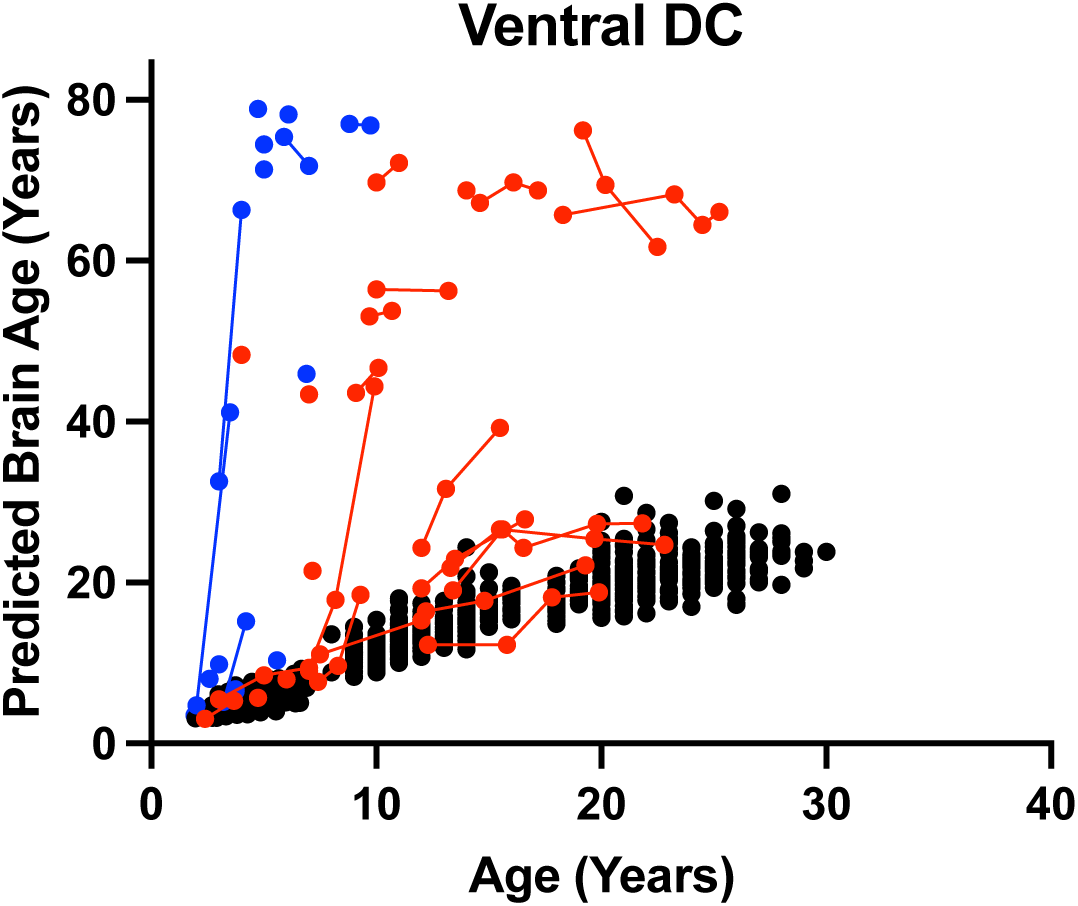
The relationship between predicted brain age and biological age in the Ventral Diencephalon (DC) plotted against chronological age. Late-infantile (n = 15, 20 total MRI scans) GM1 gangliosidosis patients are shown in blue, juvenile (n = 26, 61 total MRI scans) GM1 gangliosidosis patients are shown in red, and neurotypical controls (n = 556, 897 total MRI scans) participants are shown in black. Connecting lines indicate repeated scans on the same participant. Statistical analysis was performed using a linear mixed effects model to test the interactions between chronological age and cohort on predicted brain age and the results are summarized in Table I, with the estimated and standard errors shown in Table G1 of the Supplementary Materials.

### Supplement I: Longitudinal Brain structures age gap estimation (BSAGE)

**Figure I1.**
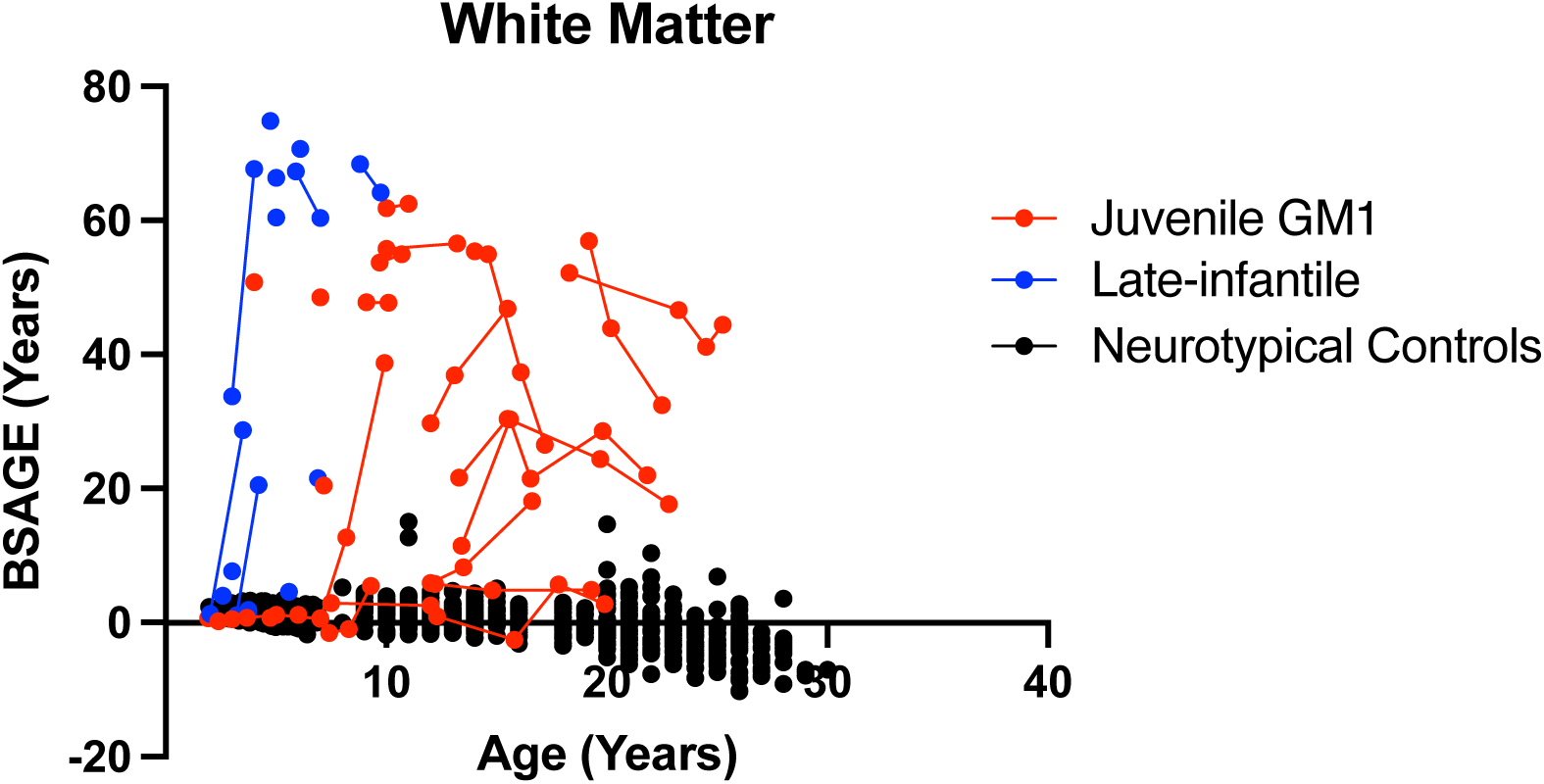
The longitudinal relationship between white matter BSAGE and chronological age. Late-infantile GM1 gangliosidosis patients are shown in blue, juvenile GM1 gangliosidosis patients are shown in red, and neurotypical controls participants are shown in black. Connecting lines indicate repeated scans on the same participant.

**Figure I2.**
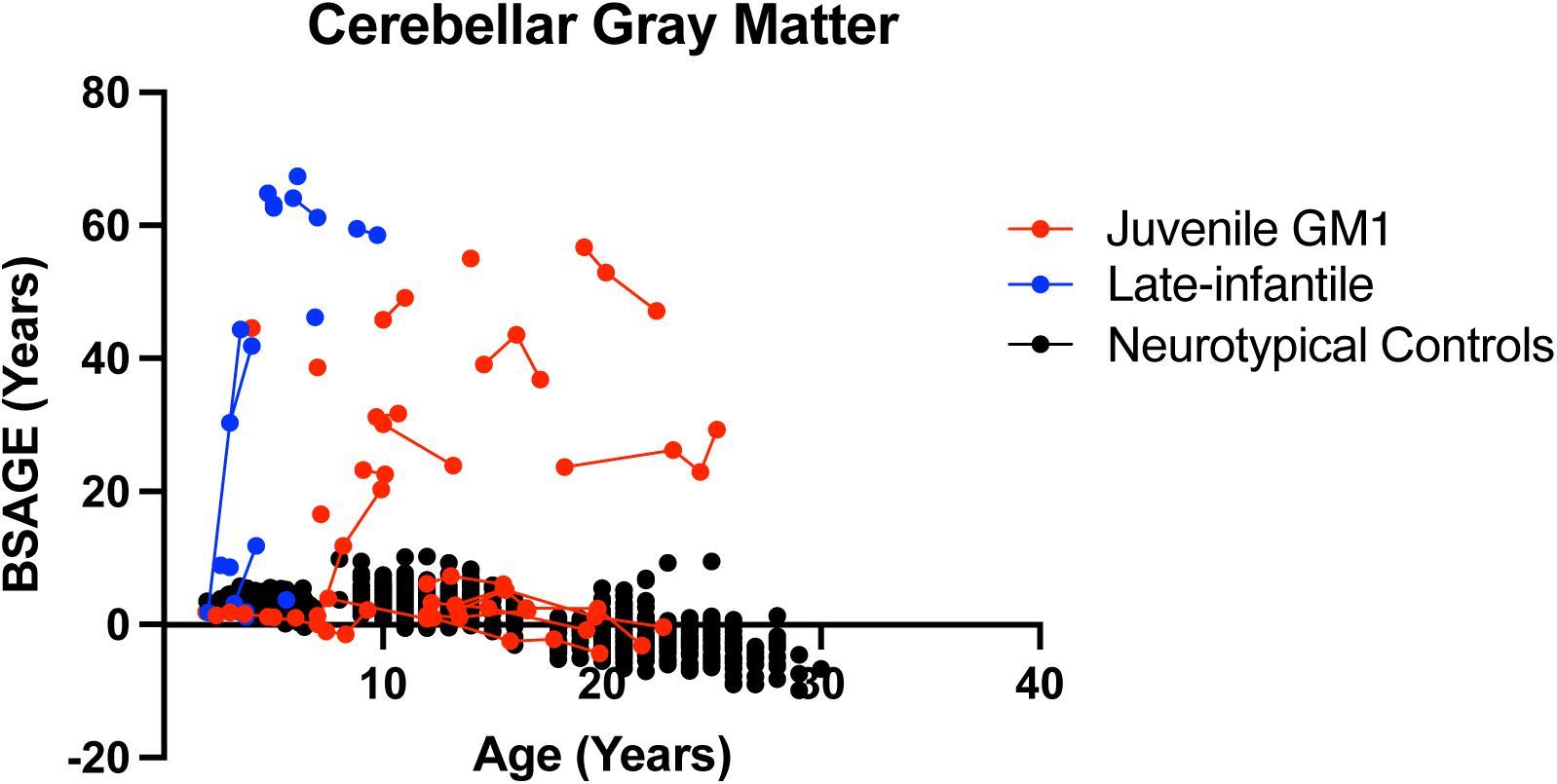
The longitudinal relationship between cerebellar gray matter BSAGE and chronological age. Late-infantile GM1 gangliosidosis patients are shown in blue, juvenile GM1 gangliosidosis patients are shown in red, and neurotypical controls participants are shown in black. Connecting lines indicate repeated scans on the same participant.

**Figure I3.**
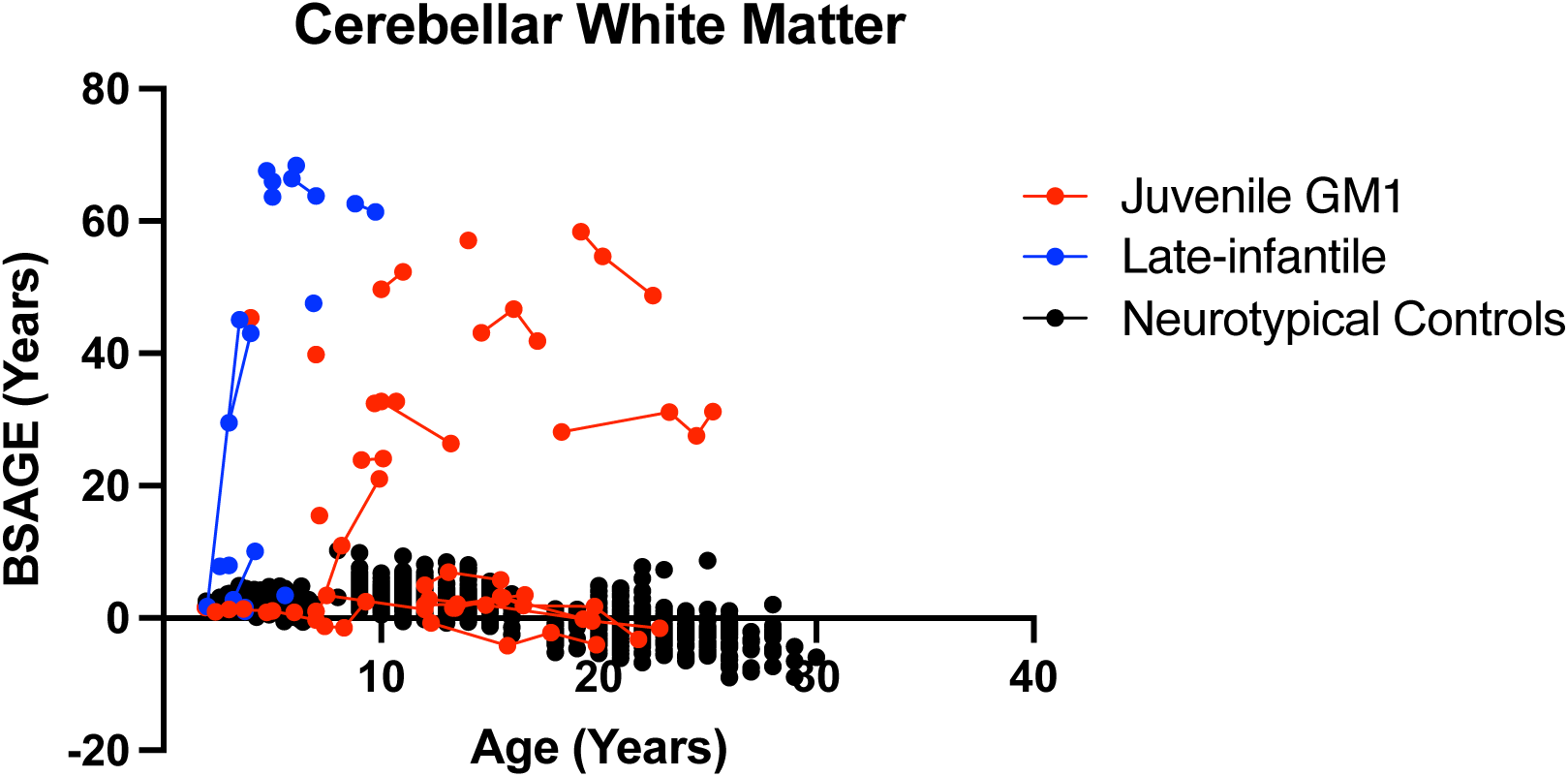
The longitudinal relationship between cerebellar white matter BSAGE and chronological age. Late-infantile GM1 gangliosidosis patients are shown in blue, juvenile GM1 gangliosidosis patients are shown in red, and neurotypical controls participants are shown in black. Connecting lines indicate repeated scans on the same participant.

**Figure I4.**
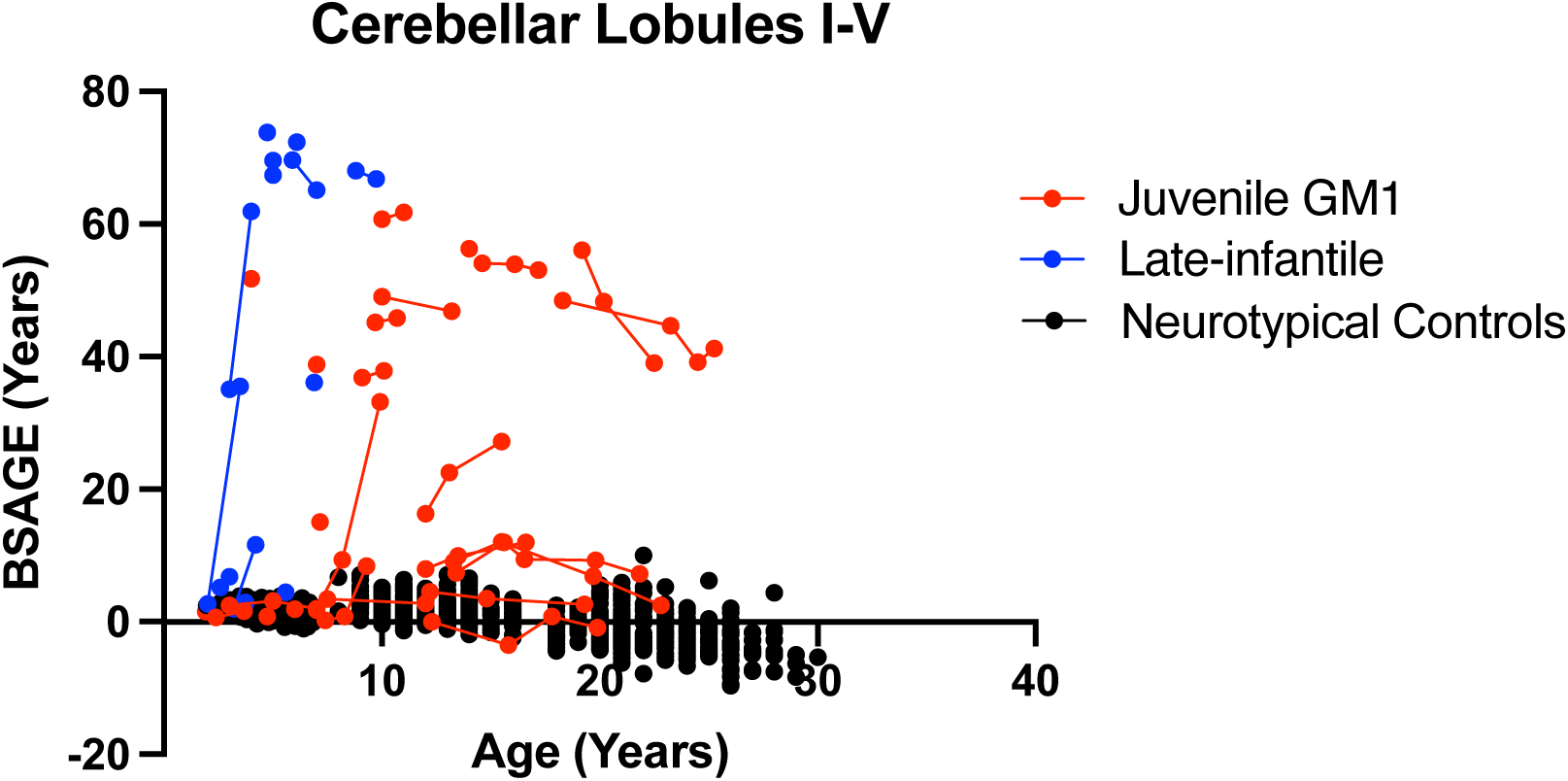
The longitudinal relationship between cerebellar lobules I-V BSAGE and chronological age. Late-infantile GM1 gangliosidosis patients are shown in blue, juvenile GM1 gangliosidosis patients are shown in red, and neurotypical controls participants are shown in black. Connecting lines indicate repeated scans on the same participant.

**Figure I5.**
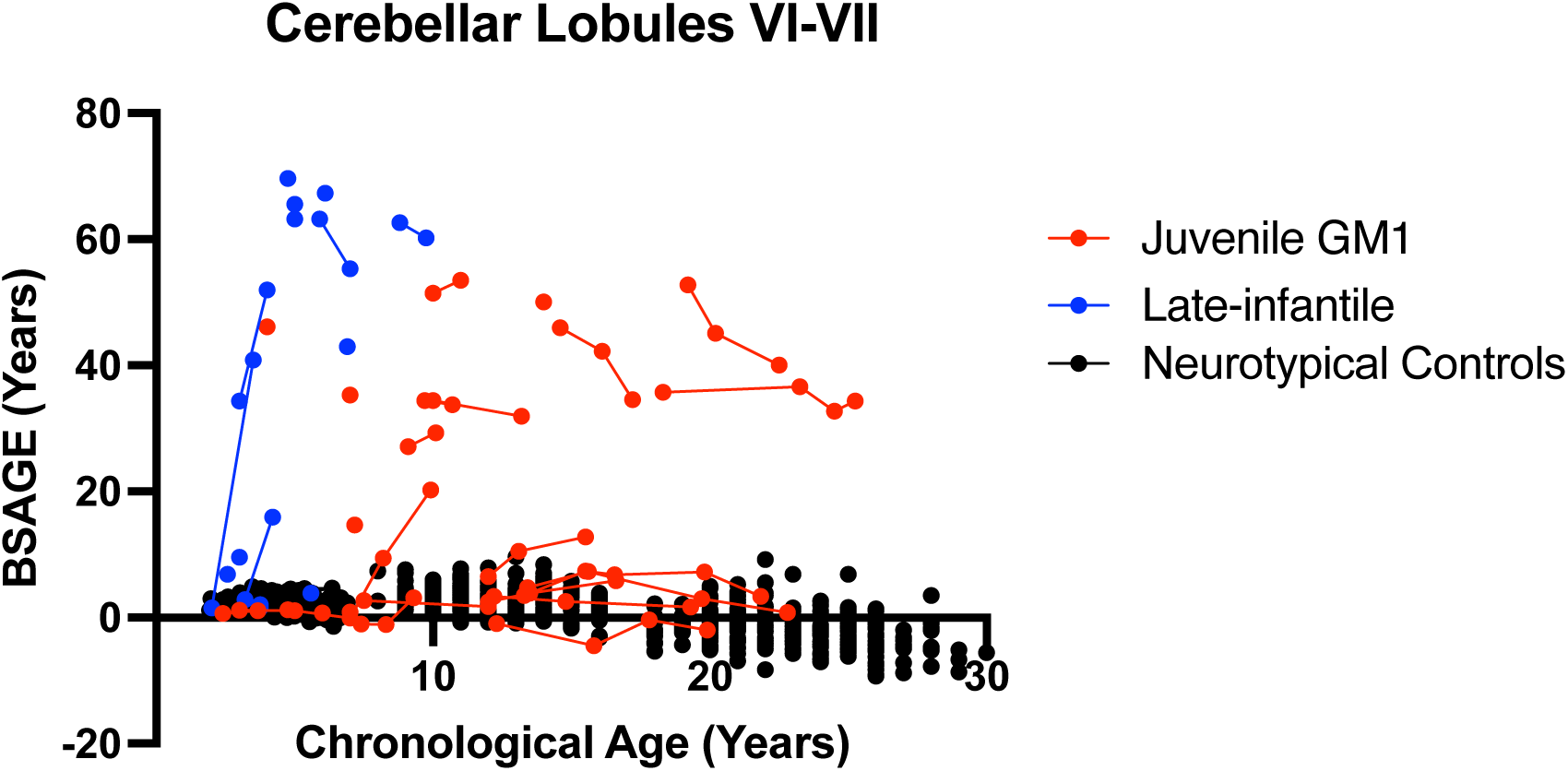
The longitudinal relationship between cerebellar lobules VI-VII BSAGE and chronological age. Late-infantile GM1 gangliosidosis patients are shown in blue, juvenile GM1 gangliosidosis patients are shown in red, and neurotypical controls participants are shown in black. Connecting lines indicate repeated scans on the same participant.

**Figure I6.**
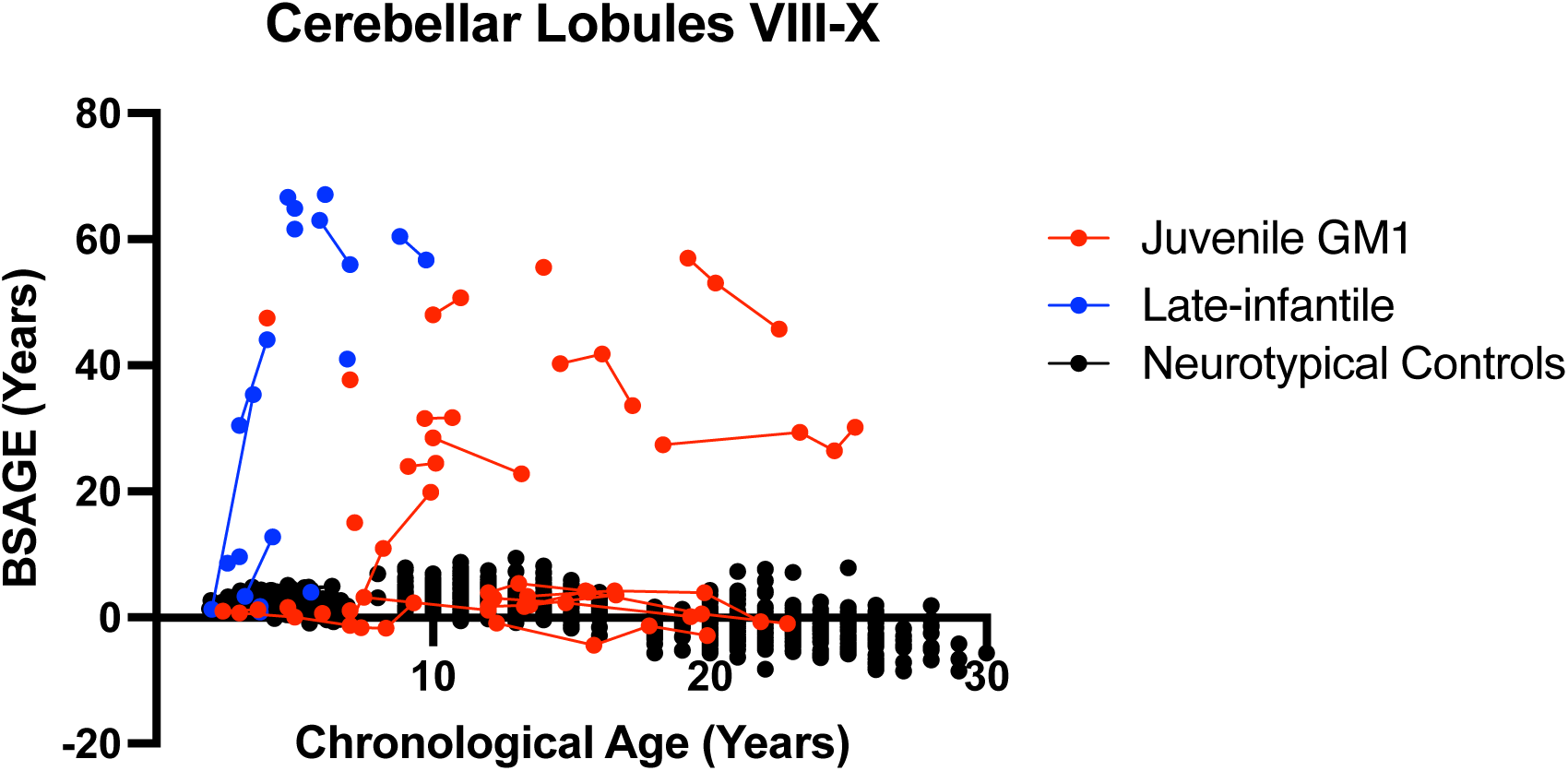
The longitudinal relationship between cerebellar lobules VIII-X BSAGE and chronological age. Late-infantile GM1 gangliosidosis patients are shown in blue, juvenile GM1 gangliosidosis patients are shown in red, and neurotypical controls participants are shown in black. Connecting lines indicate repeated scans on the same participant.

**Figure I7.**
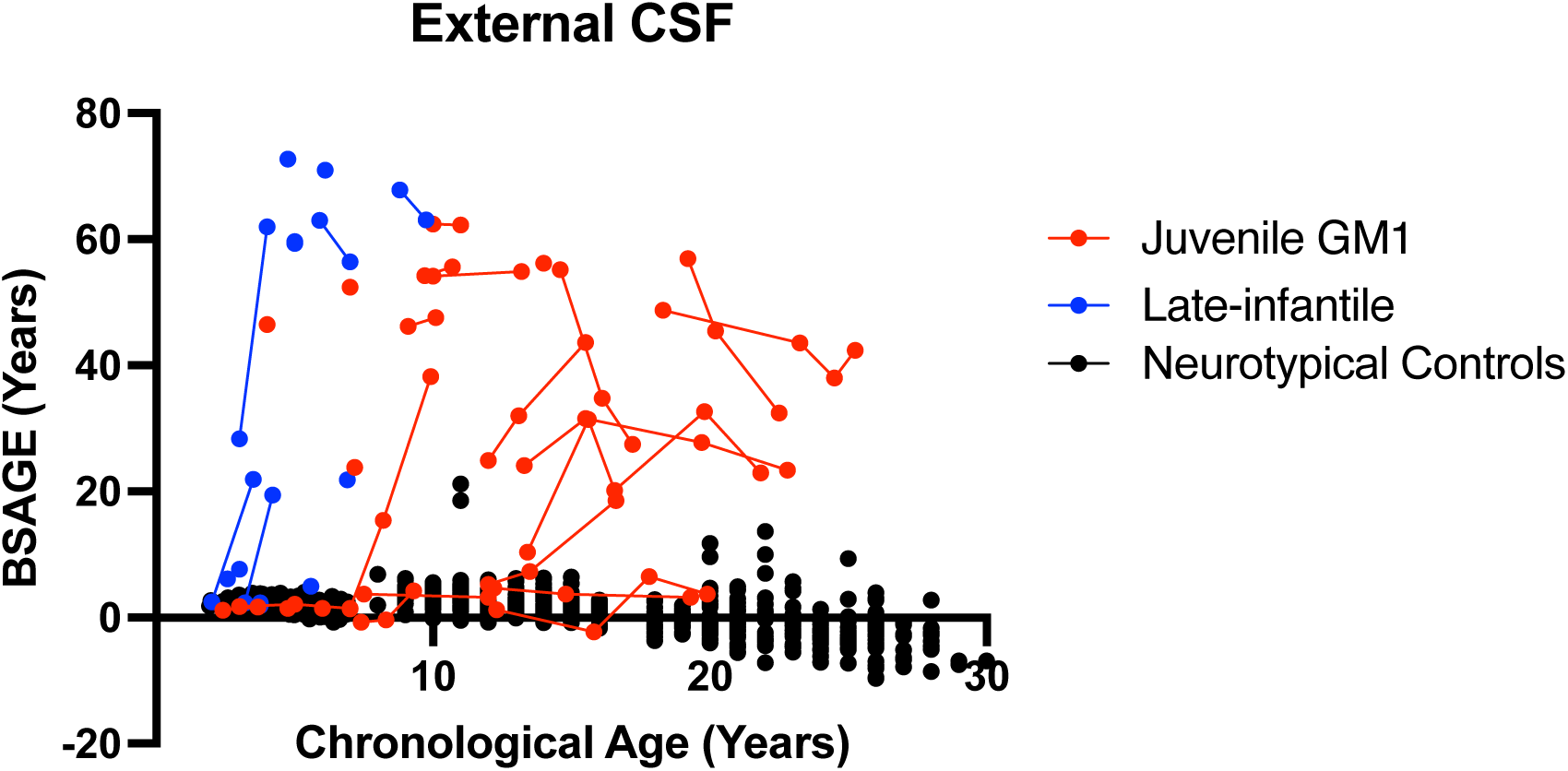
The longitudinal relationship between external cerebrospinal fluid (CSF) BSAGE and chronological age. Late-infantile GM1 gangliosidosis patients are shown in blue, juvenile GM1 gangliosidosis patients are shown in red, and neurotypical controls participants are shown in black. Connecting lines indicate repeated scans on the same participant.

**Figure I8.**
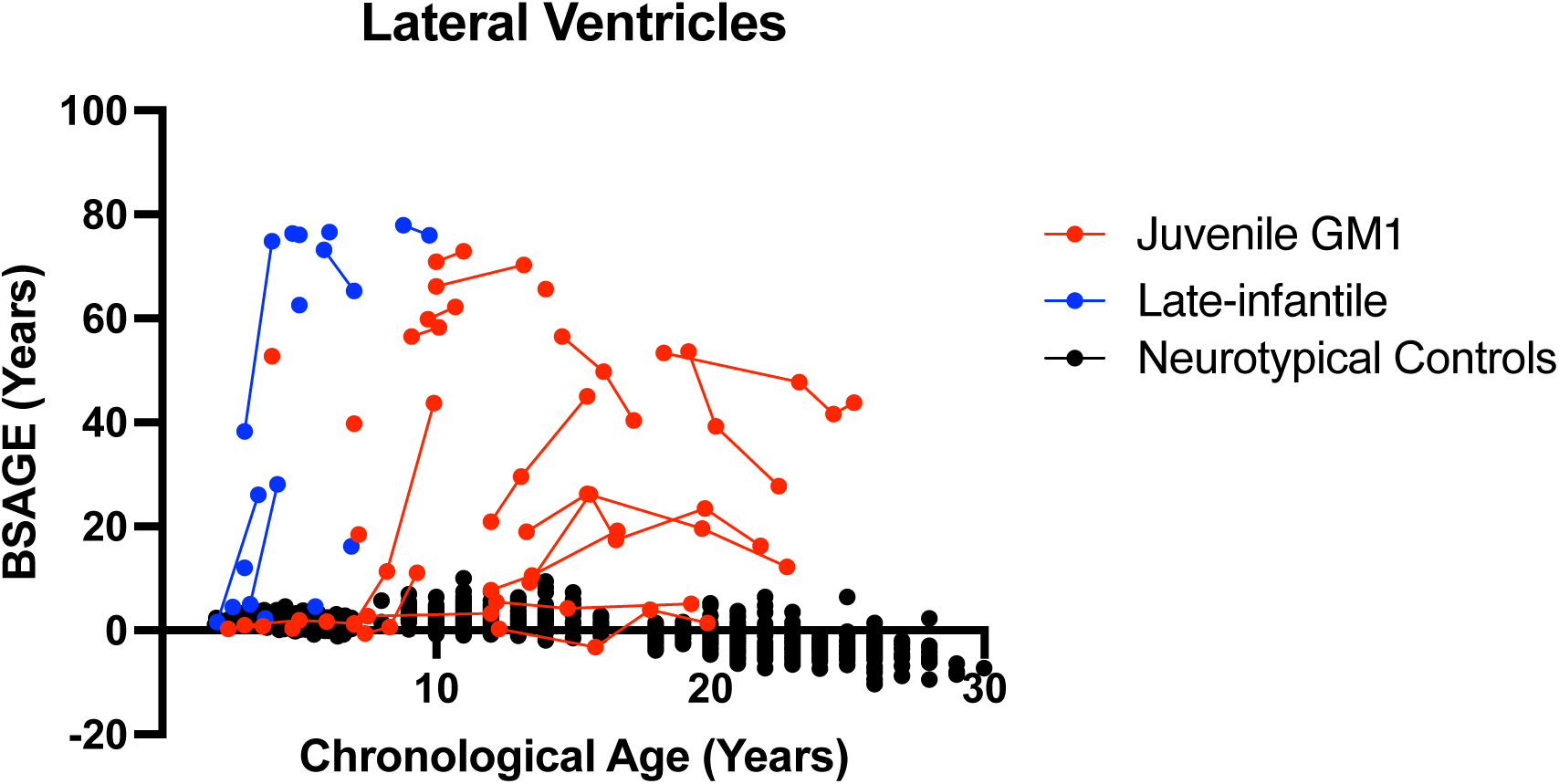
The longitudinal relationship between lateral ventricles BSAGE and chronological age. Late-infantile GM1 gangliosidosis patients are shown in blue, juvenile GM1 gangliosidosis patients are shown in red, and neurotypical controls participants are shown in black. Connecting lines indicate repeated scans on the same participant.

**Figure I9.**
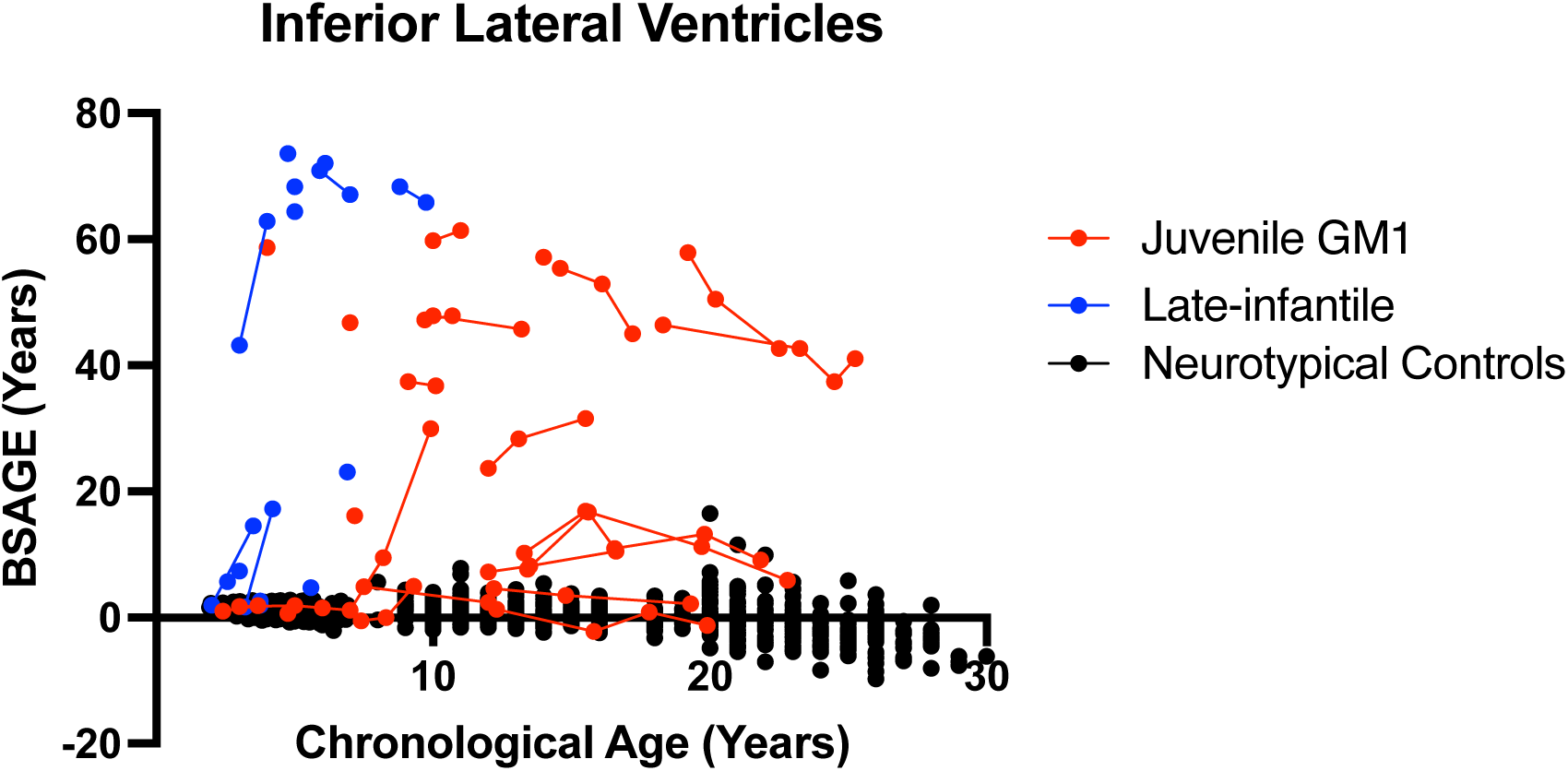
The longitudinal relationship between inferior lateral ventricles BSAGE and chronological age. Late-infantile GM1 gangliosidosis patients are shown in blue, juvenile GM1 gangliosidosis patients are shown in red, and neurotypical controls participants are shown in black. Connecting lines indicate repeated scans on the same participant.

**Figure I10.**
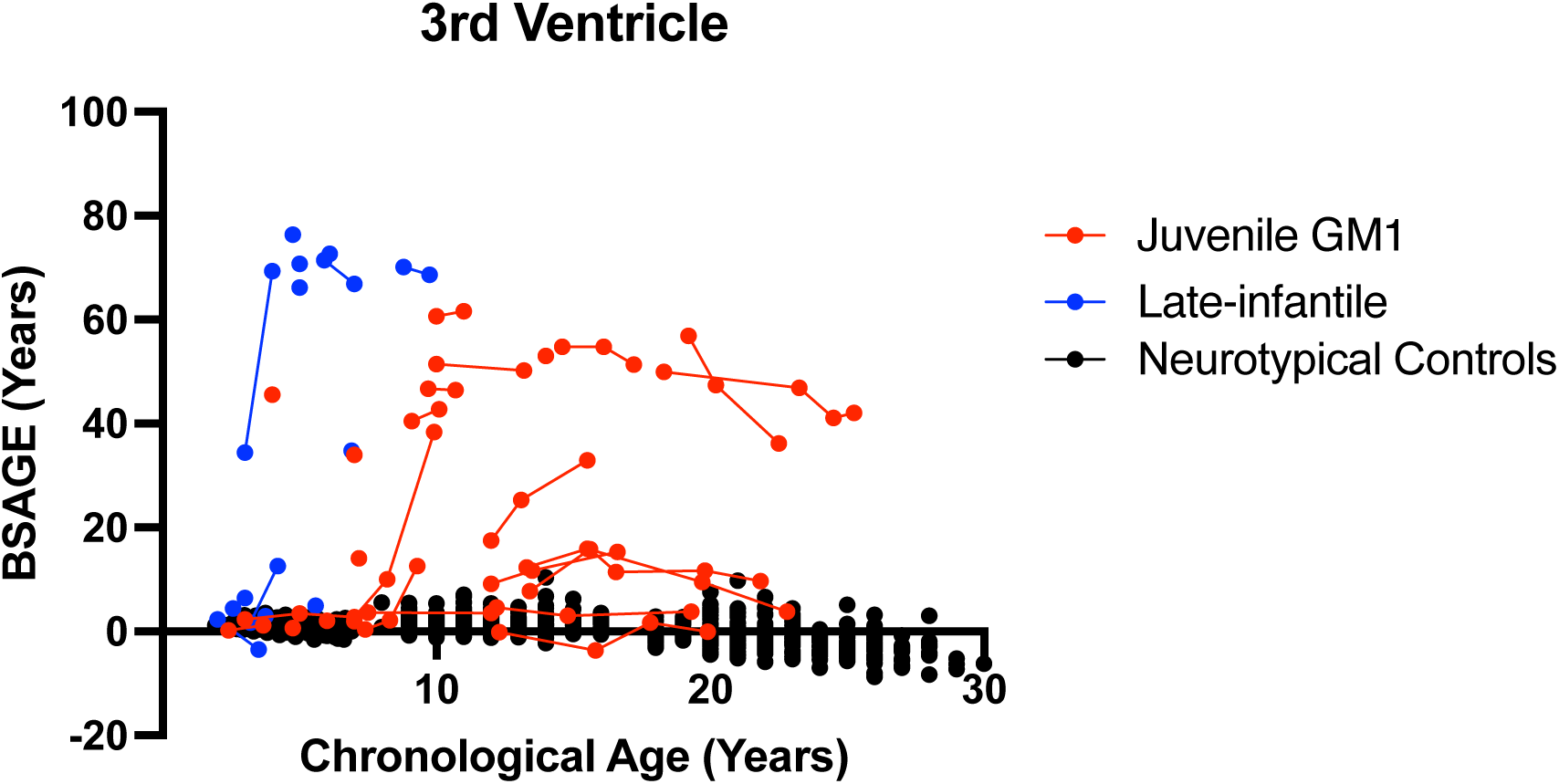
The longitudinal relationship between 3^rd^ ventricle BSAGE and chronological age. Late-infantile GM1 gangliosidosis patients are shown in blue, juvenile GM1 gangliosidosis patients are shown in red, and neurotypical controls participants are shown in black. Connecting lines indicate repeated scans on the same participant.

**Figure I11.**
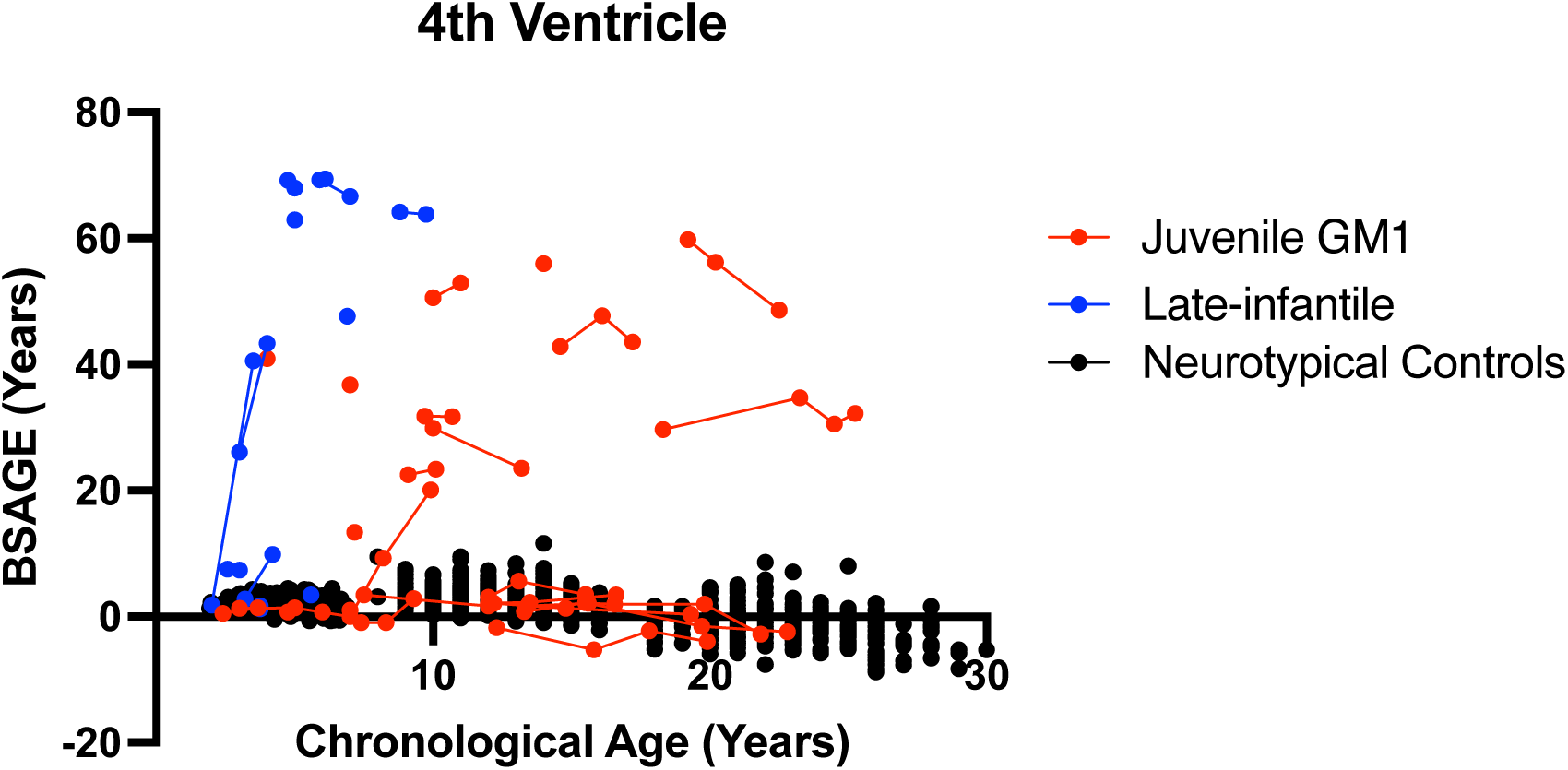
The longitudinal relationship between 4^th^ ventricle BSAGE and chronological age. Late-infantile GM1 gangliosidosis patients are shown in blue, juvenile GM1 gangliosidosis patients are shown in red, and neurotypical controls participants are shown in black. Connecting lines indicate repeated scans on the same participant.

**Figure I12.**
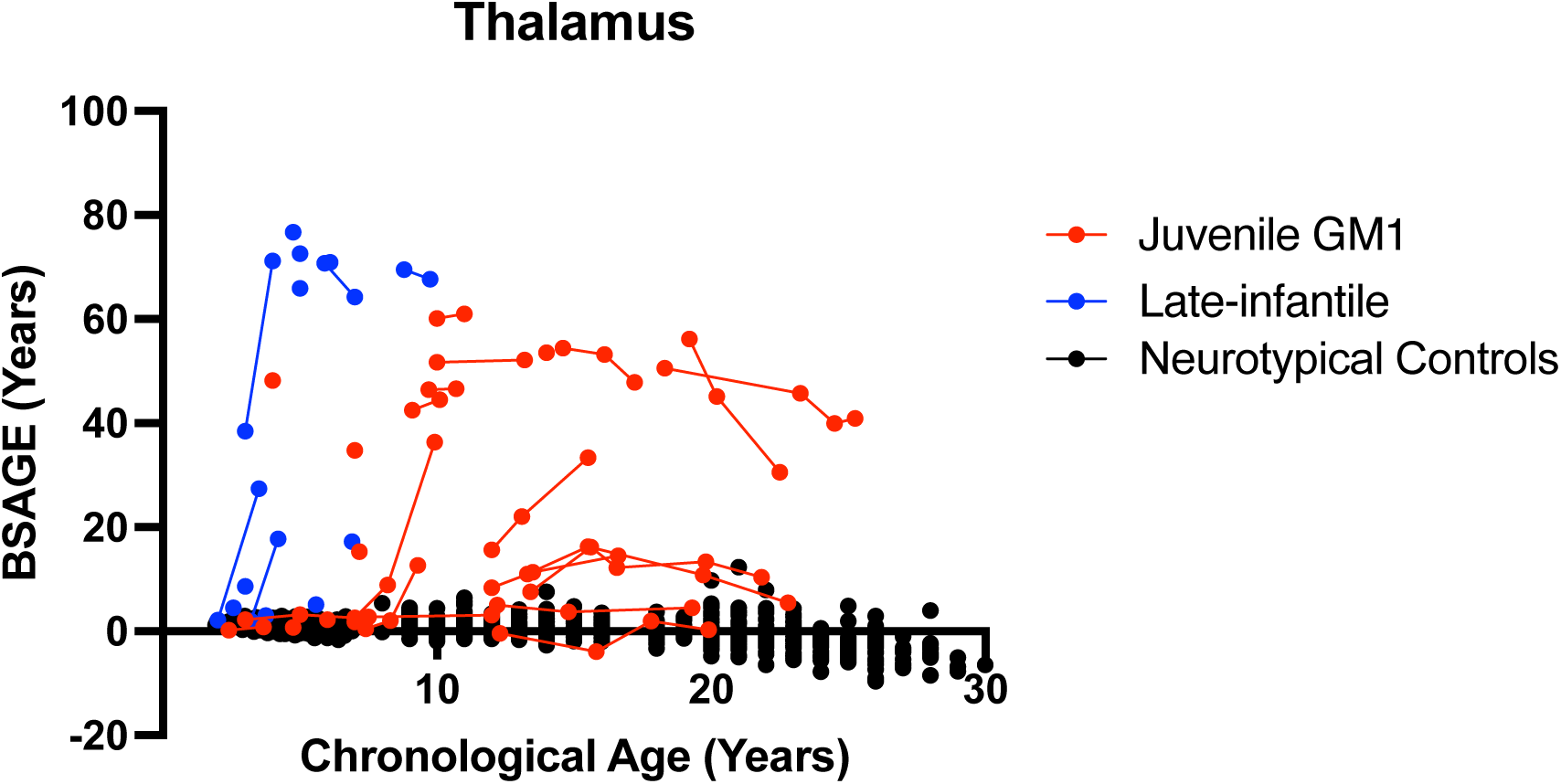
The longitudinal relationship between thalamus BSAGE and chronological age. Late-infantile GM1 gangliosidosis patients are shown in blue, juvenile GM1 gangliosidosis patients are shown in red, and neurotypical controls participants are shown in black. Connecting lines indicate repeated scans on the same participant.

**Figure I13.**
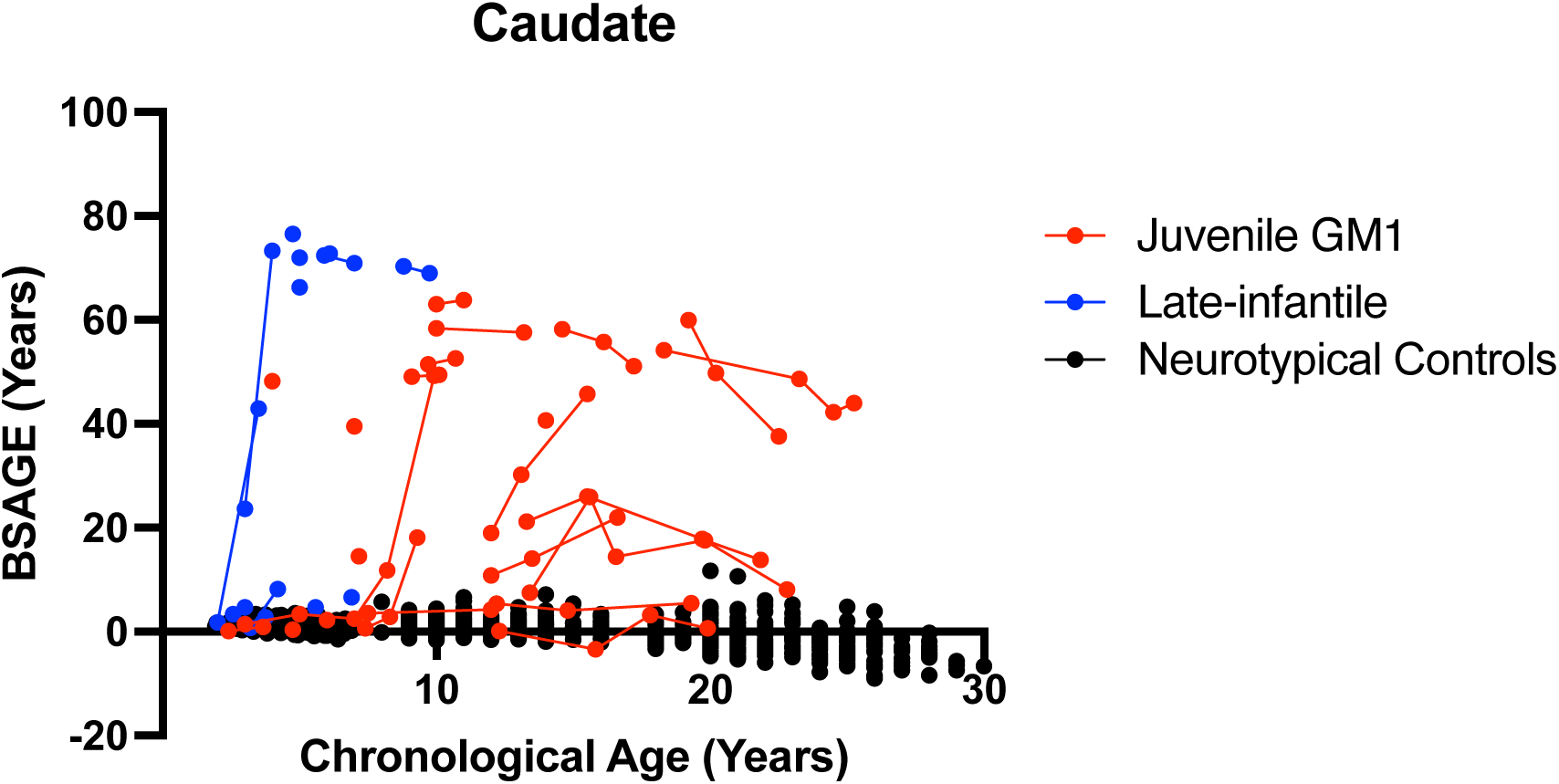
The longitudinal relationship between caudate nucleus BSAGE and chronological age. Late-infantile GM1 gangliosidosis patients are shown in blue, juvenile GM1 gangliosidosis patients are shown in red, and neurotypical controls participants are shown in black. Connecting lines indicate repeated scans on the same participant.

**Figure I14.**
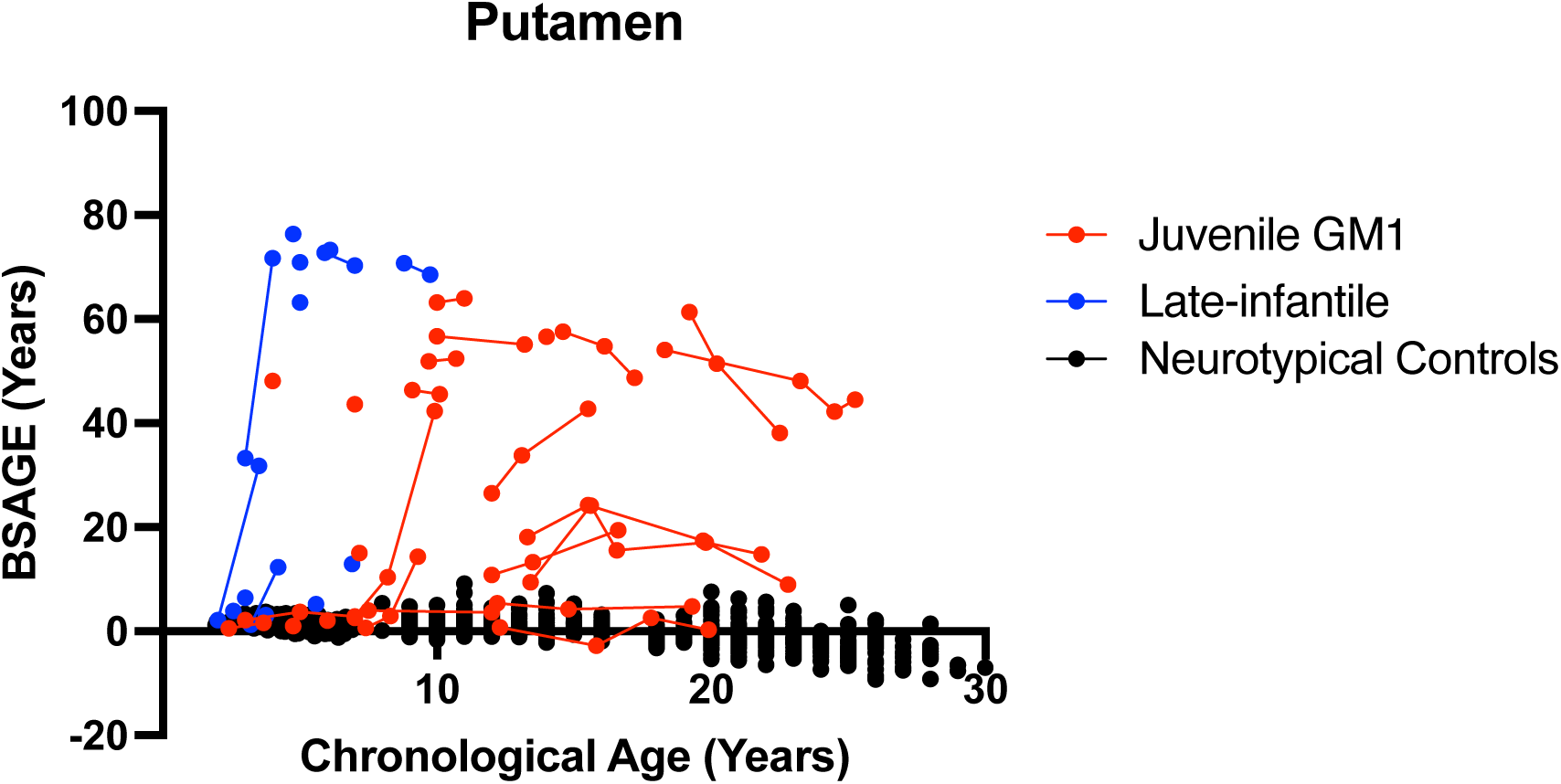
The longitudinal relationship between putamen BSAGE and chronological age. Late-infantile GM1 gangliosidosis patients are shown in blue, juvenile GM1 gangliosidosis patients are shown in red, and neurotypical controls participants are shown in black. Connecting lines indicate repeated scans on the same participant.

**Figure I15.**
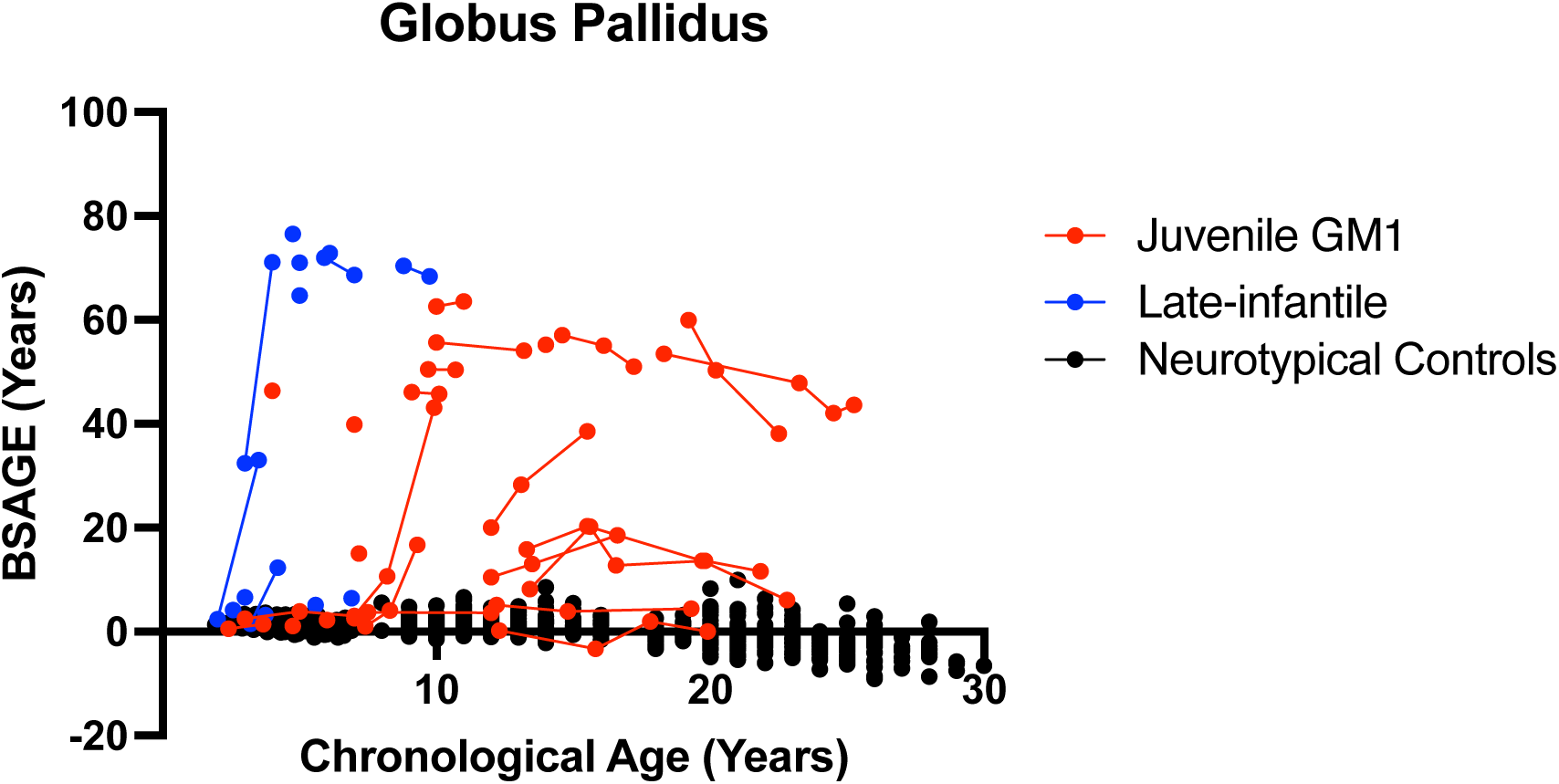
The longitudinal relationship between globus pallidus BSAGE and chronological age. Late-infantile GM1 gangliosidosis patients are shown in blue, juvenile GM1 gangliosidosis patients are shown in red, and neurotypical controls participants are shown in black. Connecting lines indicate repeated scans on the same participant.

**Figure I16.**
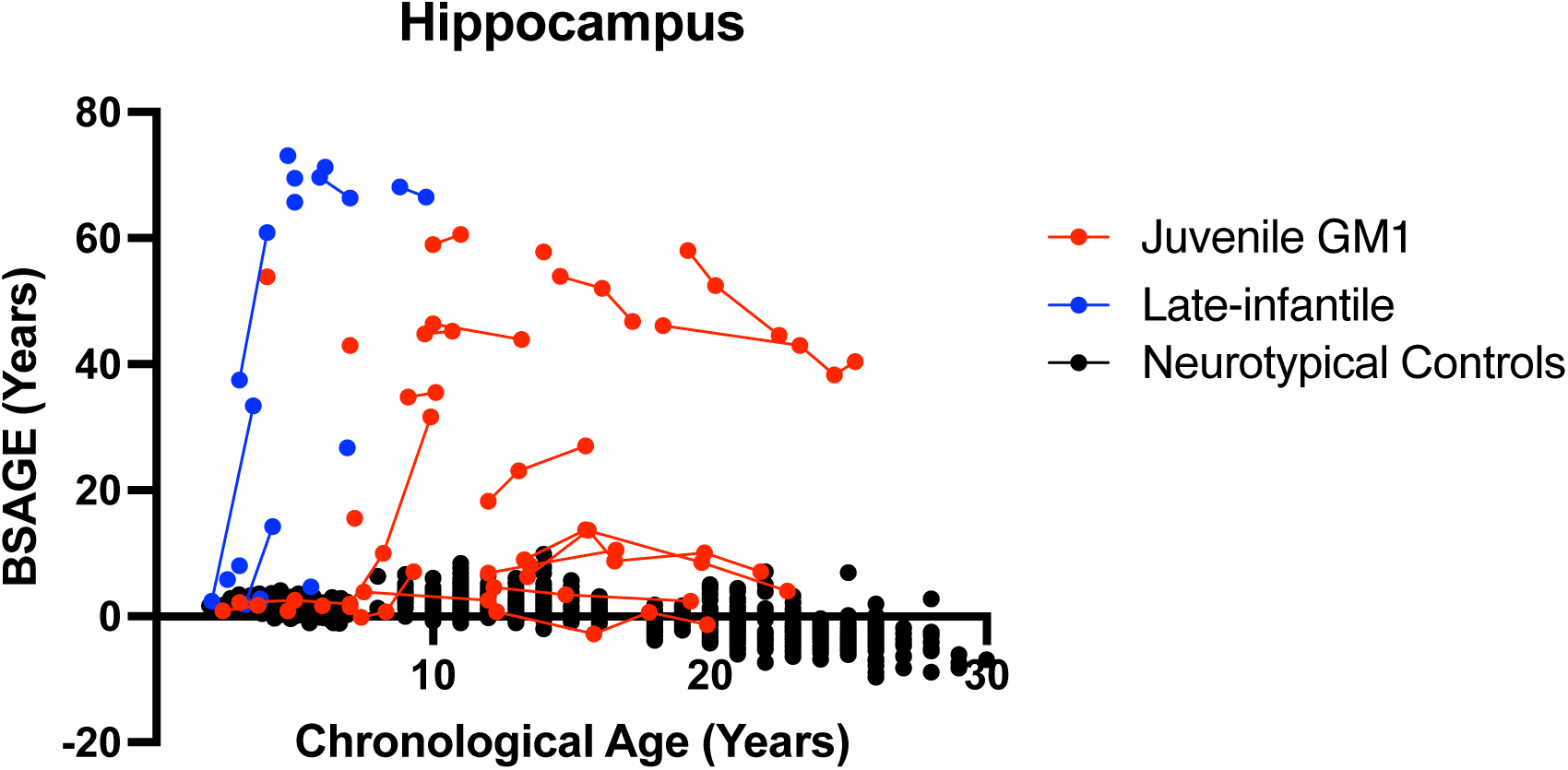
The longitudinal relationship between hippocampus BSAGE and chronological age. Late-infantile GM1 gangliosidosis patients are shown in blue, juvenile GM1 gangliosidosis patients are shown in red, and neurotypical controls participants are shown in black. Connecting lines indicate repeated scans on the same participant.

**Figure I17.**
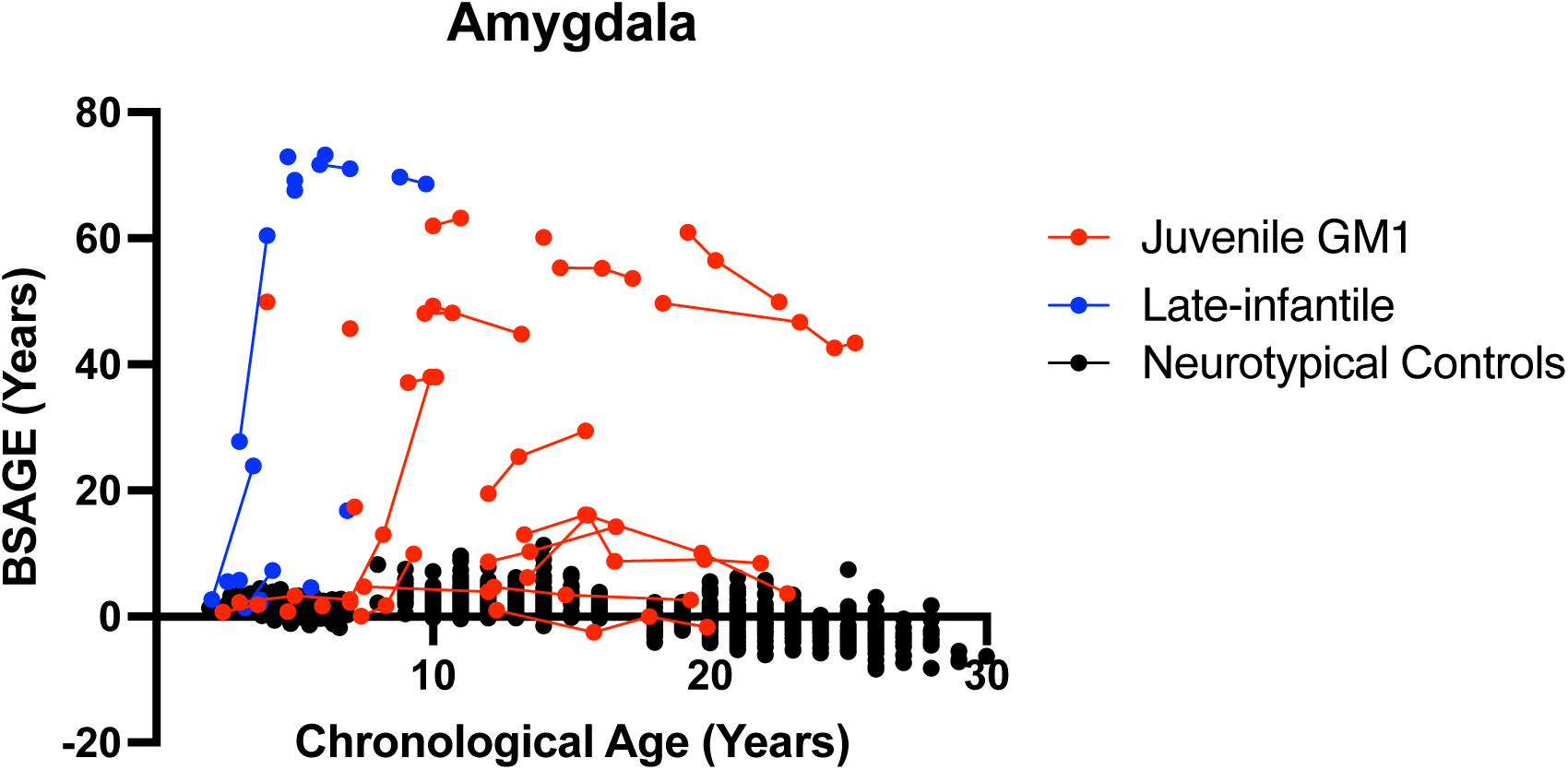
The longitudinal relationship between amygdala BSAGE and chronological age. Late-infantile GM1 gangliosidosis patients are shown in blue, juvenile GM1 gangliosidosis patients are shown in red, and neurotypical controls participants are shown in black. Connecting lines indicate repeated scans on the same participant.

**Figure I18.**
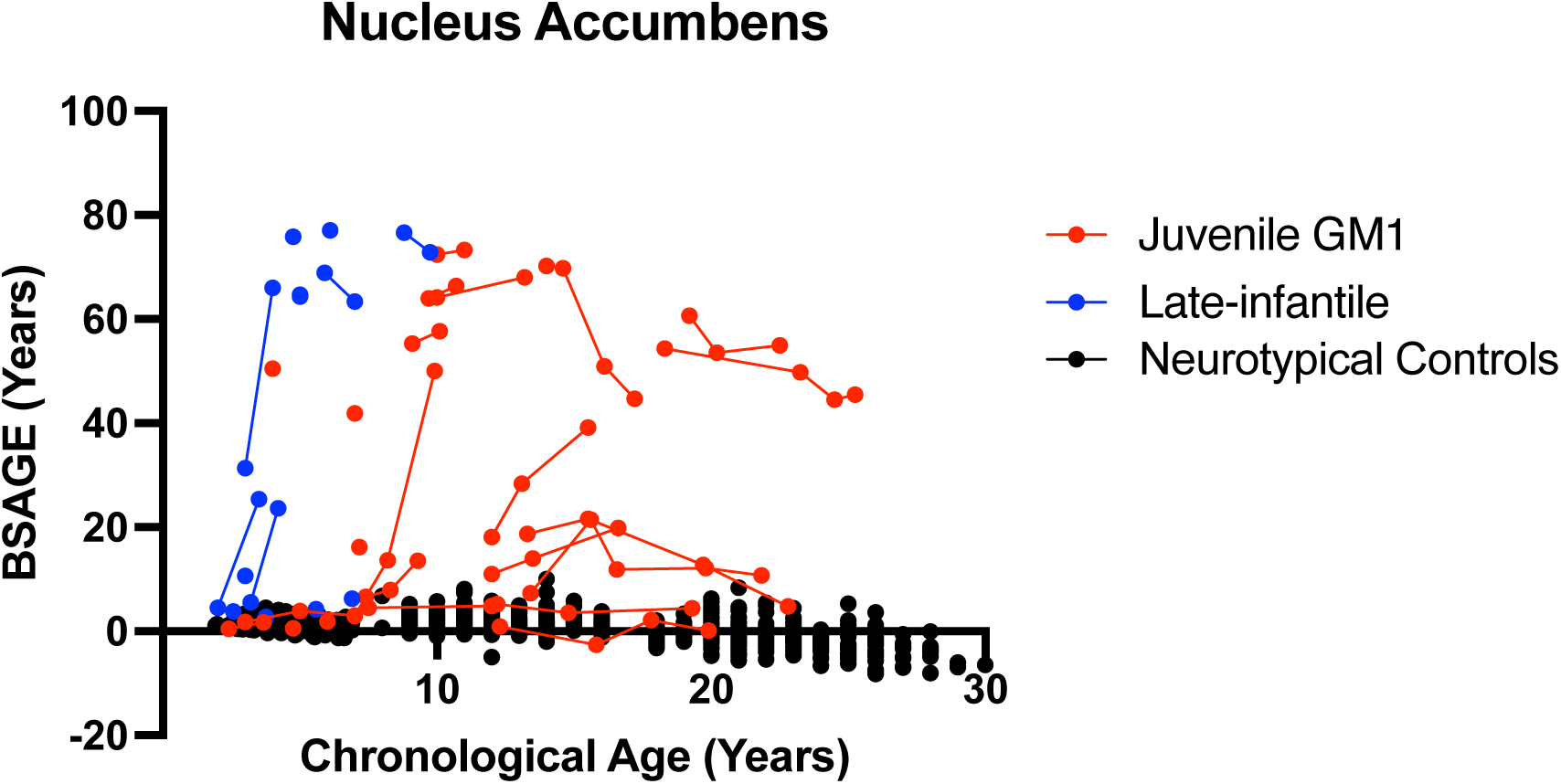
The longitudinal relationship between nucleus accumbens BSAGE and chronological age. Late-infantile GM1 gangliosidosis patients are shown in blue, juvenile GM1 gangliosidosis patients are shown in red, and neurotypical controls participants are shown in black. Connecting lines indicate repeated scans on the same participant.

**Figure I19.**
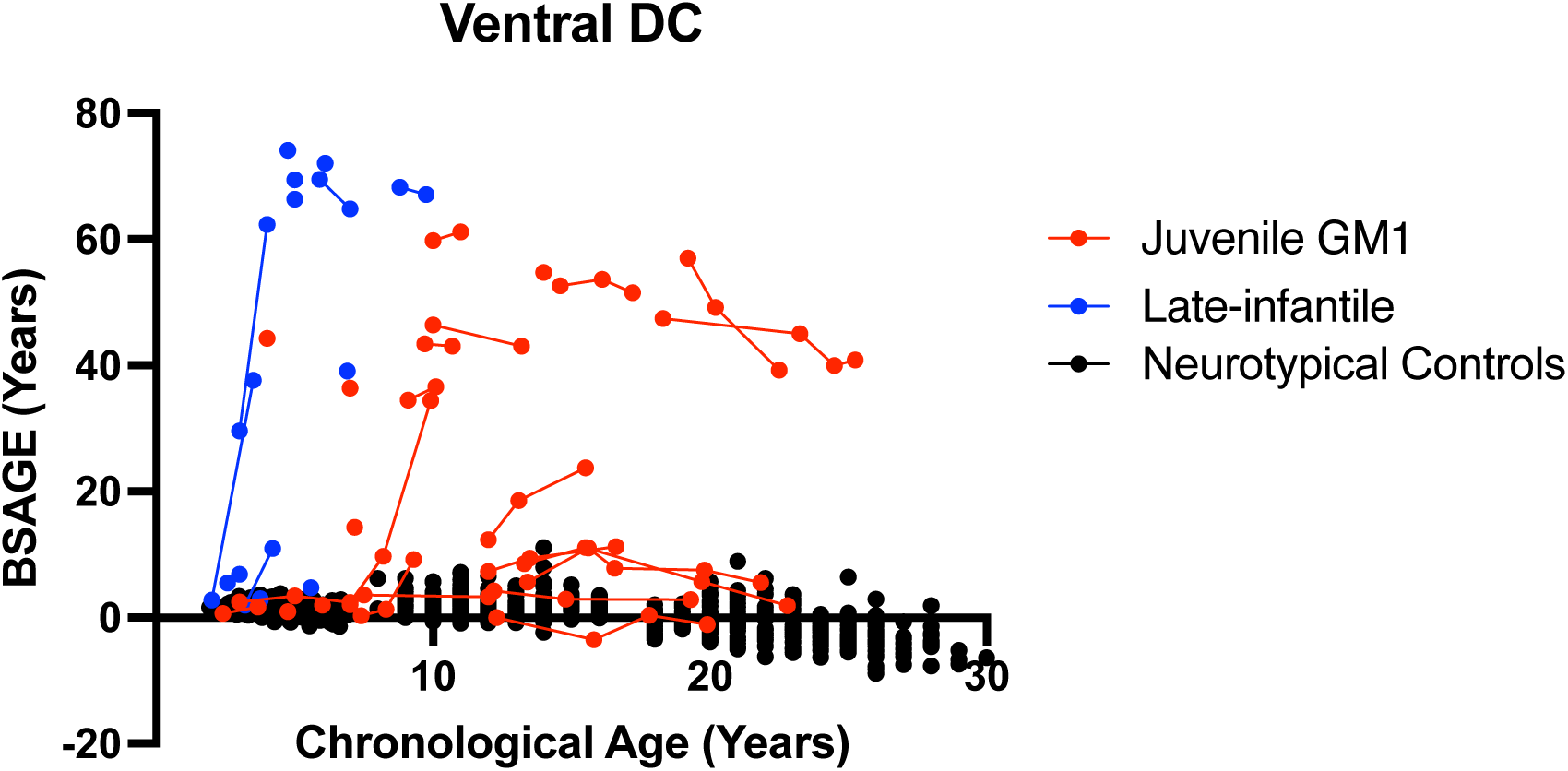
The longitudinal relationship between ventral diencephalon (DC) BSAGE and chronological age. Late-infantile GM1 gangliosidosis patients are shown in blue, juvenile GM1 gangliosidosis patients are shown in red, and neurotypical controls participants are shown in black. Connecting lines indicate repeated scans on the same participant.

**Figure I20.**
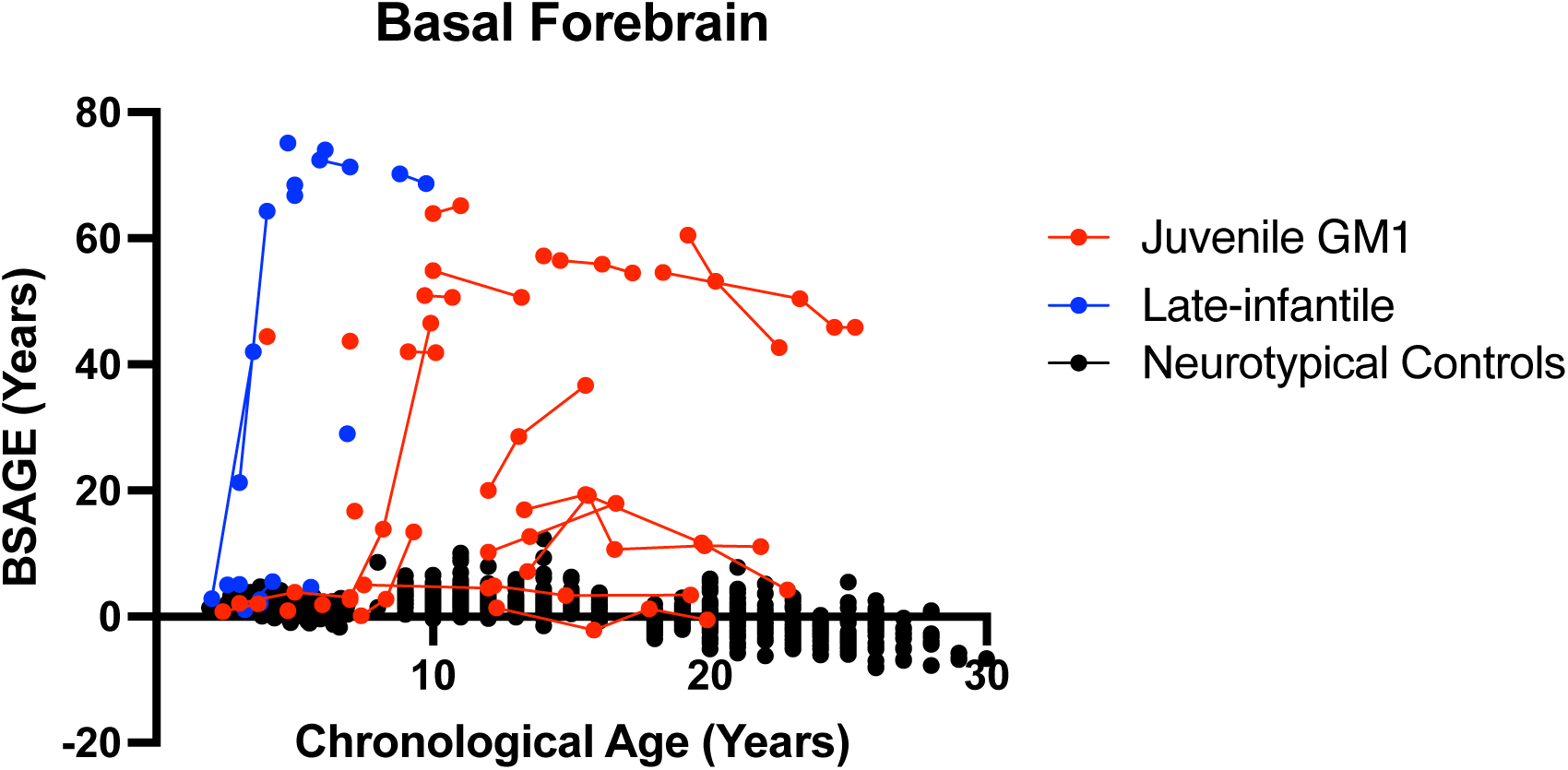
The longitudinal relationship between basal forebrain BSAGE and chronological age. Late-infantile GM1 gangliosidosis patients are shown in blue, juvenile GM1 gangliosidosis patients are shown in red, and neurotypical controls participants are shown in black. Connecting lines indicate repeated scans on the same participant.

**Figure I21.**
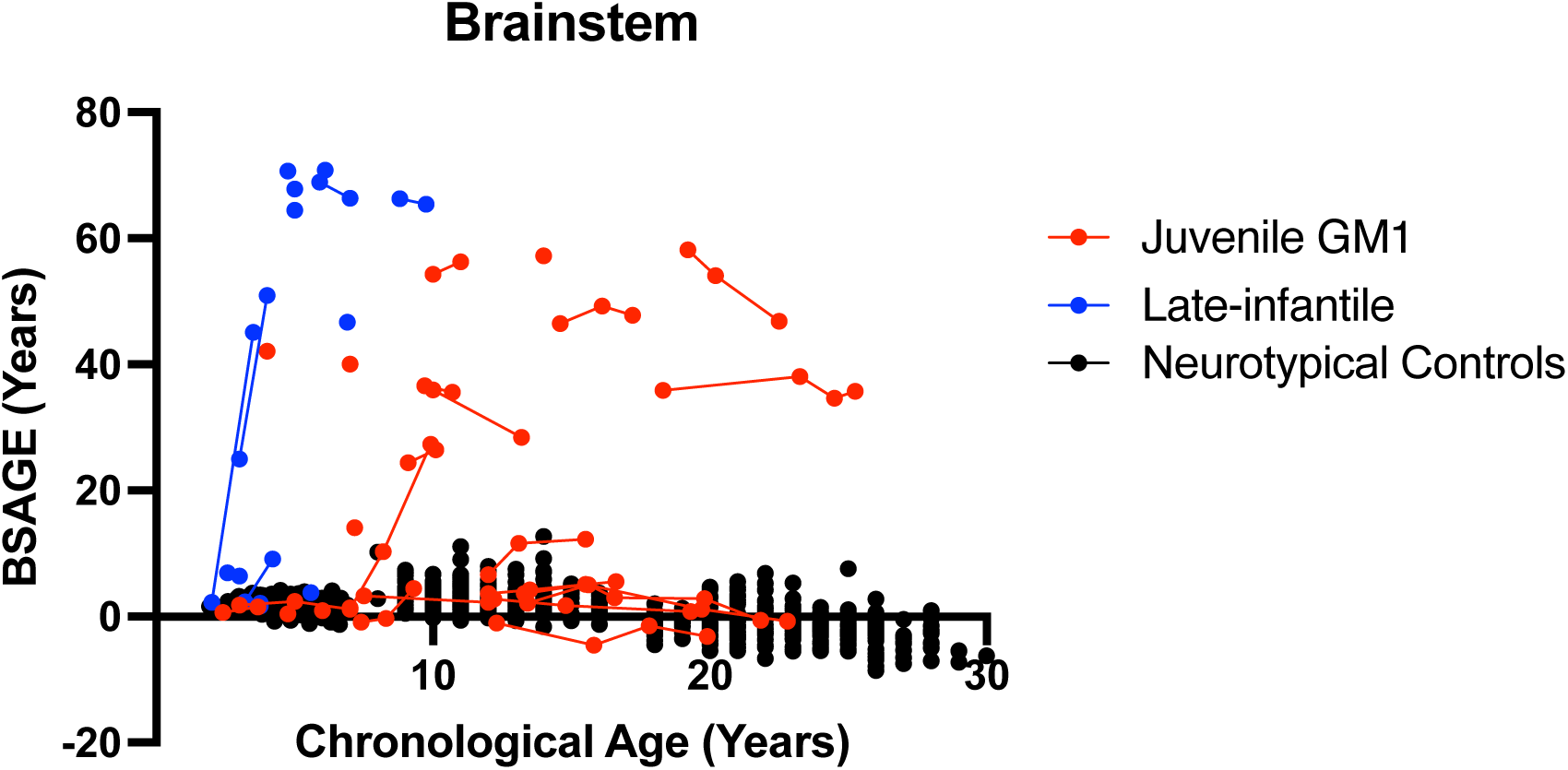
The longitudinal relationship between brainstem BSAGE and chronological age. Late-infantile GM1 gangliosidosis patients are shown in blue, juvenile GM1 gangliosidosis patients are shown in red, and neurotypical controls participants are shown in black. Connecting lines indicate repeated scans on the same participant.

### Supplement J: Cross-Sectional Brain structures age gap estimation (BSAGE)

**Figure J1.**
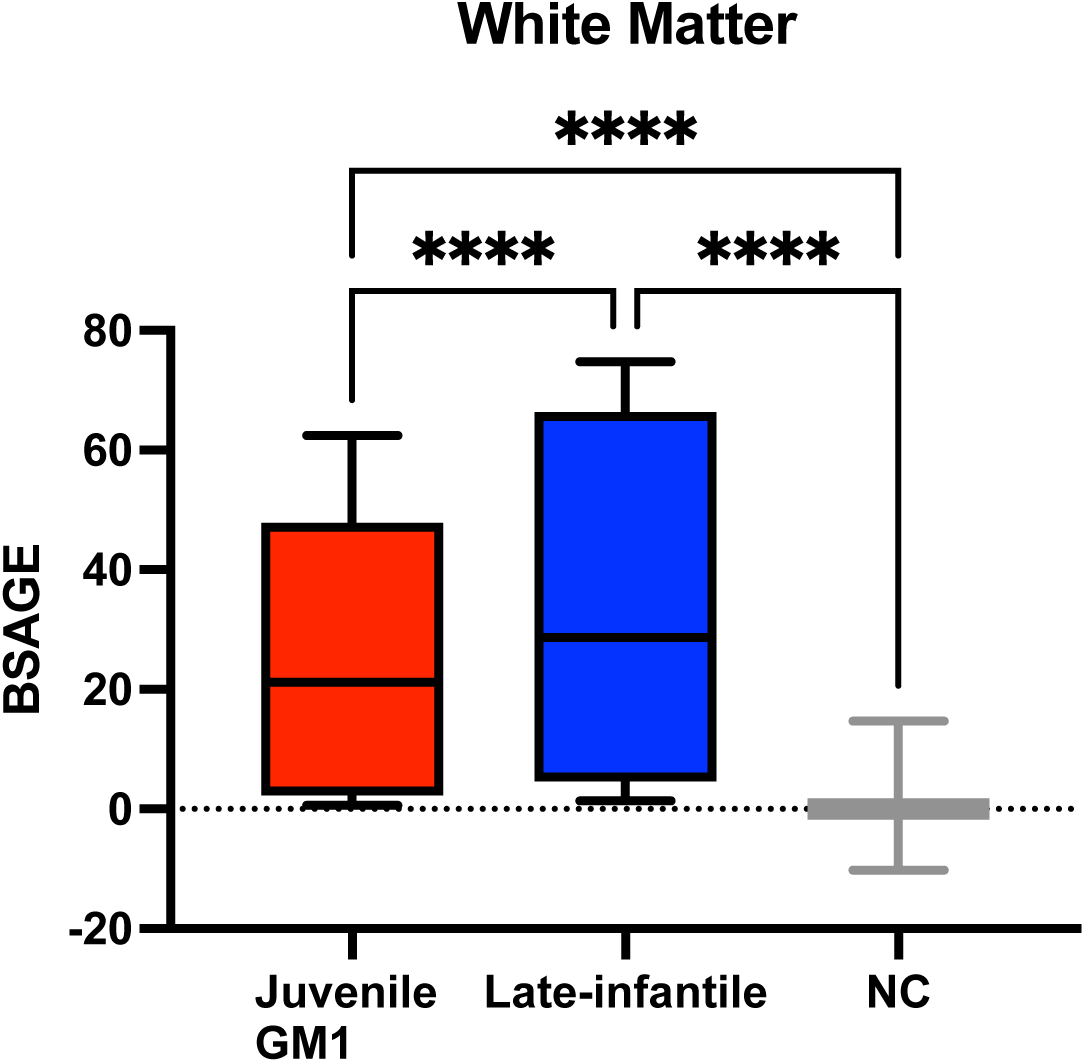
Comparison of White Matter BSAGE between Juvenile GM1 patients, late-infantile GM1 patients, and neurotypical controls. Late-infantile GM1 patients had an average BSAGE = 36.99, juvenile GM1 patients had an average BSAGE = 25.57, and NC participants had an average BSAGE = −0.21. An analysis of variance (ANOVA) showed statistically significant (F(2, 595) = 358.8, *p* < 0.0001) differences in BSAGE between the cohorts. A post-hoc Tukey test showed the late-infantile cohort had a statistically significantly higher BSAGE compared to both the neurotypical controls (*p* < 0.0001) and juvenile GM1 patients (*p* < 0.0001). The juvenile GM1 cohort also had a significantly higher BSAGE compared to the neurotypical controls (*p* < 0.0001).

**Figure J2.**
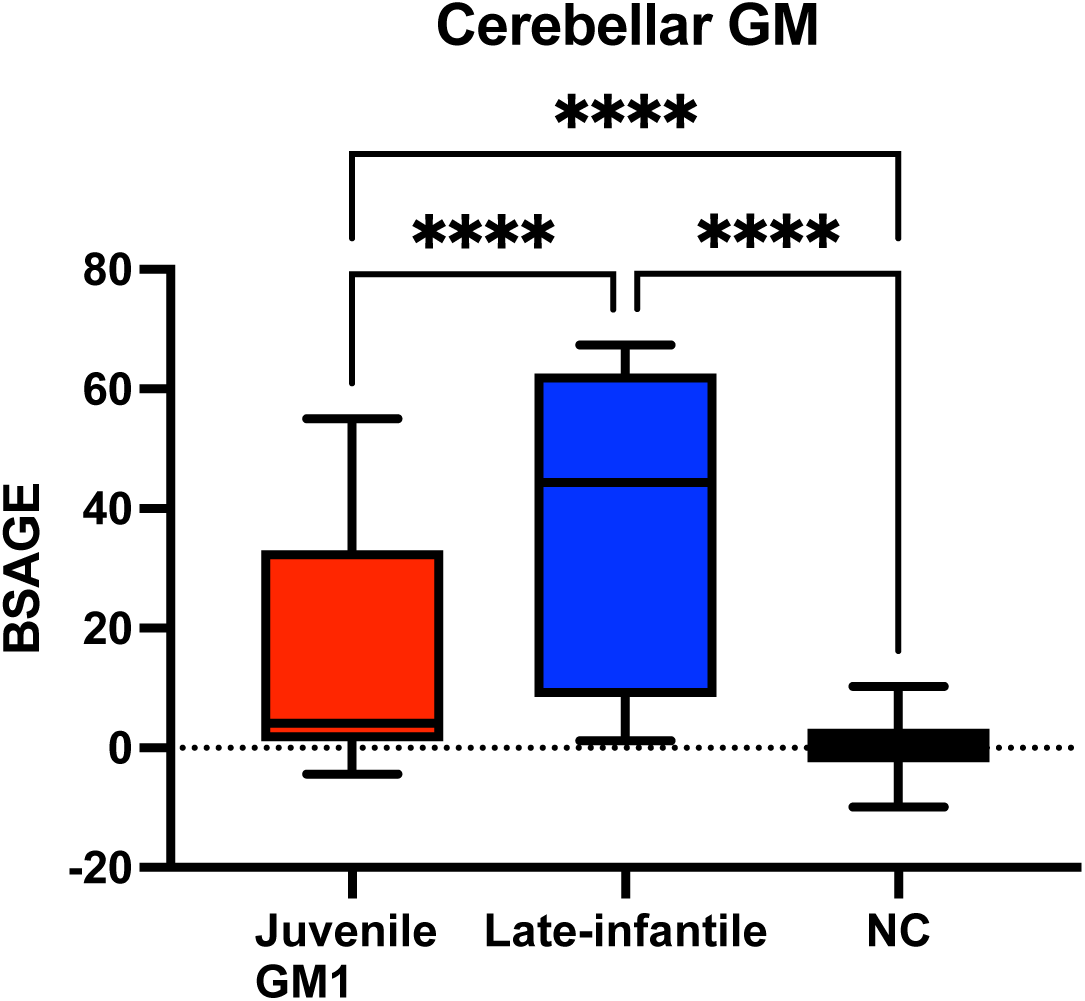
Comparison of Cerebellar Gray Matter BSAGE between Juvenile GM1 patients, late-infantile GM1 patients, and neurotypical controls. Late-infantile GM1 patients had an average BSAGE = 36.41, juvenile GM1 patients had an average BSAGE = 16.42, and NC participants had an average BSAGE = 0.49. An analysis of variance (ANOVA) showed statistically significant (F(2, 595) = 270.8, *p* < 0.0001) differences in BSAGE between the cohorts. A post-hoc Tukey test showed the late-infantile cohort had a statistically significantly higher BSAGE compared to both the neurotypical controls (*p* < 0.0001) and juvenile GM1 patients (*p* < 0.0001). The juvenile GM1 cohort also had a significantly higher BSAGE compared to the neurotypical controls (*p* < 0.0001).

**Figure J3.**
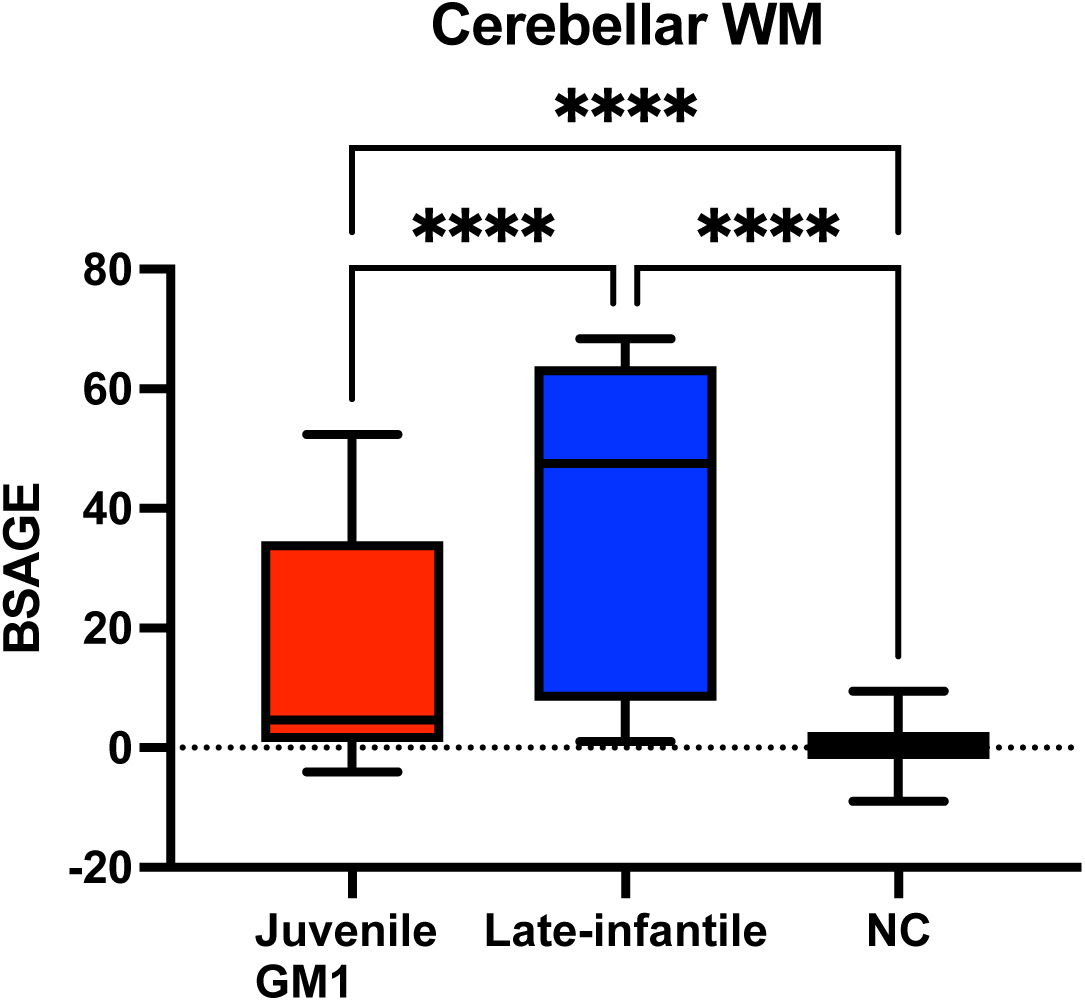
Comparison of Cerebellar White Matter BSAGE between Juvenile GM1 patients, late-infantile GM1 patients, and neurotypical controls. Late-infantile GM1 patients had an average BSAGE = 38.03, juvenile GM1 patients had an average BSAGE = 16.75, and NC participants had an average BSAGE = 0.36. An analysis of variance (ANOVA) showed statistically significant (F(2, 595) = 285.9, *p* < 0.0001) differences in BSAGE between the cohorts. A post-hoc Tukey test showed the late-infantile cohort had a statistically significantly higher BSAGE compared to both the neurotypical controls (*p* < 0.0001) and juvenile GM1 patients (*p* < 0.0001). The juvenile GM1 cohort also had a significantly higher BSAGE compared to the neurotypical controls (*p* < 0.0001).

**Figure J4.**
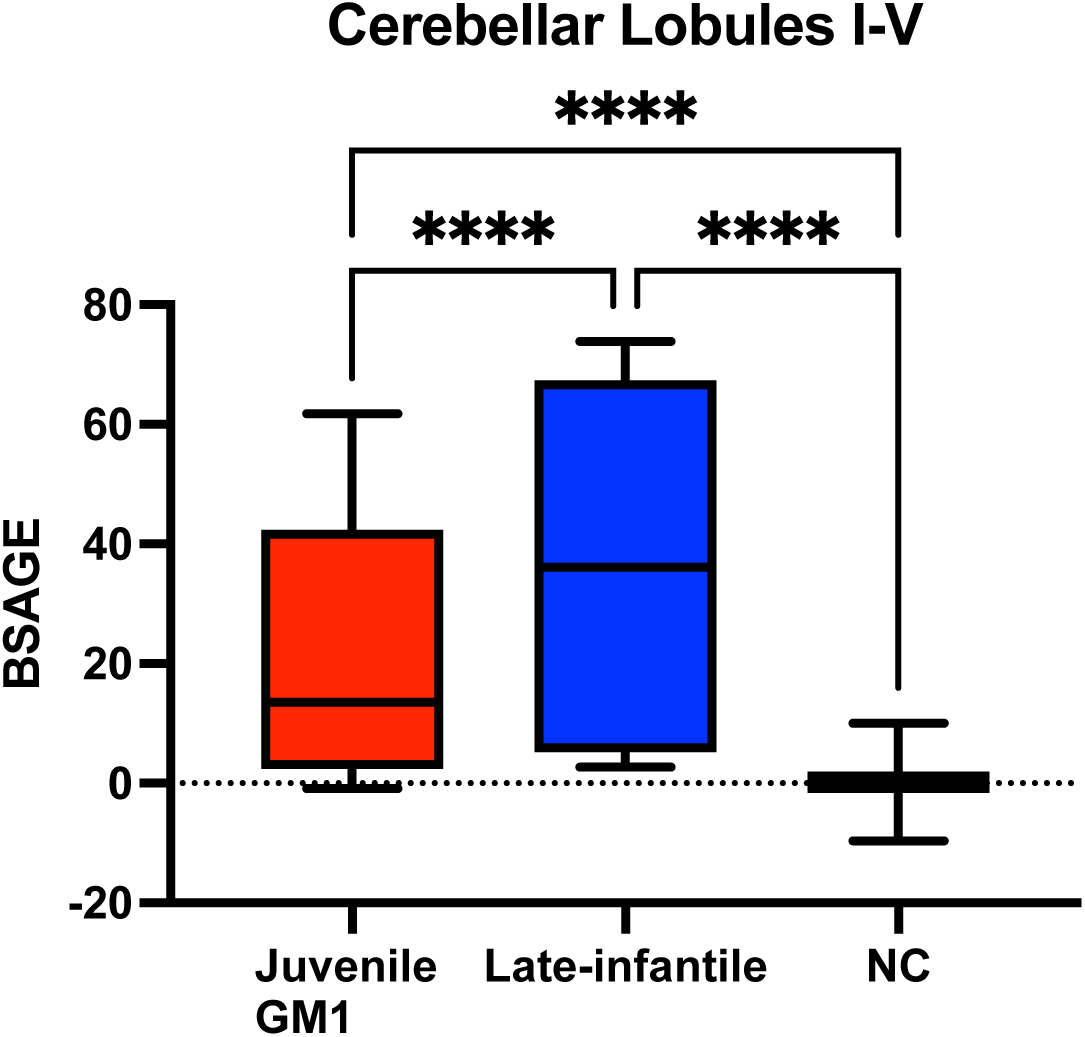
Comparison of Cerebellar Lobules I-V BSAGE between Juvenile GM1 patients, late-infantile GM1 patients, and neurotypical controls. Late-infantile GM1 patients had an average BSAGE = 38.82, juvenile GM1 patients had an average BSAGE = 22.79, and NC participants had an average BSAGE = 0.16. An analysis of variance (ANOVA) showed statistically significant (F(2, 595) = 585.6, *p* < 0.0001) differences in BSAGE between the cohorts. A post-hoc Tukey test showed the late-infantile cohort had a statistically significantly higher BSAGE compared to both the neurotypical controls (*p* < 0.0001) and juvenile GM1 patients (*p* < 0.0001). The juvenile GM1 cohort also had a significantly higher BSAGE compared to the neurotypical controls (*p* < 0.0001).

**Figure J5.**
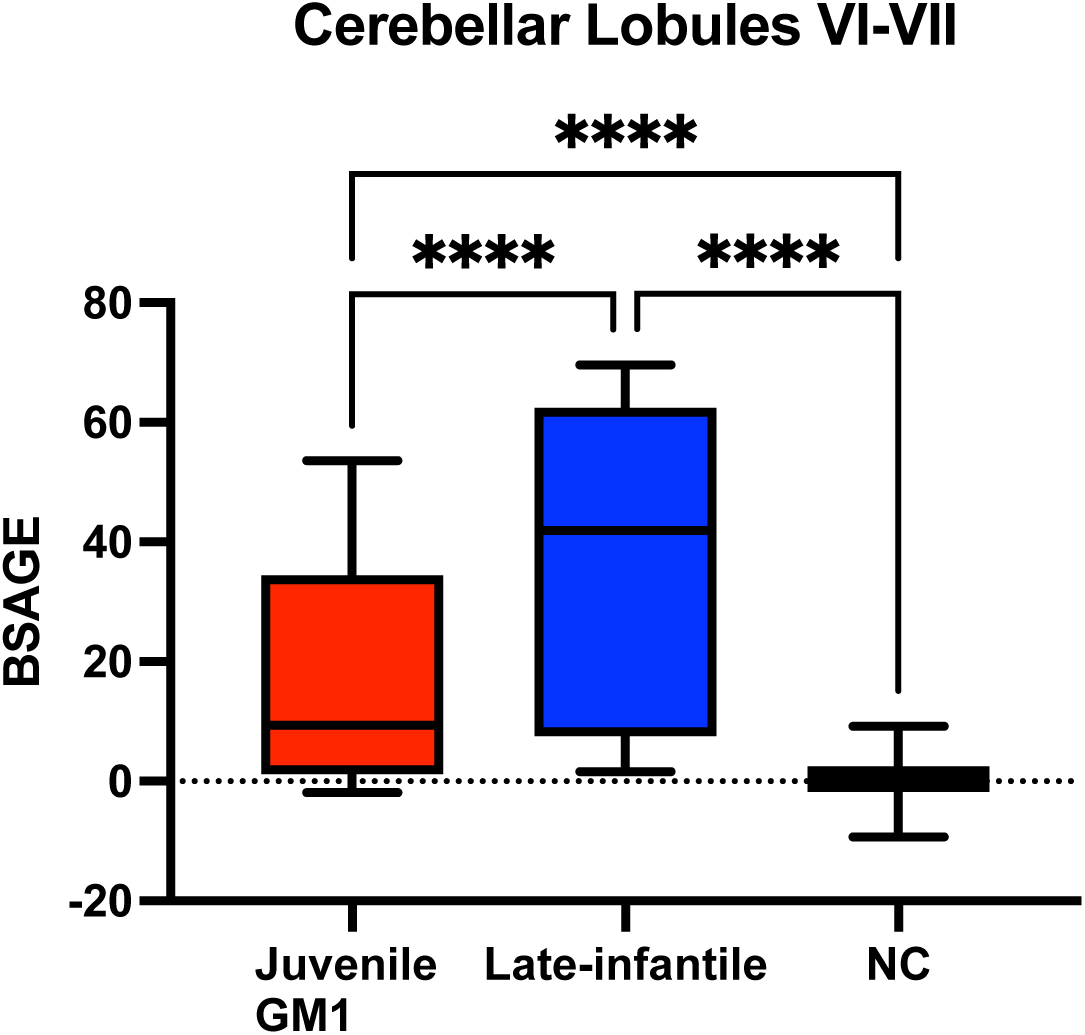
Comparison of Cerebellar Lobules VI-VII BSAGE between Juvenile GM1 patients, late-infantile GM1 patients, and neurotypical controls. Late-infantile GM1 patients had an average BSAGE = 36.97, juvenile GM1 patients had an average BSAGE = 17.58, and NC participants had an average BSAGE = 0.35. An analysis of variance (ANOVA) showed statistically significant (F(2, 595) = 358.3, *p* < 0.0001) differences in BSAGE between the cohorts. A post-hoc Tukey test showed the late-infantile cohort had a statistically significantly higher BSAGE compared to both the neurotypical controls (*p* < 0.0001) and juvenile GM1 patients (*p* < 0.0001). The juvenile GM1 cohort also had a significantly higher BSAGE compared to the neurotypical controls (*p* < 0.0001).

**Figure J6.**
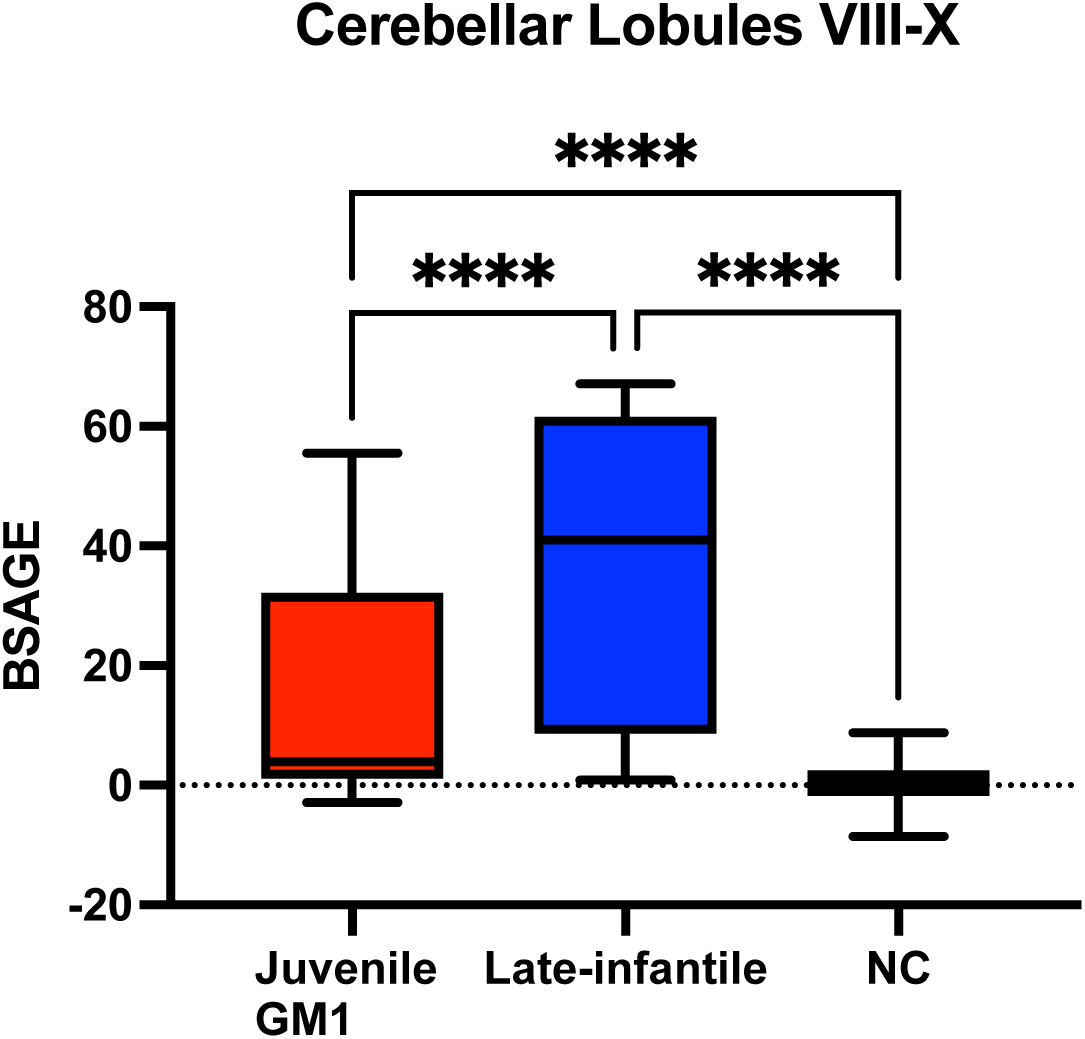
Comparison of Cerebellar Lobules VIII-X BSAGE between Juvenile GM1 patients, late-infantile GM1 patients, and neurotypical controls. Late-infantile GM1 patients had an average BSAGE = 35.40, juvenile GM1 patients had an average BSAGE = 16.50, and NC participants had an average BSAGE = 0.40. An analysis of variance (ANOVA) showed statistically significant (F(2, 595) = 284.5, *p* < 0.0001) differences in BSAGE between the cohorts. A post-hoc Tukey test showed the late-infantile cohort had a statistically significantly higher BSAGE compared to both the neurotypical controls (*p* < 0.0001) and juvenile GM1 patients (*p* < 0.0001). The juvenile GM1 cohort also had a significantly higher BSAGE compared to the neurotypical controls (*p* < 0.0001). Figure J7. Comparison of External Cerebrospinal Fluid (CSF) BSAGE between Juvenile GM1 patients, late-infantile GM1 patients, and neurotypical controls (NC). Late-infantile GM1 patients had an average BSAGE = 35.35, juvenile GM1 patients had an average BSAGE = 25.87, and NC participants had an average BSAGE = 0.46. An analysis of variance (ANOVA) showed statistically significant (F(2, 595) = 428.0, *p* < 0.0001) differences in BSAGE between the cohorts. A post-hoc Tukey test showed the late-infantile cohort had a statistically significantly higher BSAGE compared to both the neurotypical controls (*p* < 0.0001) and juvenile GM1 patients (*p* < 0.0001). The juvenile GM1 cohort also had a significantly higher BSAGE compared to the neurotypical controls (*p* < 0.0001).

**Figure J7.**
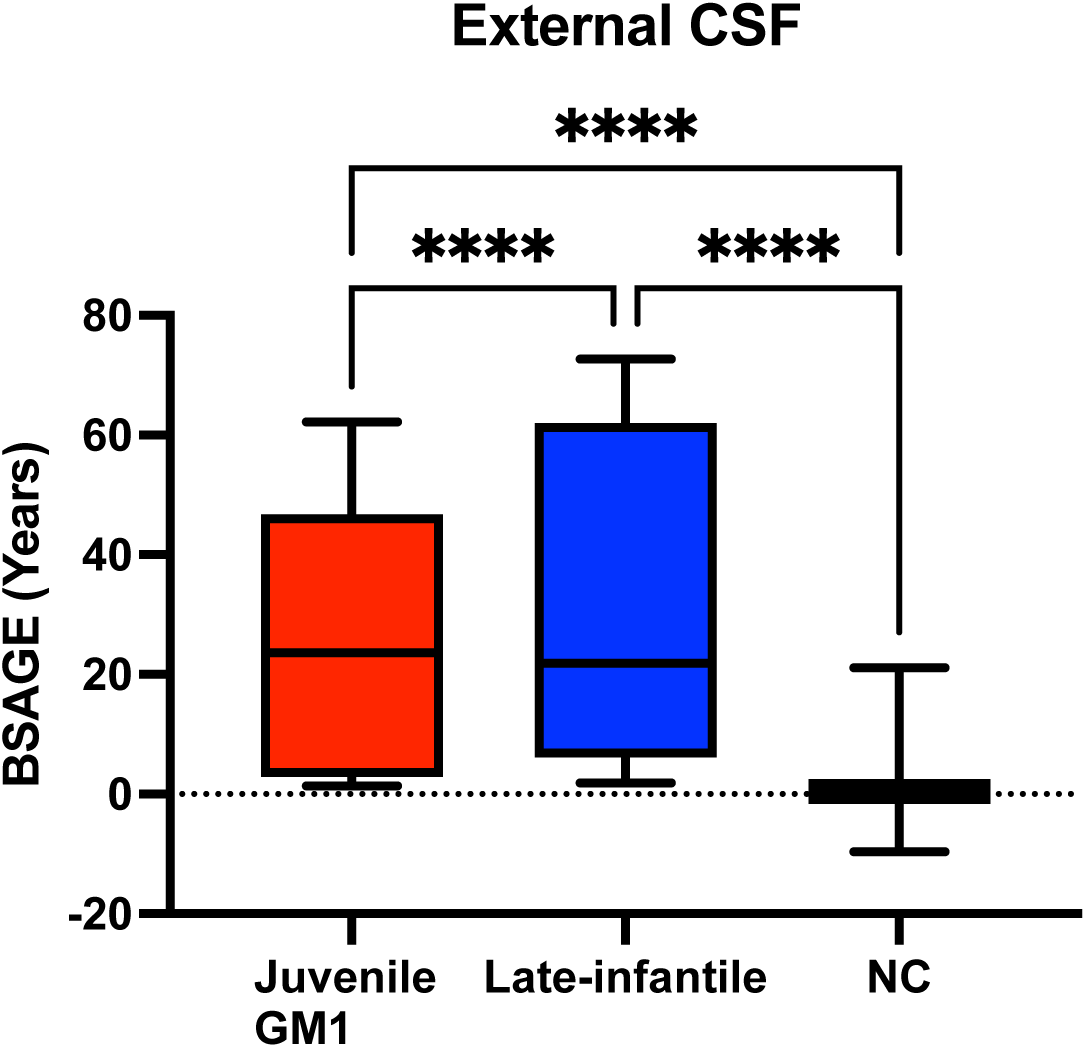
Comparison of External Cerebrospinal Fluid (CSF) BSAGE between Juvenile GM1 patients, late-infantile GM1 patients, and neurotypical controls (NC). Late-infantile GM1 patients had an average BSAGE = 35.35, juvenile GM1 patients had an average BSAGE = 25.87, and NC participants had an average BSAGE = 0.46. An analysis of variance (ANOVA) showed statistically significant (F(2, 595) = 428.0, *p* < 0.0001) differences in BSAGE between the cohorts. A post-hoc Tukey test showed the late-infantile cohort had a statistically significantly higher BSAGE compared to both the neurotypical controls (*p* < 0.0001) and juvenile GM1 patients (*p* < 0.0001). The juvenile GM1 cohort also had a significantly higher BSAGE compared to the neurotypical controls (*p* < 0.0001).

**Figure J8.**
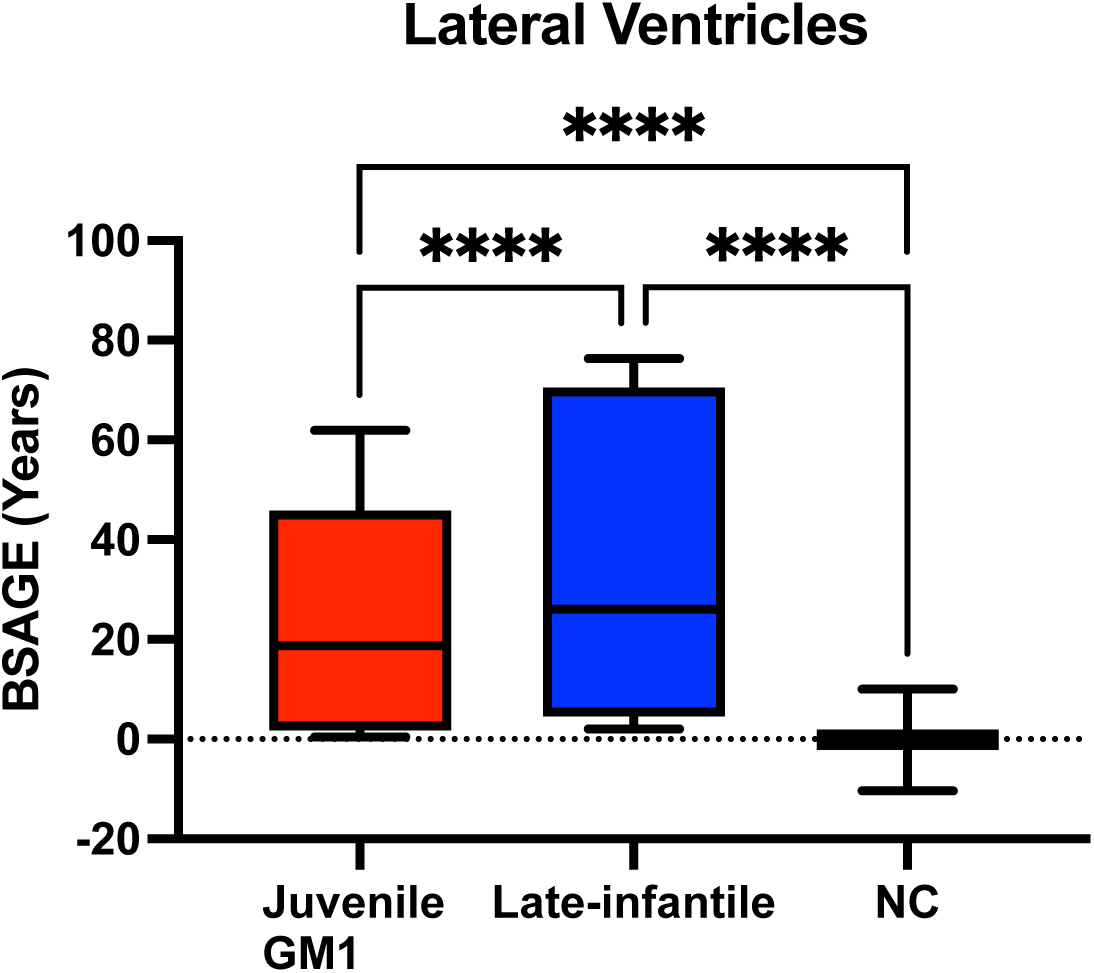
Comparison of lateral ventricle BSAGE between Juvenile GM1 patients, late-infantile GM1 patients, and neurotypical controls (NC). Late-infantile GM1 patients had an average BSAGE = 37.63, juvenile GM1 patients had an average BSAGE = 25.14, and NC participants had an average BSAGE = −0.06. An analysis of variance (ANOVA) showed statistically significant (F(2, 595) = 572.6, *p* < 0.0001) differences in BSAGE between the cohorts. A post- hoc Tukey test showed the late-infantile cohort had a statistically significantly higher BSAGE compared to both the neurotypical controls (*p* < 0.0001) and juvenile GM1 patients (*p* < 0.0001). The juvenile GM1 cohort also had a significantly higher BSAGE compared to the neurotypical controls (*p* < 0.0001).

**Figure J9.**
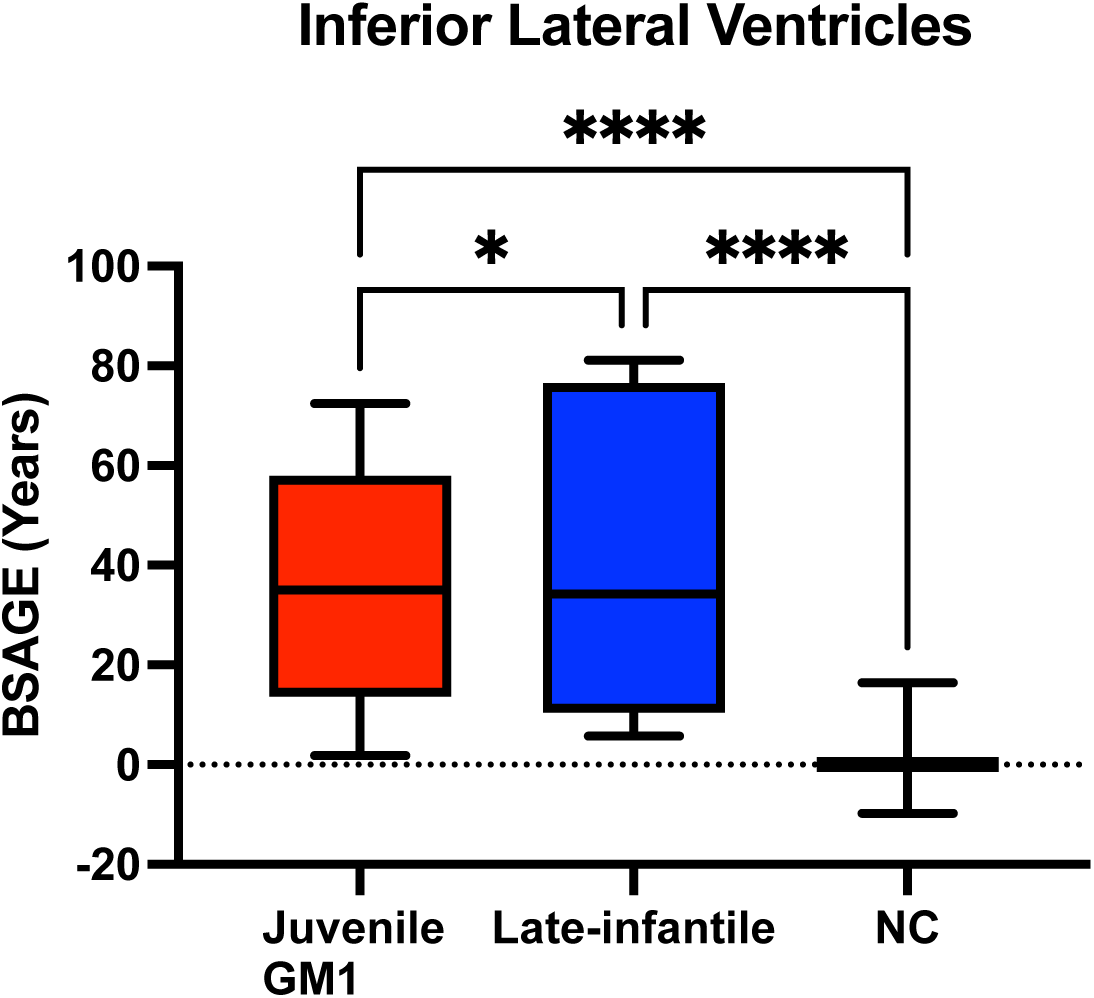
Comparison of the inferior lateral ventricle BSAGE between Juvenile GM1 patients, late-infantile GM1 patients, and neurotypical controls (NC). Late-infantile GM1 patients had an average BSAGE = 42.49, juvenile GM1 patients had an average BSAGE = 35.77, and NC participants had an average BSAGE = −0.09. An analysis of variance (ANOVA) showed statistically significant (F(2, 595) = 673.1, *p* < 0.0001) differences in BSAGE between the cohorts. A post-hoc Tukey test showed the late-infantile cohort had a statistically significantly higher BSAGE compared to both the neurotypical controls (*p* < 0.0001) and juvenile GM1 patients (*p* < 0.0001). The juvenile GM1 cohort also had a significantly higher BSAGE compared to the neurotypical controls (*p* < 0.0001).

**Figure J10.**
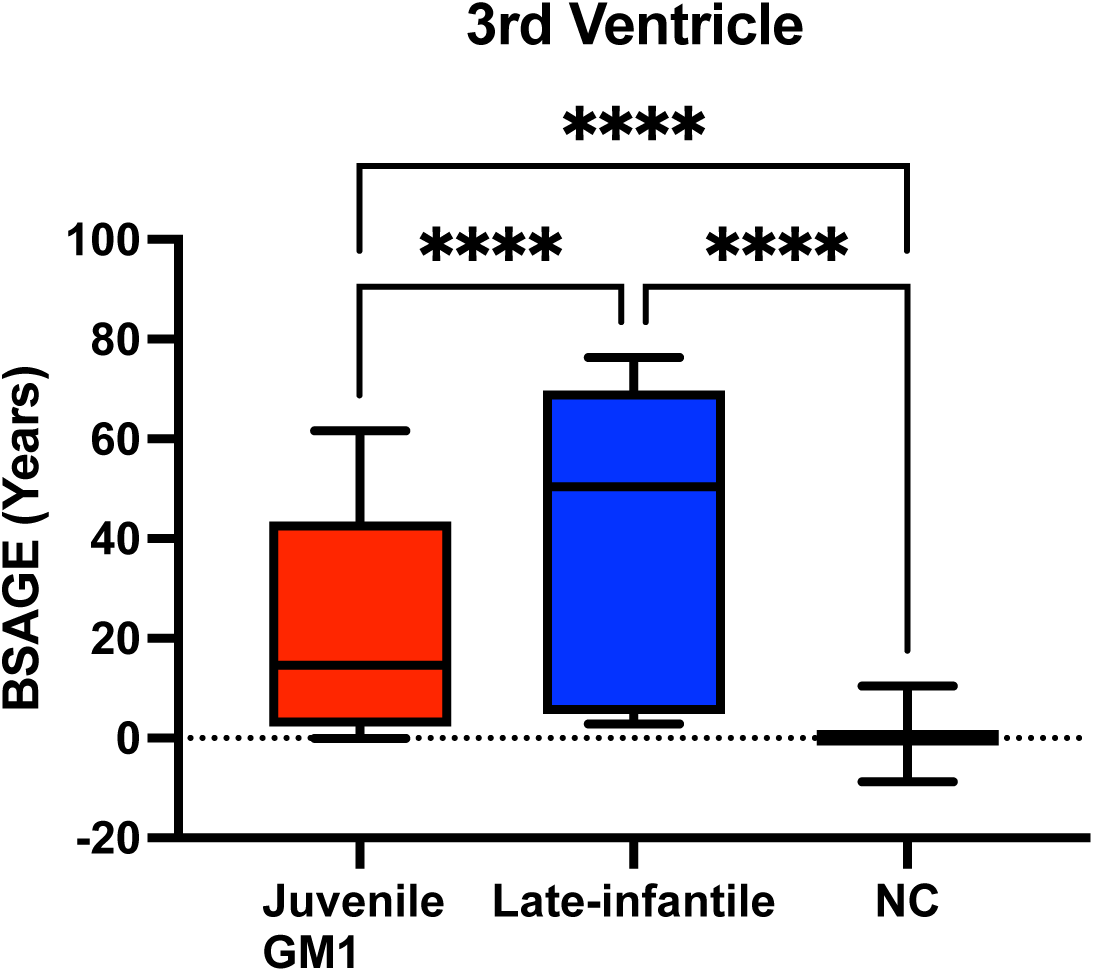
Comparison of the 3^rd^ ventricle BSAGE between Juvenile GM1 patients, late-infantile GM1 patients, and neurotypical controls (NC). Late-infantile GM1 patients had an average BSAGE = 40.00, juvenile GM1 patients had an average BSAGE = 23.34, and NC participants had an average BSAGE = 0.04. An analysis of variance (ANOVA) showed statistically significant (F(2,594) = 650.1, *p* < 0.0001) differences in BSAGE between the cohorts. A post-hoc Tukey test showed the late-infantile cohort had a statistically significantly higher BSAGE compared to both the neurotypical controls (*p* < 0.0001) and juvenile GM1 patients (*p* < 0.0001). The juvenile GM1 cohort also had a significantly higher BSAGE compared to the neurotypical controls (*p* < 0.0001).

**Figure J11.**
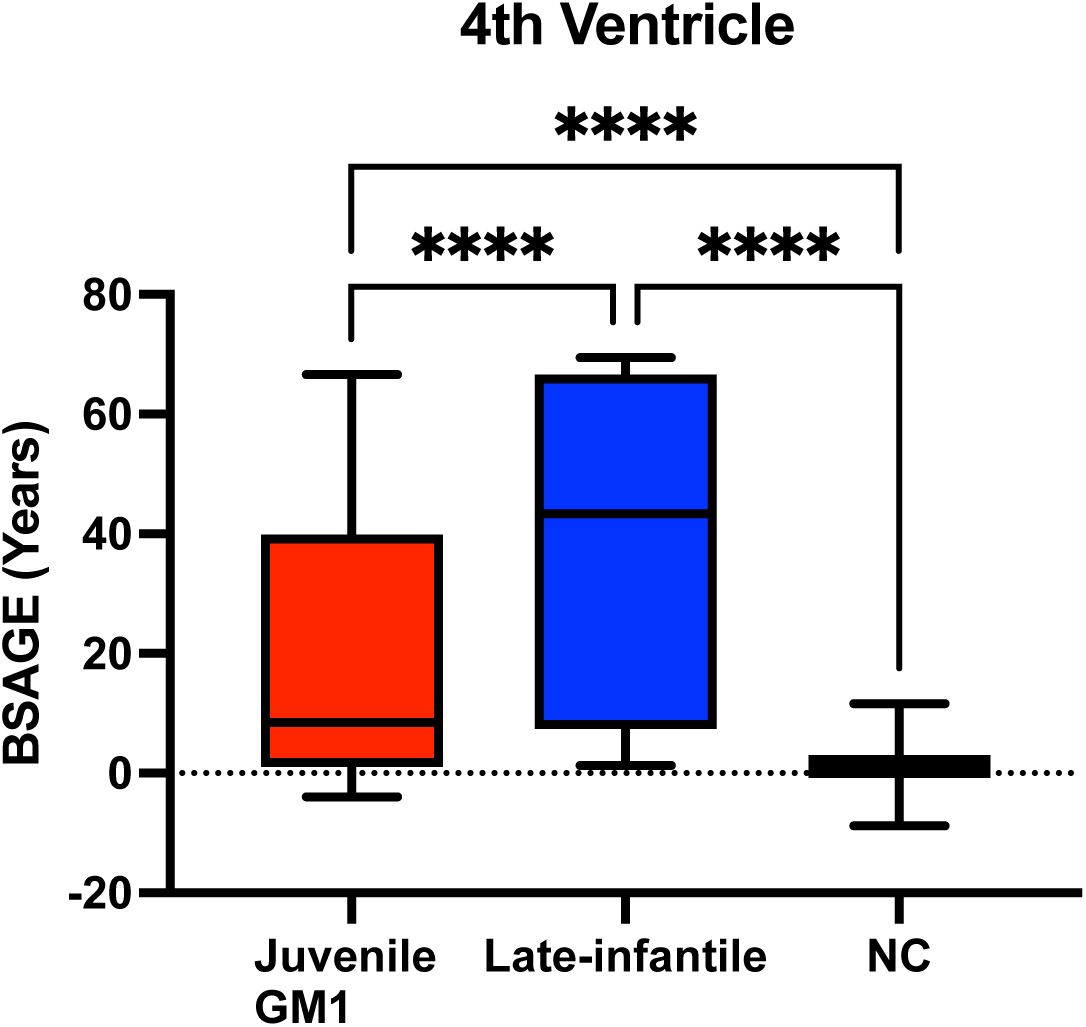
Comparison of the 4^rd^ ventricle BSAGE between Juvenile GM1 patients, late-infantile GM1 patients, and neurotypical controls (NC). Late-infantile GM1 patients had an average BSAGE = 37.52, juvenile GM1 patients had an average BSAGE = 20.09, and NC participants had an average BSAGE = 0.96. An analysis of variance (ANOVA) showed statistically significant (F(2, 595) = 438.0, *p* < 0.0001) differences in BSAGE between the cohorts. A post-hoc Tukey test showed the late-infantile cohort had a statistically significantly higher BSAGE compared to both the neurotypical controls (*p* < 0.0001) and juvenile GM1 patients (*p* < 0.0001). The juvenile GM1 cohort also had a significantly higher BSAGE compared to the neurotypical controls (*p* < 0.0001).

**Figure J12.**
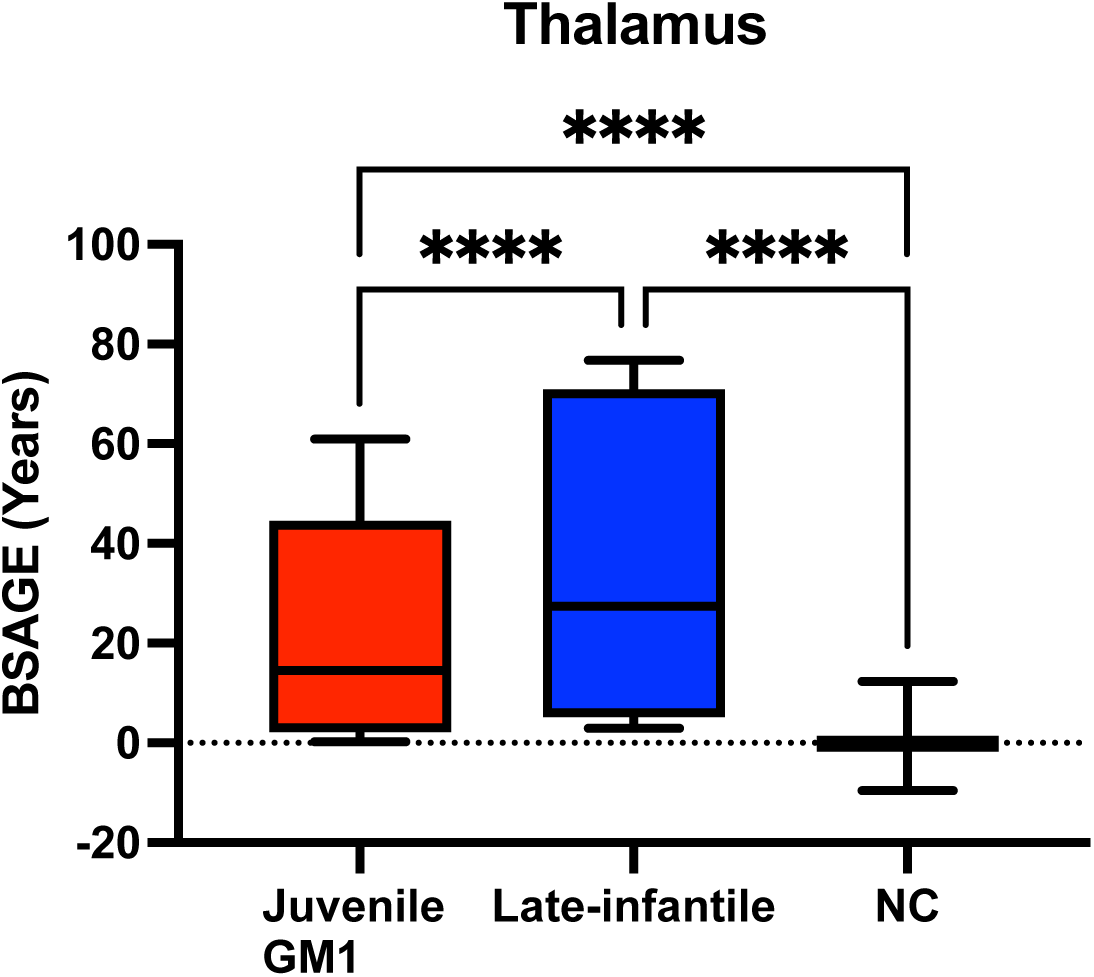
Comparison of the thalamus BSAGE between Juvenile GM1 patients, late-infantile GM1 patients, and neurotypical controls (NC). Late-infantile GM1 patients had an average BSAGE = 38.41, juvenile GM1 patients had an average BSAGE = 22.49, and NC participants had an average BSAGE = −0.24. An analysis of variance (ANOVA) showed statistically significant (F(2, 595) = 558.1, *p* < 0.0001) differences in BSAGE between the cohorts. A post-hoc Tukey test showed the late-infantile cohort had a statistically significantly higher BSAGE compared to both the neurotypical controls (*p* < 0.0001) and juvenile GM1 patients (*p* < 0.0001). The juvenile GM1 cohort also had a significantly higher BSAGE compared to the neurotypical controls (*p* < 0.0001).

**Figure J13.**
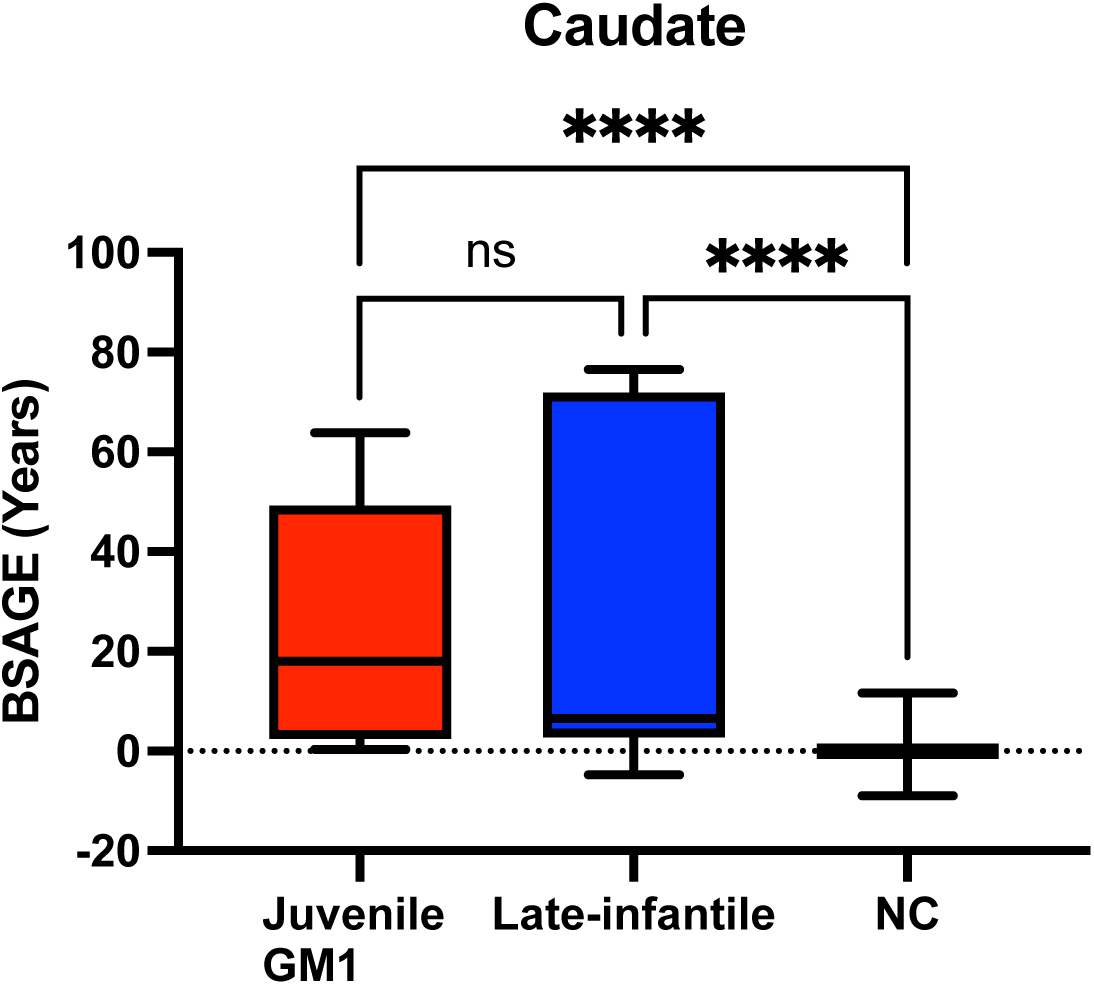
Comparison of the Caudate Nucleus BSAGE between Juvenile GM1 patients, late-infantile GM1 patients, and neurotypical controls (NC). Late-infantile GM1 patients had an average BSAGE = 30.64, juvenile GM1 patients had an average BSAGE = 25.34, and NC participants had an average BSAGE = −0.11. An analysis of variance (ANOVA) showed statistically significant (F(2, 595) = 289.0, *p* < 0.0001) differences in BSAGE between the cohorts. A post-hoc Tukey test showed the late-infantile cohort had a statistically significantly higher BSAGE compared to both the neurotypical controls (*p* < 0.0001) and juvenile GM1 patients (*p* < 0.0001). The juvenile GM1 cohort also had a significantly higher BSAGE compared to the neurotypical controls (*p* < 0.0001).

**Figure J14.**
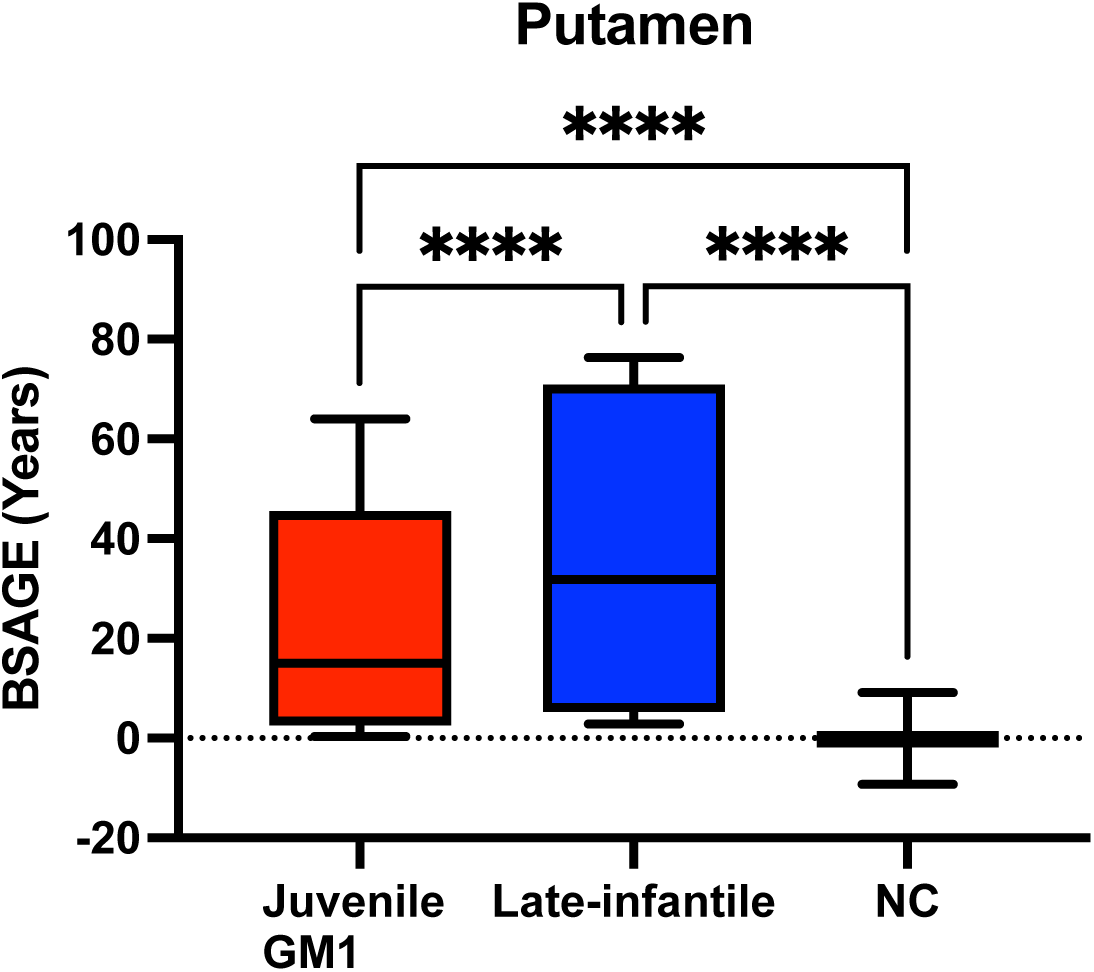
Comparison of the Putamen BSAGE between Juvenile GM1 patients, late-infantile GM1 patients, and neurotypical controls (NC). Late-infantile GM1 patients had an average BSAGE = 38.20, juvenile GM1 patients had an average BSAGE = 25.05, and NC participants had an average BSAGE = −0.19. An analysis of variance (ANOVA) showed statistically significant (F(2, 595) = 660.8, *p* < 0.0001) differences in BSAGE between the cohorts. A post-hoc Tukey test showed the late-infantile cohort had a statistically significantly higher BSAGE compared to both the neurotypical controls (*p* < 0.0001) and juvenile GM1 patients (*p* < 0.0001). The juvenile GM1 cohort also had a significantly higher BSAGE compared to the neurotypical controls (*p* < 0.0001).

**Figure J15.**
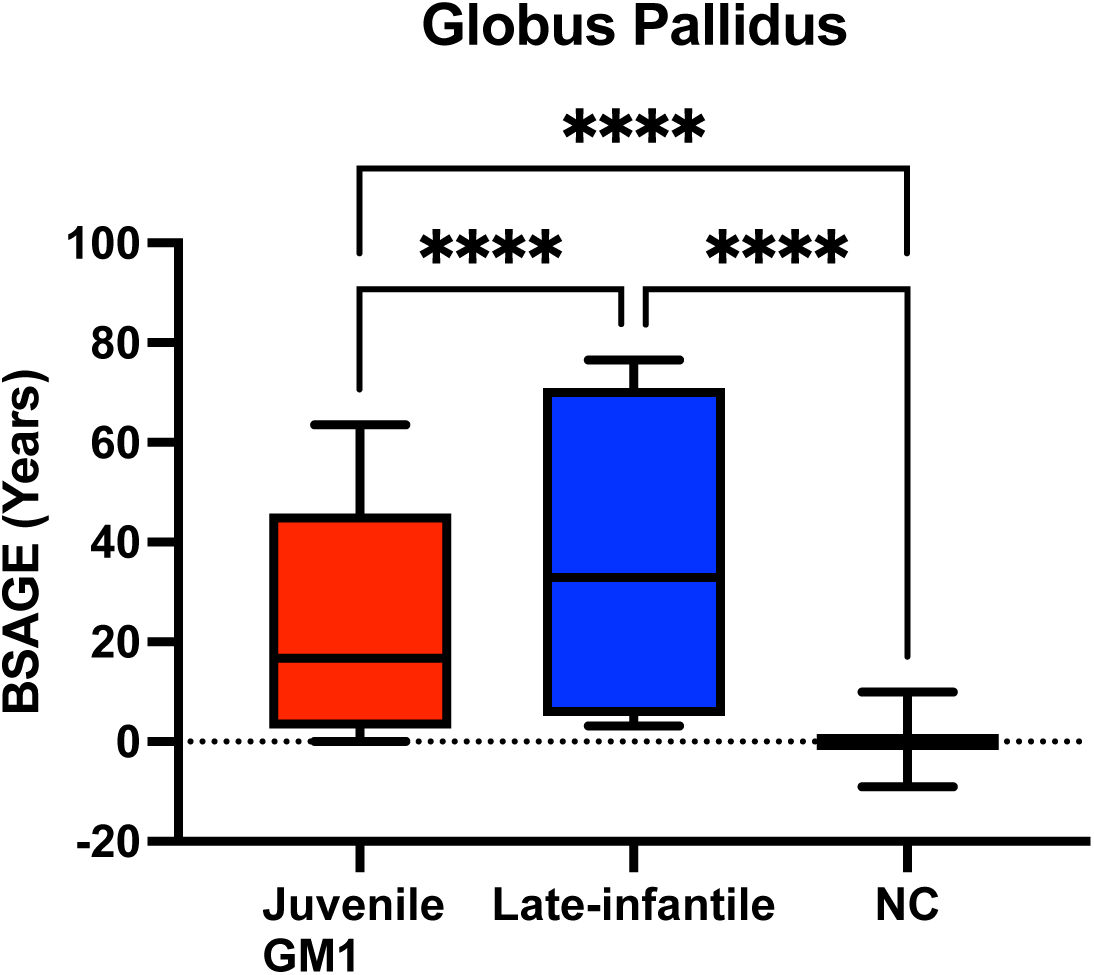
Comparison of the Globus Pallidus BSAGE between Juvenile GM1 patients, late-infantile GM1 patients, and neurotypical controls (NC). Late-infantile GM1 patients had an average BSAGE = 37.82, juvenile GM1 patients had an average BSAGE = 24.44, and NC participants had an average BSAGE = −0.10. An analysis of variance (ANOVA) showed statistically significant (F(2, 595) = 438.0, *p* < 0.0001) differences in BSAGE between the cohorts. A post-hoc Tukey test showed the late-infantile cohort had a statistically significantly higher BSAGE compared to both the neurotypical controls (*p* < 0.0001) and juvenile GM1 patients (*p* < 0.0001). The juvenile GM1 cohort also had a significantly higher BSAGE compared to the neurotypical controls (*p* < 0.0001).

**Figure J16.**
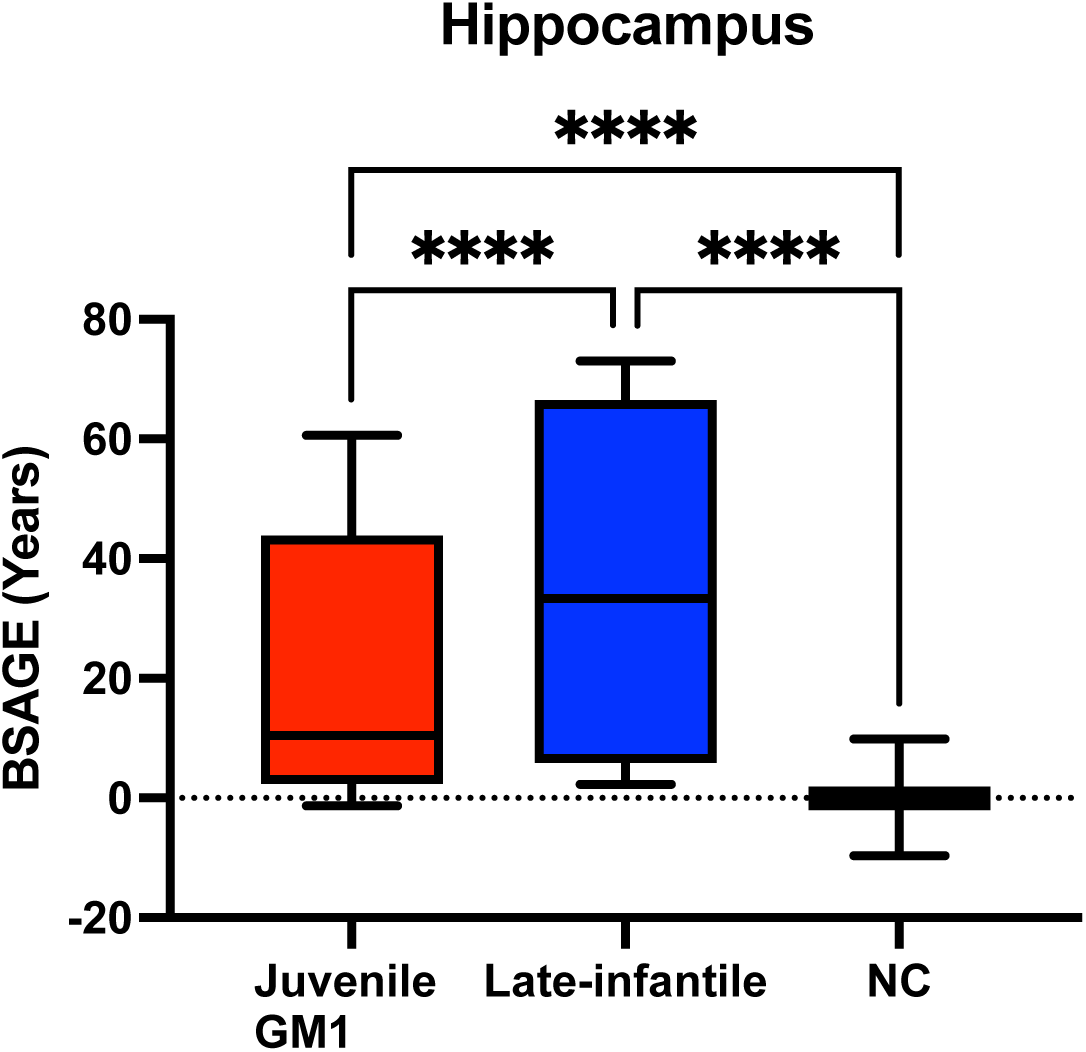
Comparison of the hippocampus BSAGE between Juvenile GM1 patients, late-infantile GM1 patients, and neurotypical controls (NC). Late-infantile GM1 patients had an average BSAGE = 38.10, juvenile GM1 patients had an average BSAGE = 21.87, and NC participants had an average BSAGE = 0.01. An analysis of variance (ANOVA) showed statistically significant (F(2, 595) = 479.9, *p* < 0.0001) differences in BSAGE between the cohorts. A post-hoc Tukey test showed the late-infantile cohort had a statistically significantly higher BSAGE compared to both the neurotypical controls (*p* < 0.0001) and juvenile GM1 patients (*p* < 0.0001). The juvenile GM1 cohort also had a significantly higher BSAGE compared to the neurotypical controls (*p* < 0.0001).

**Figure J17.**
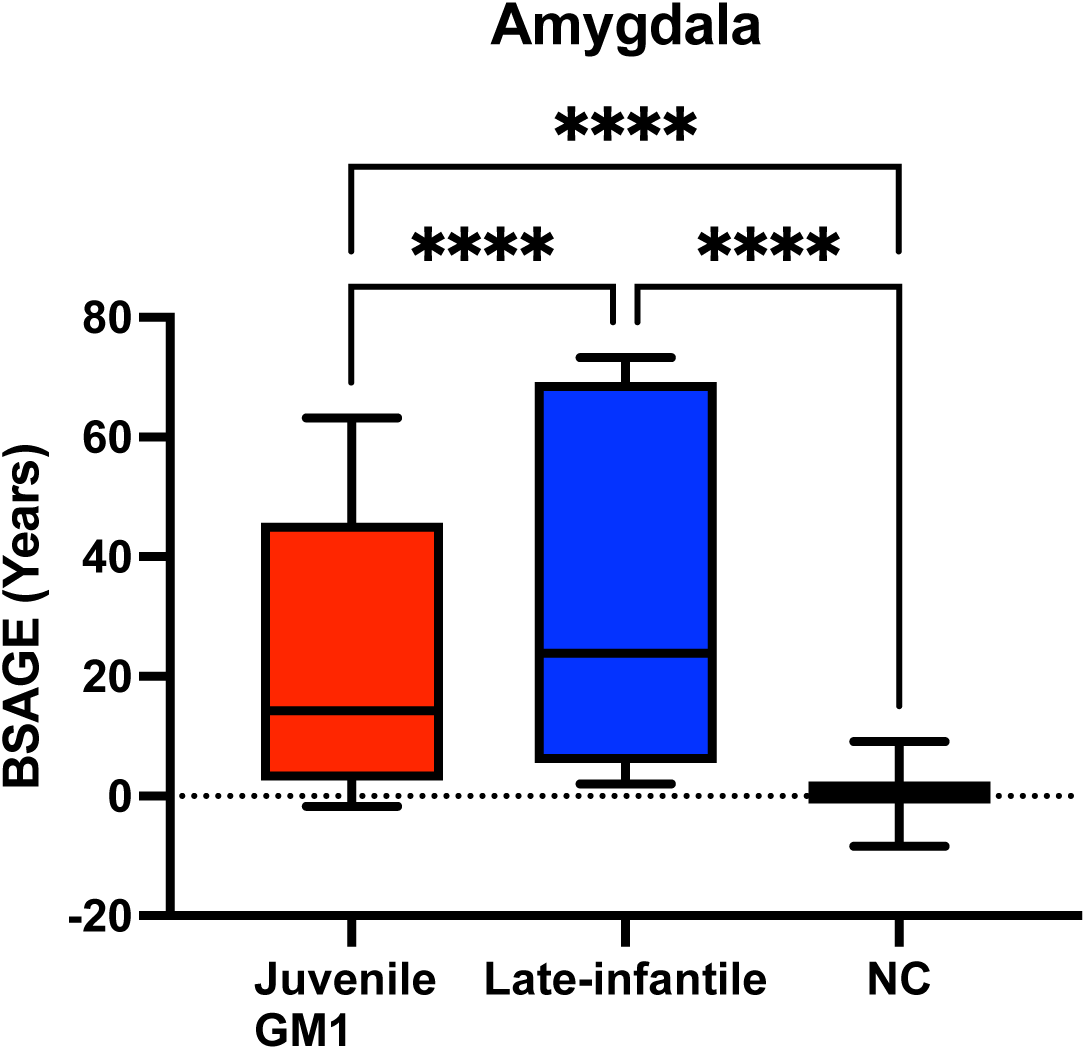
Comparison of the amygdala BSAGE between Juvenile GM1 patients, late-infantile GM1 patients, and neurotypical controls (NC). Late-infantile GM1 patients had an average BSAGE = 36.78, juvenile GM1 patients had an average BSAGE = 23.57, and NC participants had an average BSAGE = 0.55. An analysis of variance (ANOVA) showed statistically significant (F(2, 595) = 514.7, *p* < 0.0001) differences in BSAGE between the cohorts. A post-hoc Tukey test showed the late-infantile cohort had a statistically significantly higher BSAGE compared to both the neurotypical controls (*p* < 0.0001) and juvenile GM1 patients (*p* < 0.0001). The juvenile GM1 cohort also had a significantly higher BSAGE compared to the neurotypical controls (*p* < 0.0001).

**Figure J18.**
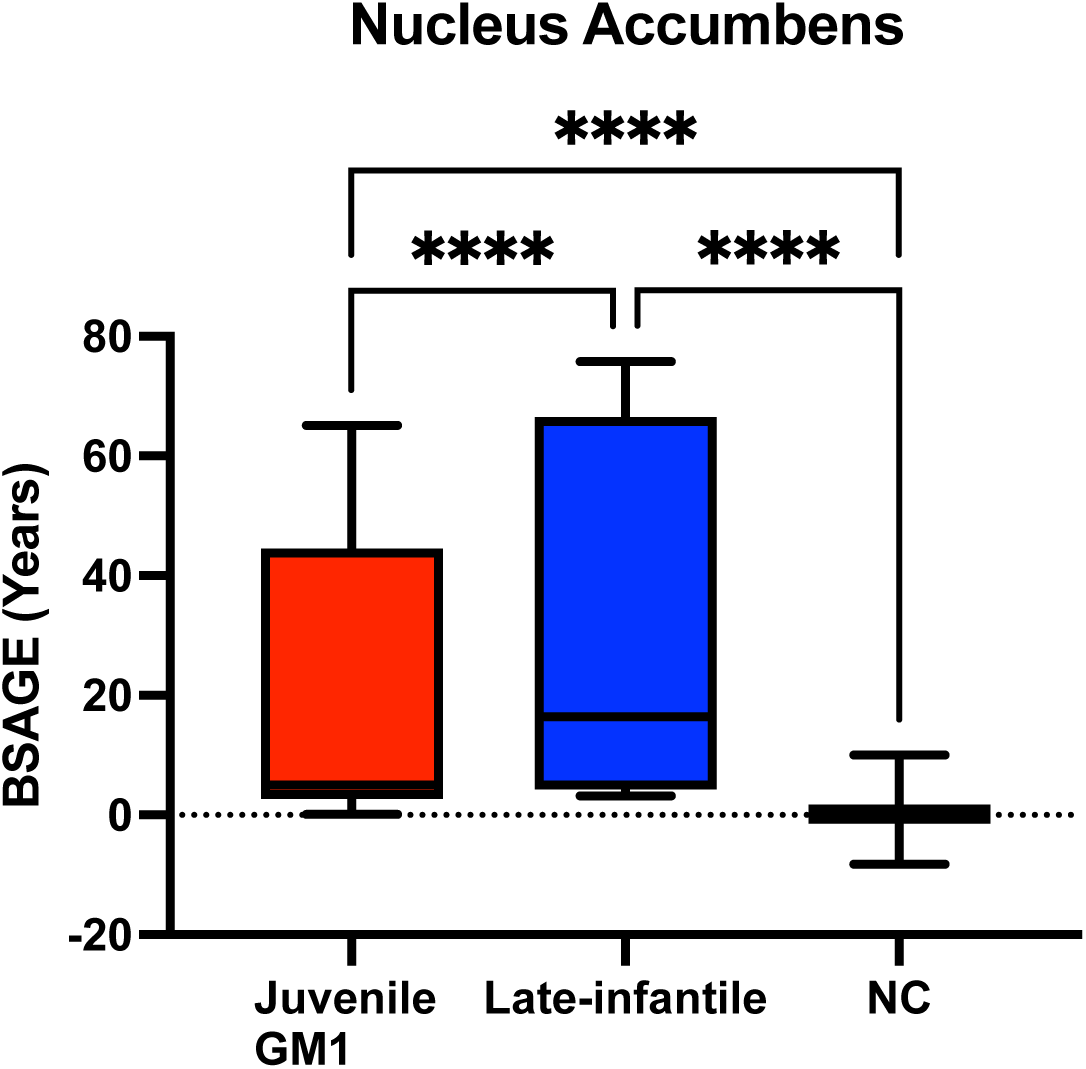
Comparison of the nucleus accumbens (NAC) BSAGE between Juvenile GM1 patients, late-infantile GM1 patients, and neurotypical controls (NC). Late-infantile GM1 patients had an average BSAGE = 30.56, juvenile GM1 patients had an average BSAGE = 23.46, and NC participants had an average BSAGE = 0.18. An analysis of variance (ANOVA) showed statistically significant (F(2, 595) = 373.2, *p* < 0.0001) differences in BSAGE between the cohorts. A post-hoc Tukey test showed the late-infantile cohort had a statistically significantly higher BSAGE compared to both the neurotypical controls (*p* < 0.0001) and juvenile GM1 patients (*p* < 0.0001). The juvenile GM1 cohort also had a significantly higher BSAGE compared to the neurotypical controls (*p* < 0.0001).

**Figure J19.**
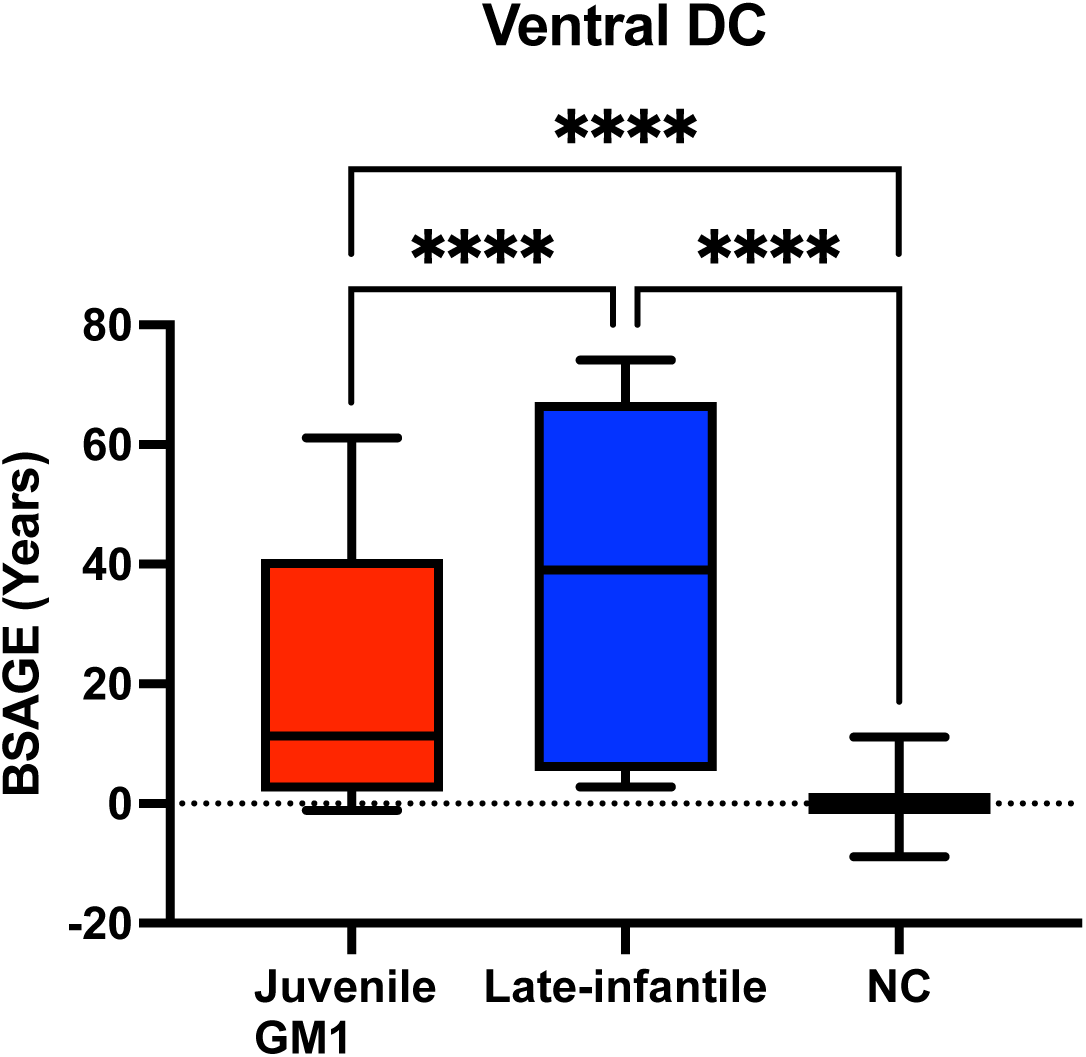
Comparison of the ventral diencephalon BSAGE between Juvenile GM1 patients, late-infantile GM1 patients, and neurotypical controls (NC). Late-infantile GM1 patients had an average BSAGE = 39.12, juvenile GM1 patients had an average BSAGE = 21.10, and NC participants had an average BSAGE = 0.08. An analysis of variance (ANOVA) showed statistically significant (F(2, 595) = 532.3, *p* < 0.0001) differences in BSAGE between the cohorts. A post-hoc Tukey test showed the late-infantile cohort had a statistically significantly higher BSAGE compared to both the neurotypical controls (*p* < 0.0001) and juvenile GM1 patients (*p* < 0.0001). The juvenile GM1 cohort also had a significantly higher BSAGE compared to the neurotypical controls (*p* < 0.0001).

**Figure J20.**
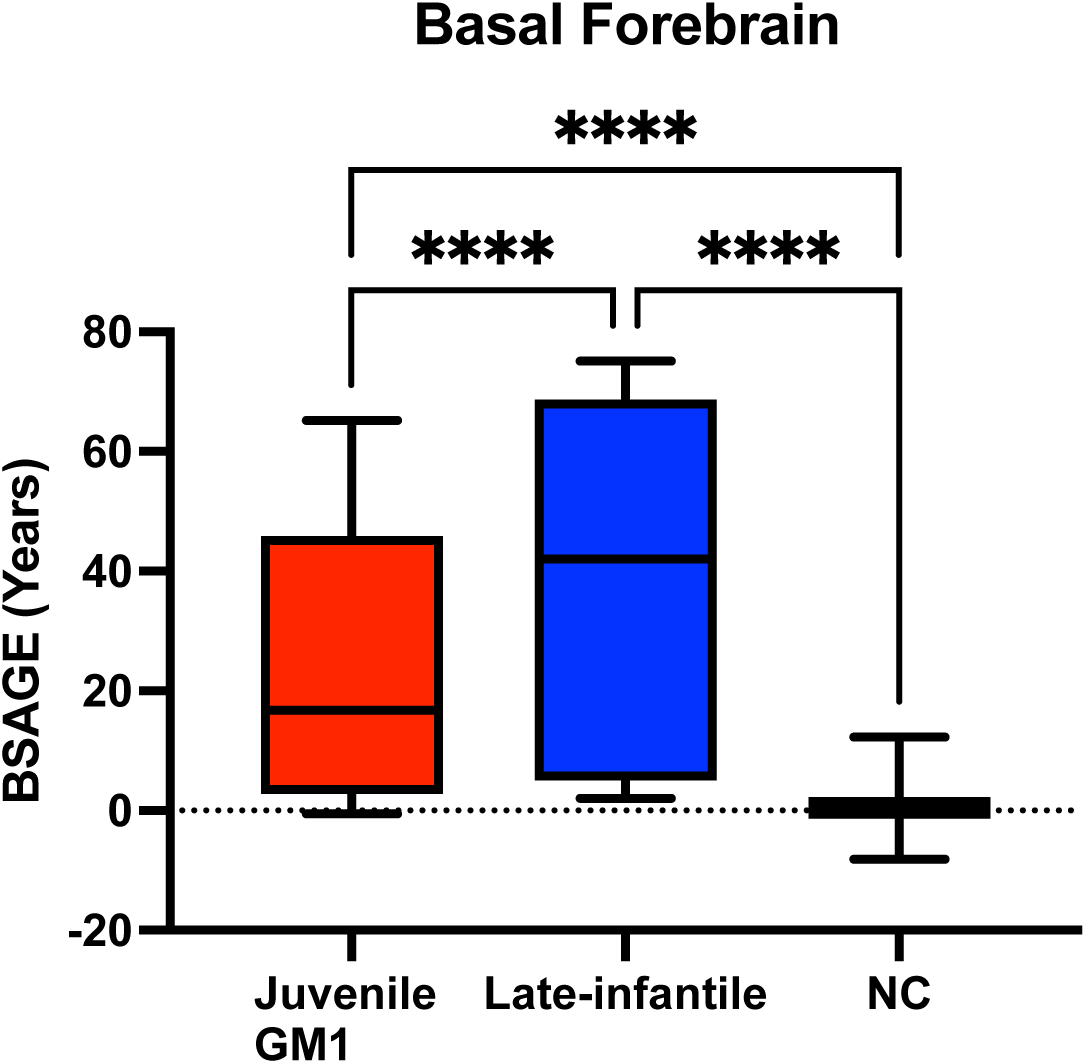
Comparison of the basal forebrain BSAGE between Juvenile GM1 patients, late-infantile GM1 patients, and neurotypical controls (NC). Late-infantile GM1 patients had an average BSAGE = 39.00, juvenile GM1 patients had an average BSAGE = 24.66, and NC participants had an average BSAGE = 0.45. An analysis of variance (ANOVA) showed statistically significant (F(2, 595) = 717.8, *p* < 0.0001) differences in BSAGE between the cohorts. A post-hoc Tukey test showed the late-infantile cohort had a statistically significantly higher BSAGE compared to both the neurotypical controls (*p* < 0.0001) and juvenile GM1 patients (*p* < 0.0001). The juvenile GM1 cohort also had a significantly higher BSAGE compared to the neurotypical controls (*p* < 0.0001).

**Figure J21.**
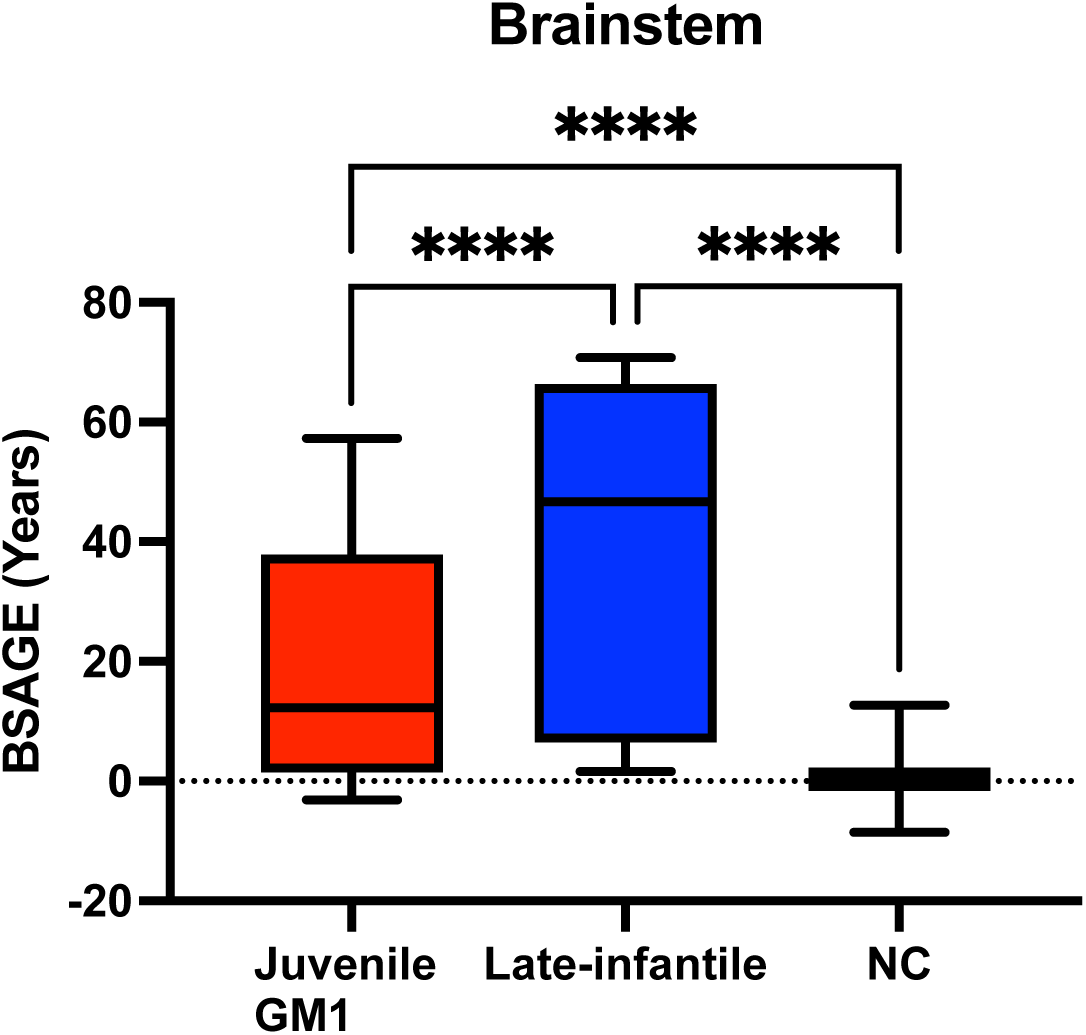
Comparison of the brainstem BSAGE between Juvenile GM1 patients, late-infantile GM1 patients, and neurotypical controls (NC). Late-infantile GM1 patients had an average BSAGE = 38.56, juvenile GM1 patients had an average BSAGE = 19.56, and NC participants had an average BSAGE = 0.40. An analysis of variance (ANOVA) showed statistically significant (F(2, 595) = 429.3, *p* < 0.0001) differences in BSAGE between the cohorts. A post-hoc Tukey test showed the late-infantile cohort had a statistically significantly higher BSAGE compared to both the neurotypical controls (*p* < 0.0001) and juvenile GM1 patients (*p* < 0.0001). The juvenile GM1 cohort also had a significantly higher BSAGE compared to the neurotypical controls (*p* < 0.0001).

### Supplement K: Comparisons Between Predicted Brain Age and Brain Volumetrics

**Figure K1.**
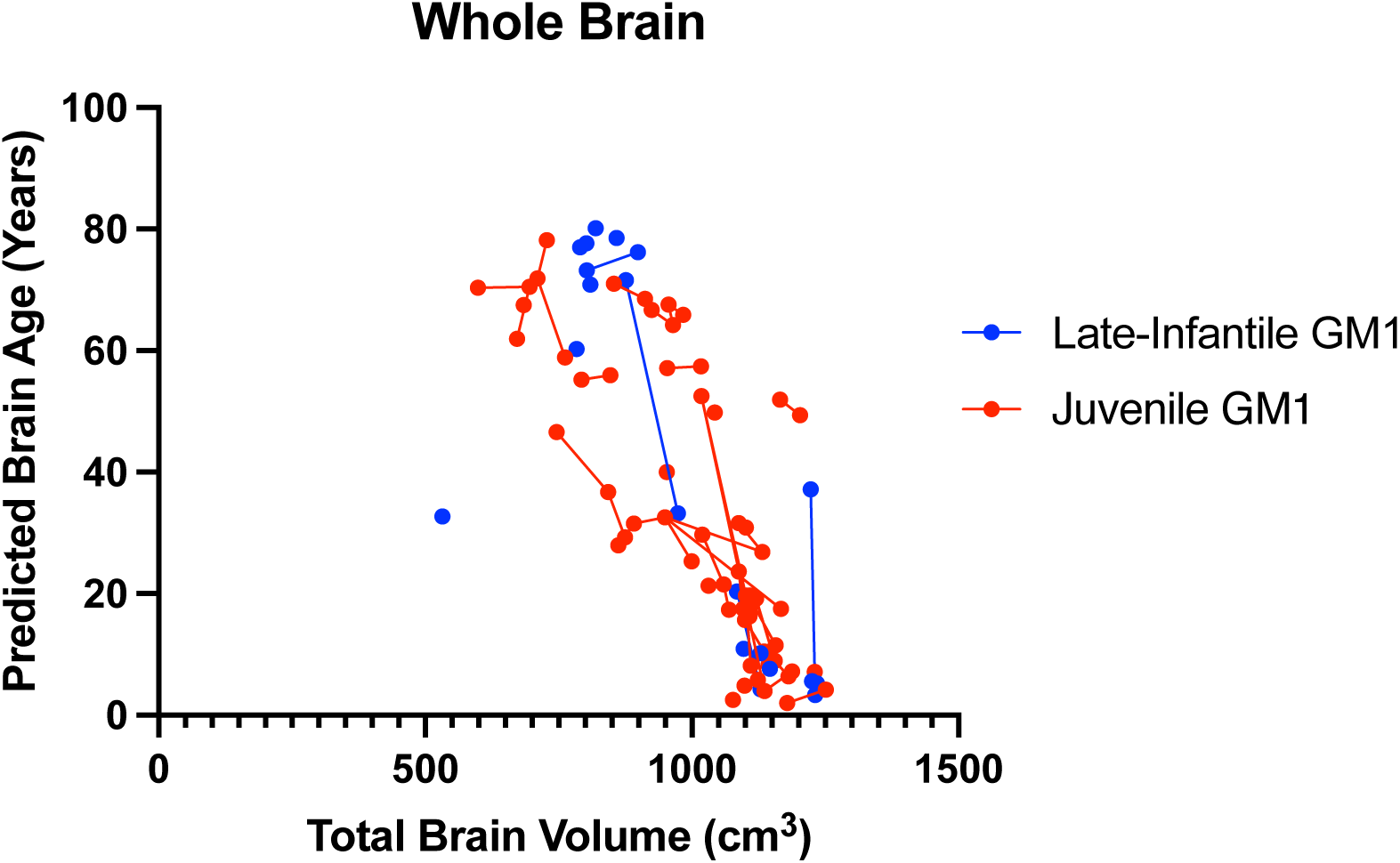
Correlation between whole brain volume (cm^3^) and predicted whole brain age in juvenile and late-infantile GM1 patients. Total brain volume correlated with predicted whole brain age in both the juvenile (R^2^ = 0.57) and late-infantile (R^2^ = 0.46) patients.

**Figure K2.**
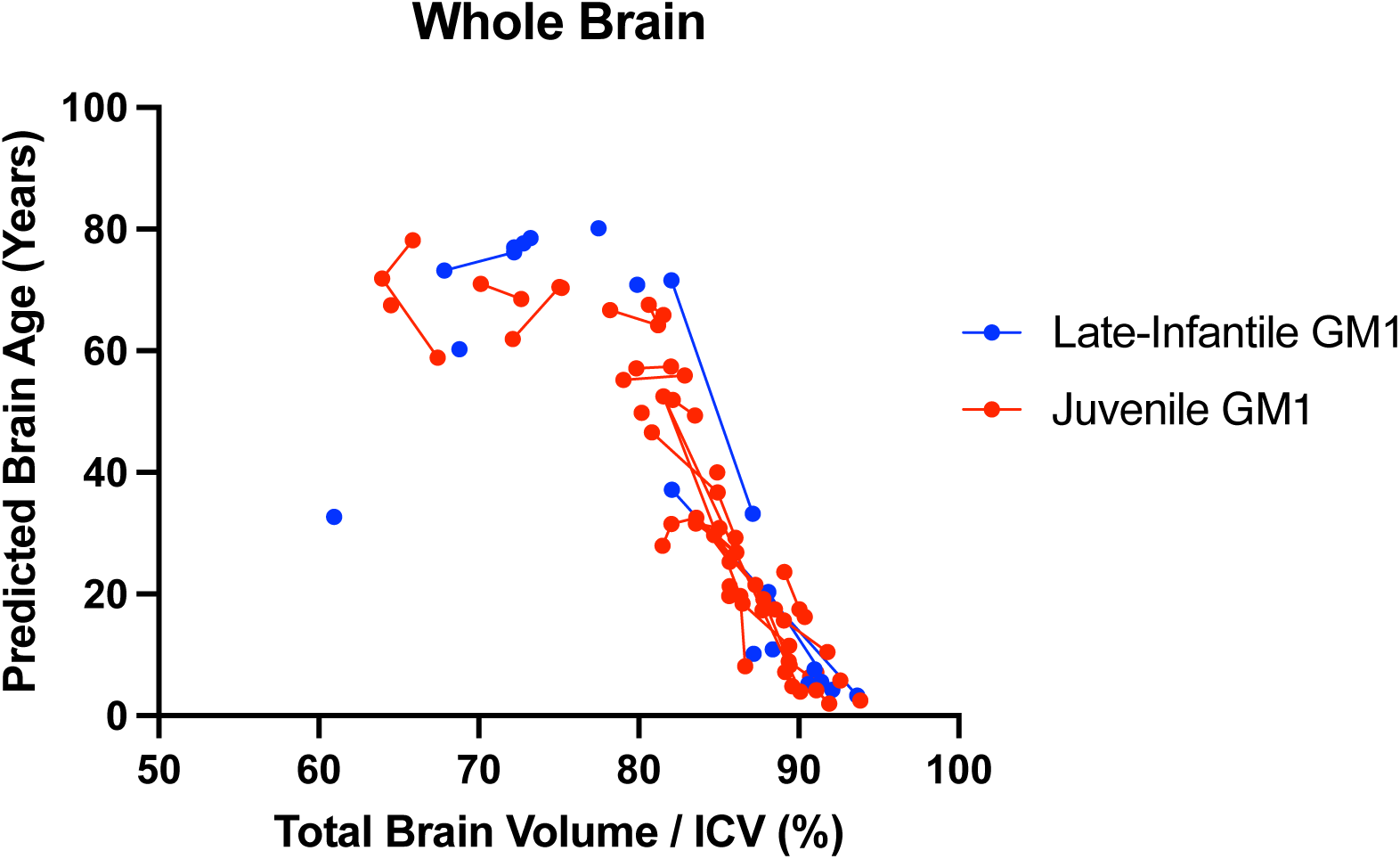
Correlation between whole brain volume controlled for total intracranial volume (ICV) and predicted whole brain age in juvenile and late-infantile GM1 patients. Total brain volume correlated with predicted whole brain age in both the juvenile (R^2^ = 0.77) and late-infantile (R^2^ = 0.51) patients.

**Figure K3.**
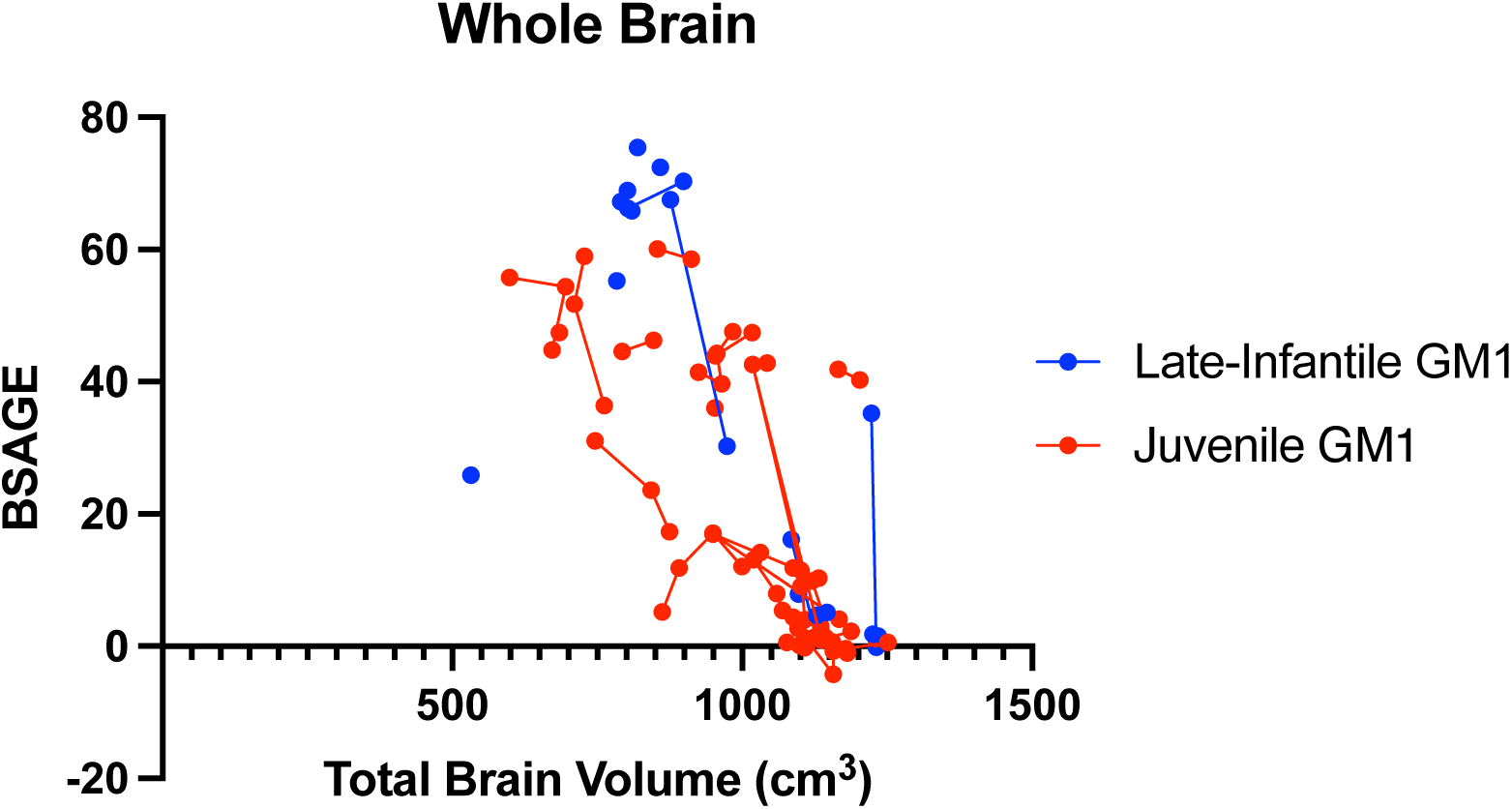
Correlation between whole brain volume (cm^3^) and predicted whole brain, brain structures age gap estimation (BSAGE) in juvenile and late-infantile GM1 patients. Total brain volume correlated with predicted whole brain age in both the juvenile (R^2^ = 0.49) and late-infantile (R^2^ = 0.43) patients.

**Figure K4.**
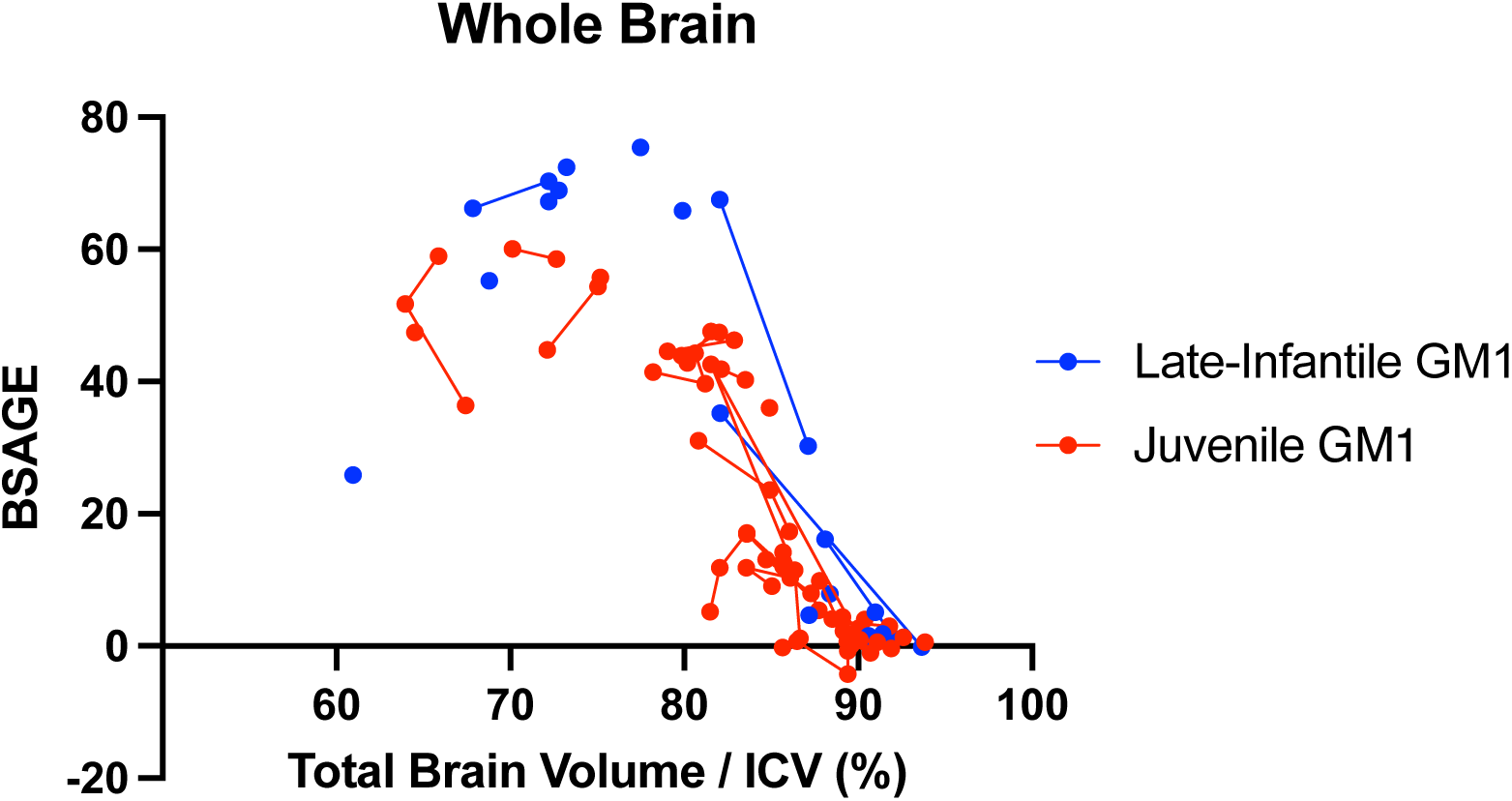
Correlation between whole brain volume controlled for total intracranial volume (ICV) and predicted whole brain, brain structures age gap estimation (BSAGE) in juvenile and late-infantile GM1 patients. Total brain volume correlated with predicted whole brain age in both the juvenile (R^2^ = 0.69) and late-infantile (R^2^ = 0.47) patients.

**Figure K5.**
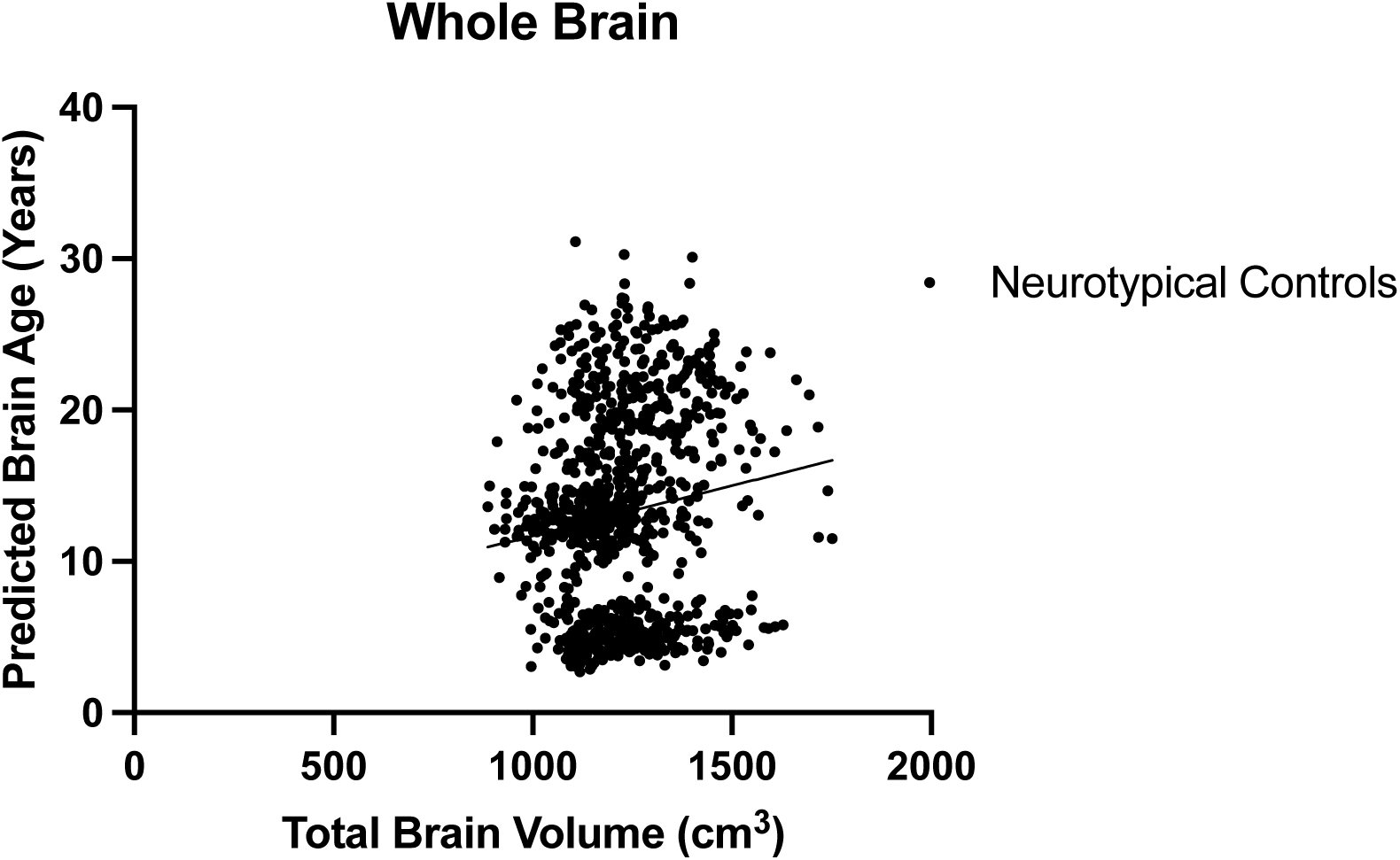
Correlation between whole brain volume (cm^3^) and predicted whole brain age in neurotypical controls. Total brain volume correlated with predicted whole brain age in the neurotypical controls (R^2^ = 0.02).

**Figure K6.**
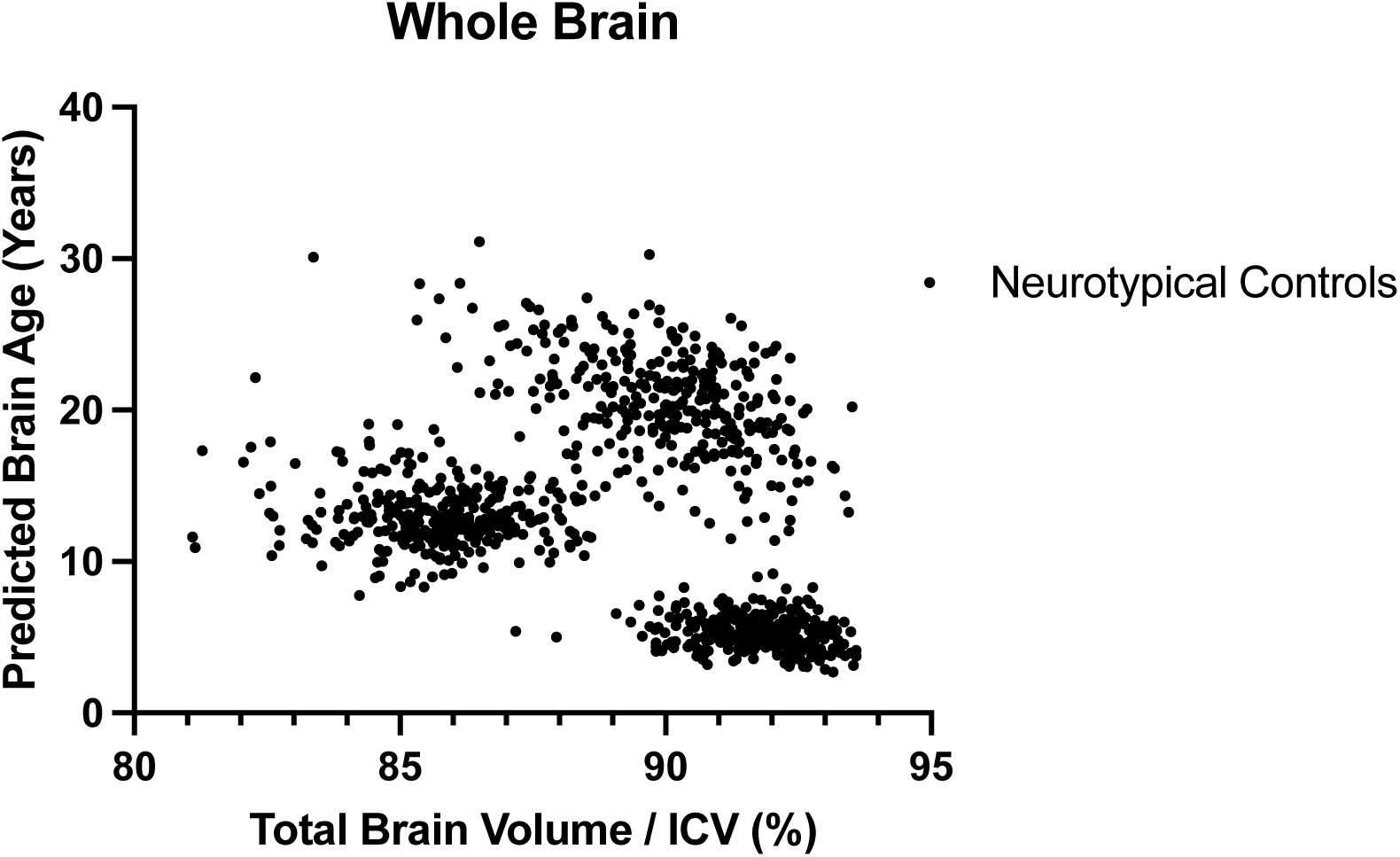
Correlation between whole brain volume controlled for total intracranial volume (ICV) and predicted whole brain age in neurotypical controls. Total brain volume correlated with predicted whole brain age in the neurotypical controls (R^2^ = 0.07).

**Figure K7.**
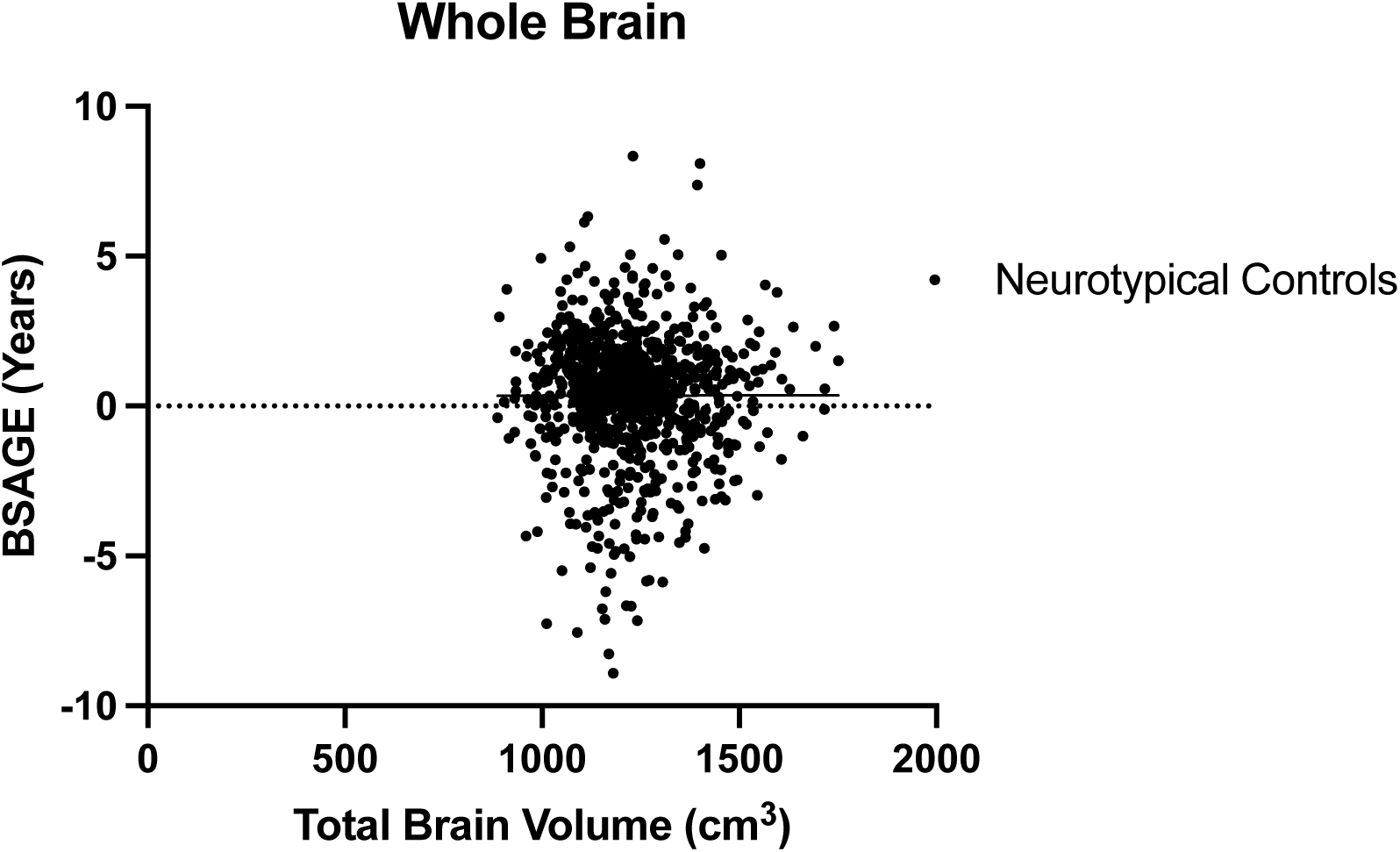
Correlation between whole brain volume (cm^3^) and predicted whole brain, brain structures age gap estimation (BSAGE) in neurotypical controls. Total brain volume did not correlate with predicted whole brain age in the neurotypical controls (R^2^ < 0.01).

**Figure K8.**
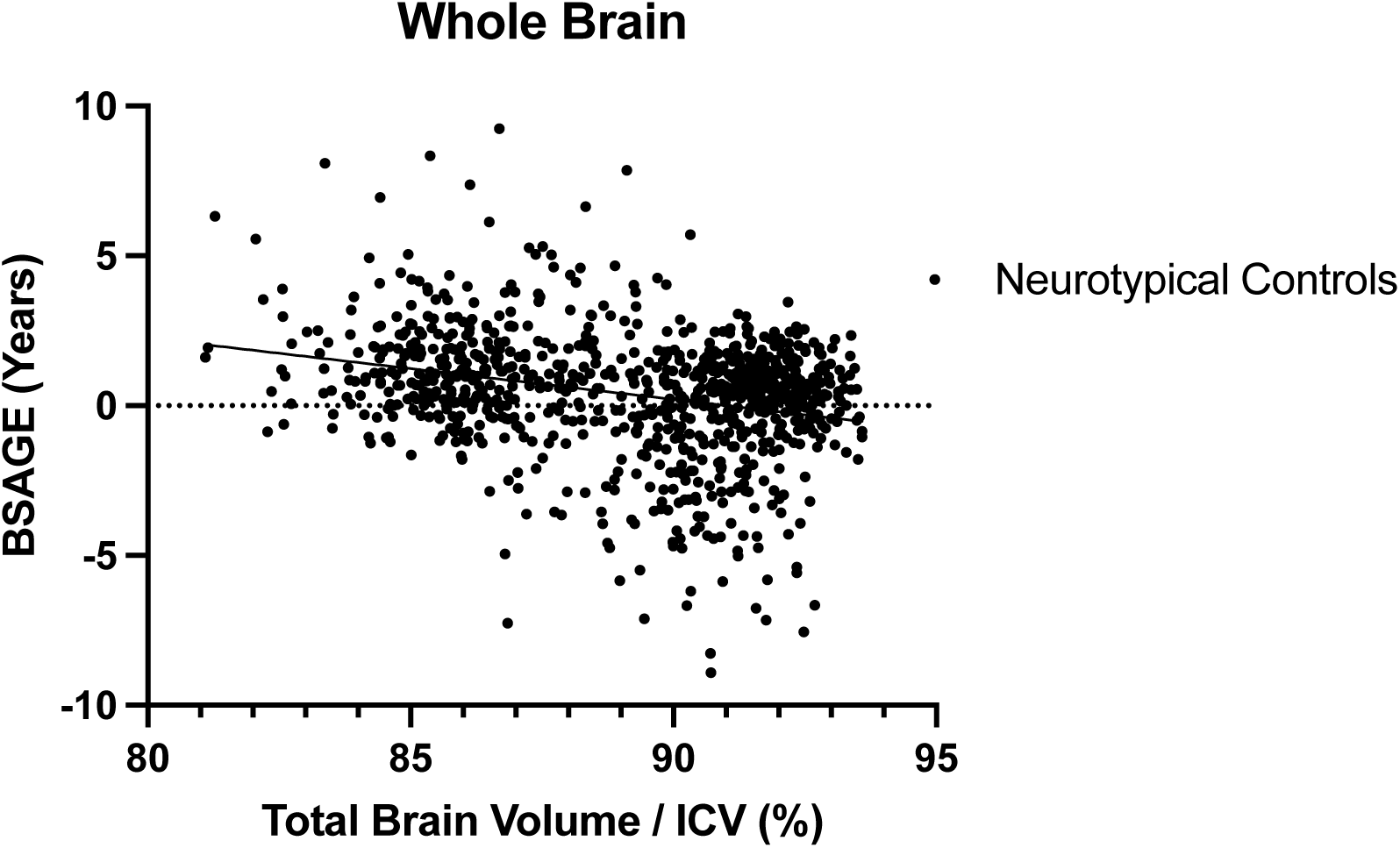
Correlation between whole brain volume controlled for total intracranial volume (ICV) and predicted whole brain, brain structures age gap estimation (BSAGE) in neurotypical controls. Total brain volume correlated with predicted whole brain age in the neurotypical controls (R^2^ = 0.08).

**Figure K9.**
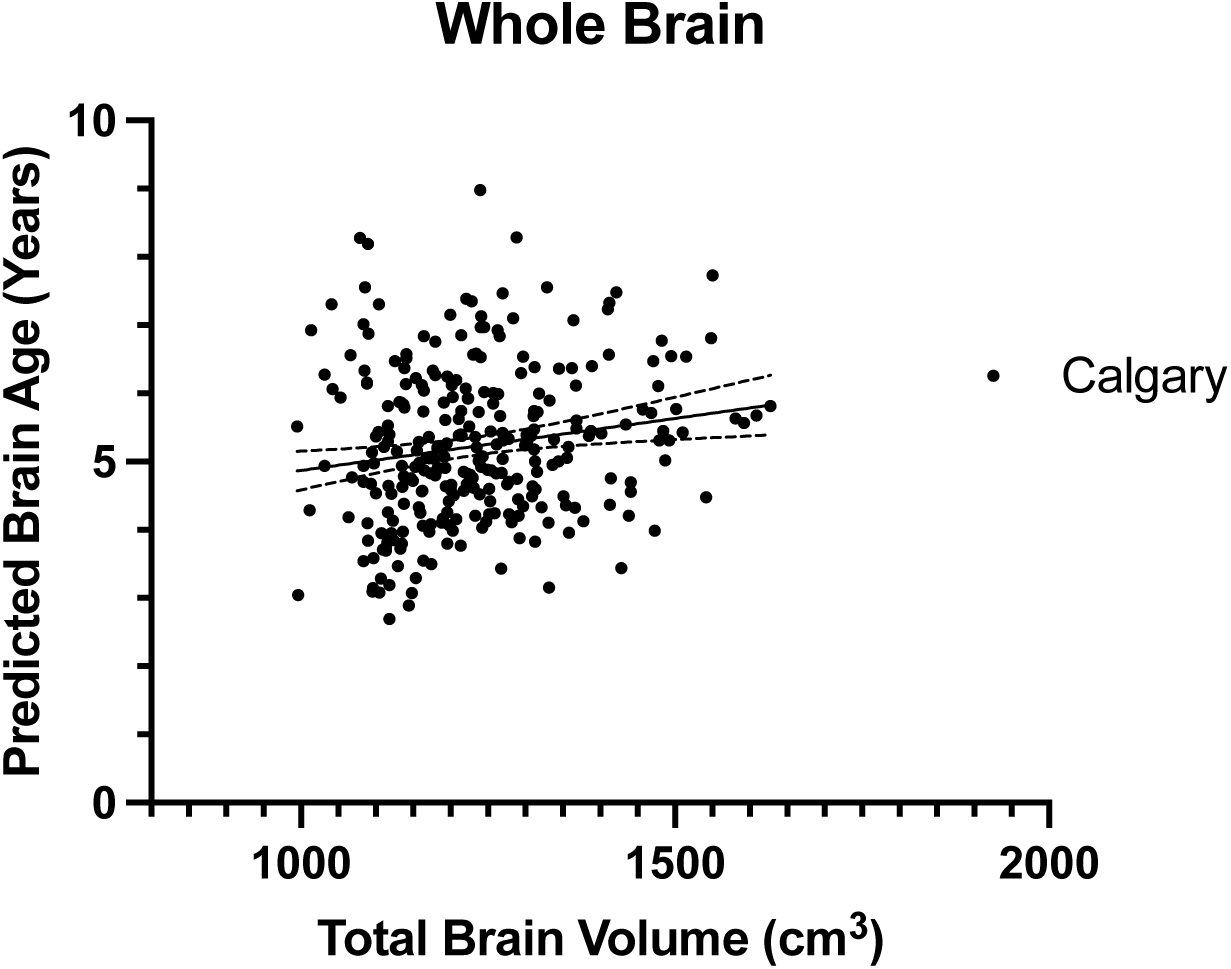
Correlation between whole brain volume (cm^3^) and predicted whole brain age in Calgary neurotypical controls. Total brain volume did correlated with predicted whole brain age in Calgary neurotypical controls (R^2^ = 0.03).

**Figure K10.**
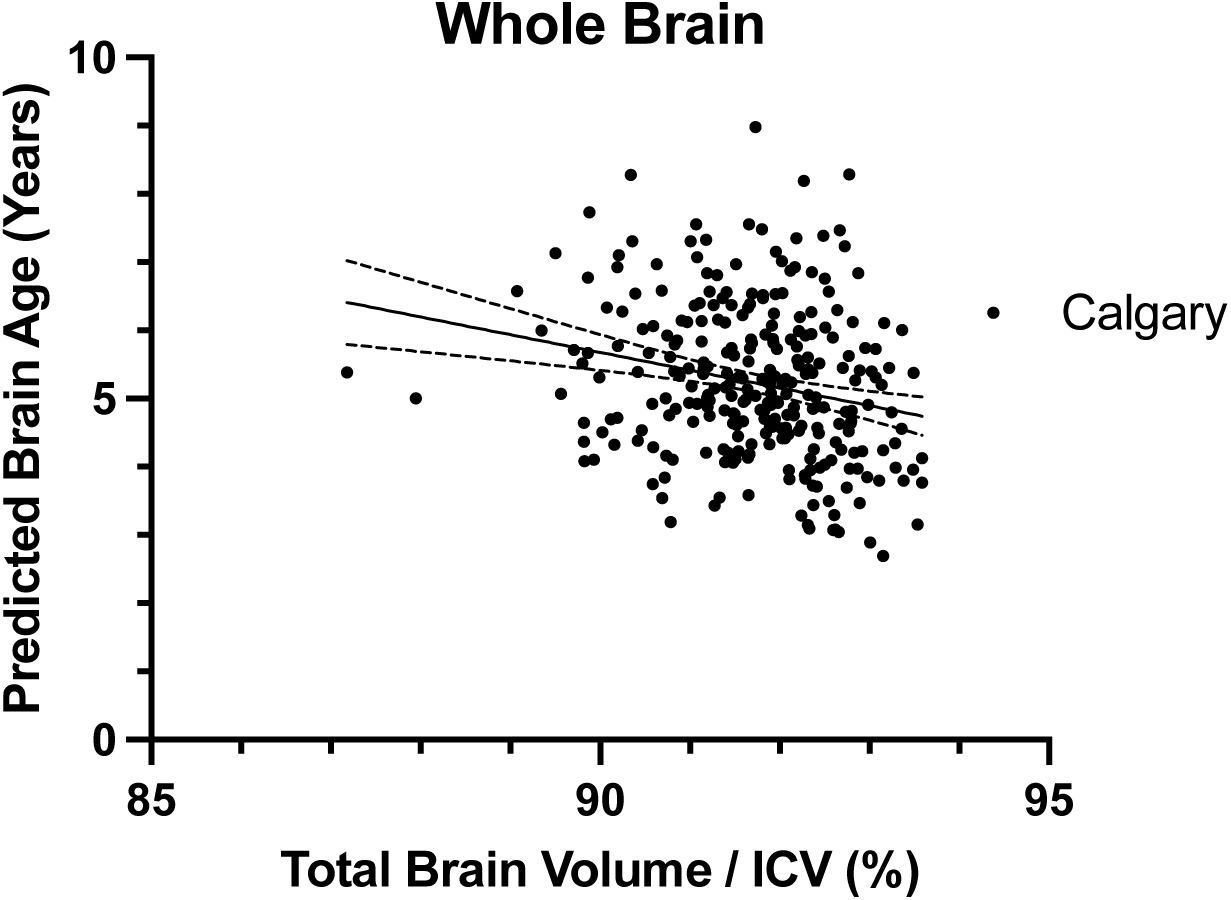
Correlation between whole brain volume controlled for total intracranial volume (ICV) and predicted whole brain, brain structures age gap estimation (BSAGE) in Calgary neurotypical controls. Total brain volume correlated with predicted whole brain age in the Calgary neurotypical controls (R^2^ = 0.05).

**Figure K11.**
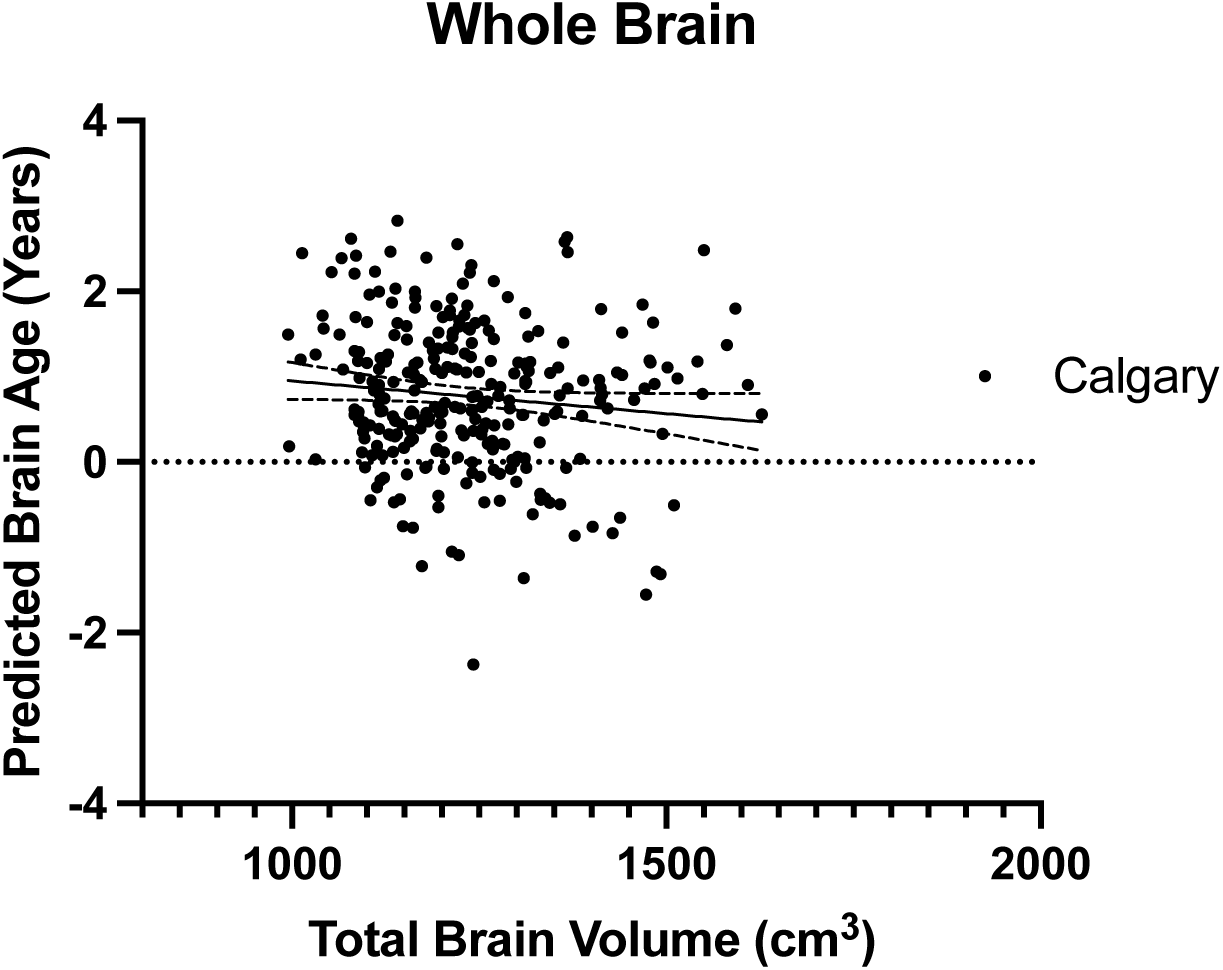
Correlation between whole brain volume (cm^3^) and predicted whole brain, brain structures age gap estimation (BSAGE) in neurotypical controls. Total brain volume did not correlate with predicted whole brain age in the neurotypical controls (R^2^ = 0.01).

**Figure K12.**
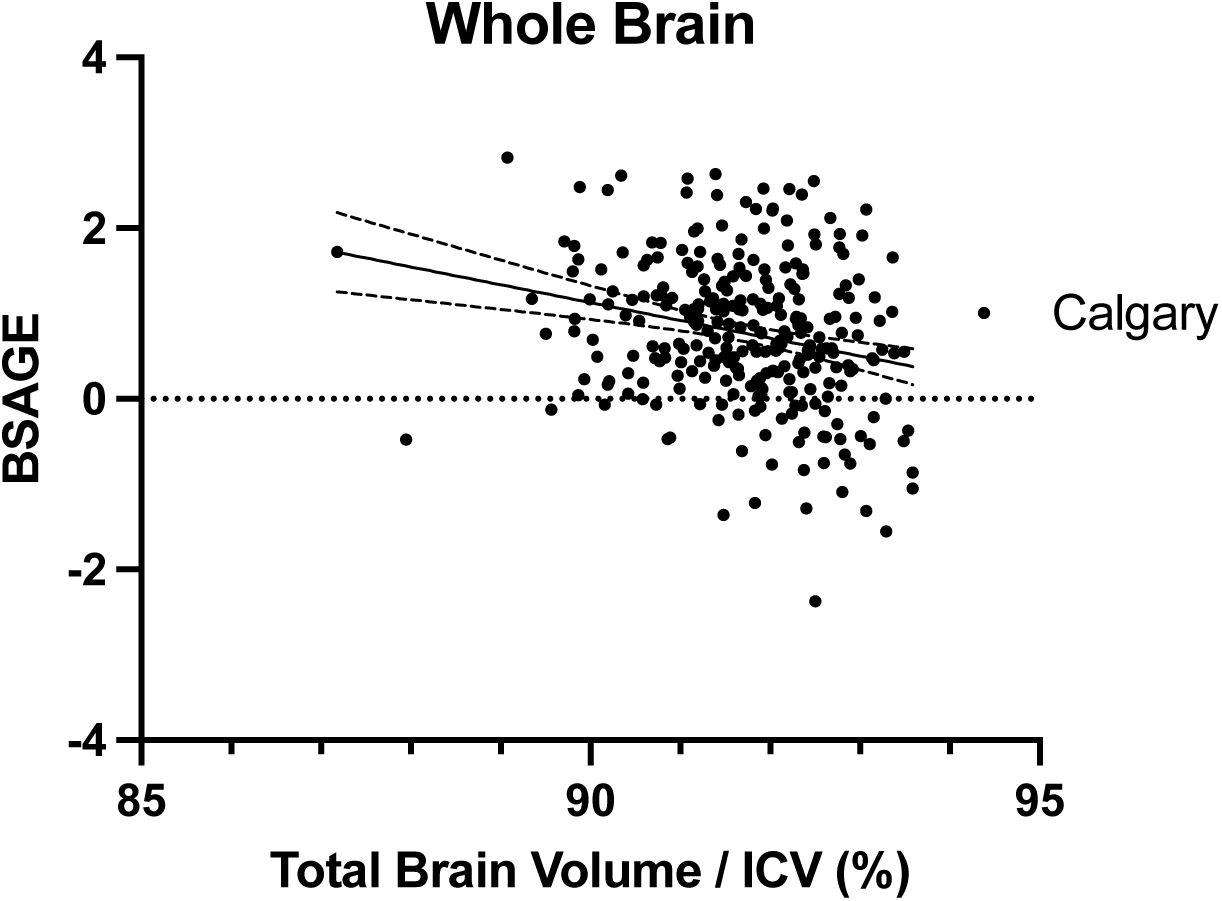
Correlation between whole brain volume controlled for ICV and predicted whole brain, brain structures age gap estimation (BSAGE) in neurotypical controls. Total brain volume did not correlate with predicted whole brain age in the neurotypical controls (R^2^ = 0.06).

**Figure K13.**
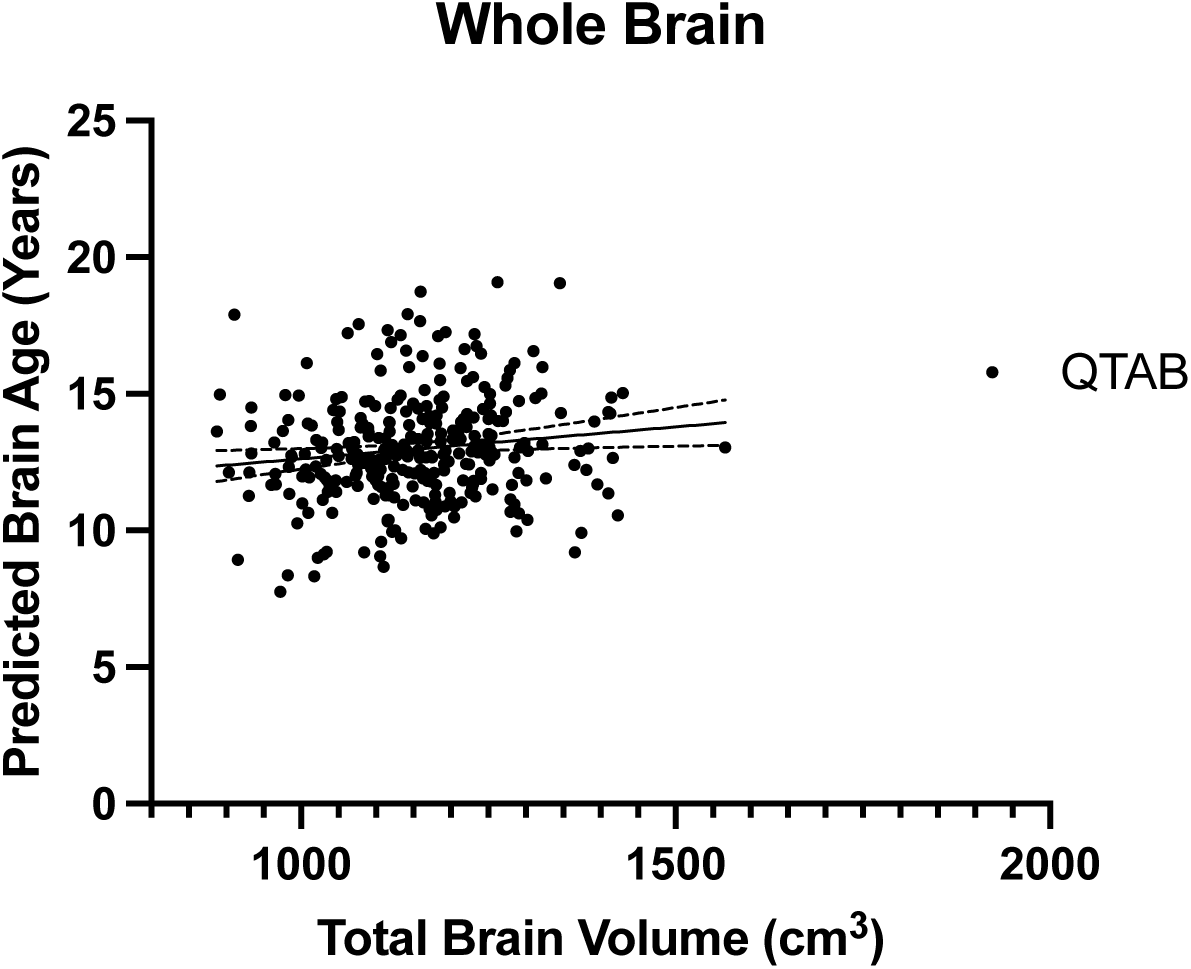
Correlation between whole brain volume and predicted whole brain age gap estimation in QTAB neurotypical controls. Total brain volume correlated with predicted whole brain age in the QTAB neurotypical controls (R^2^ = 0.02).

**Figure K14.**
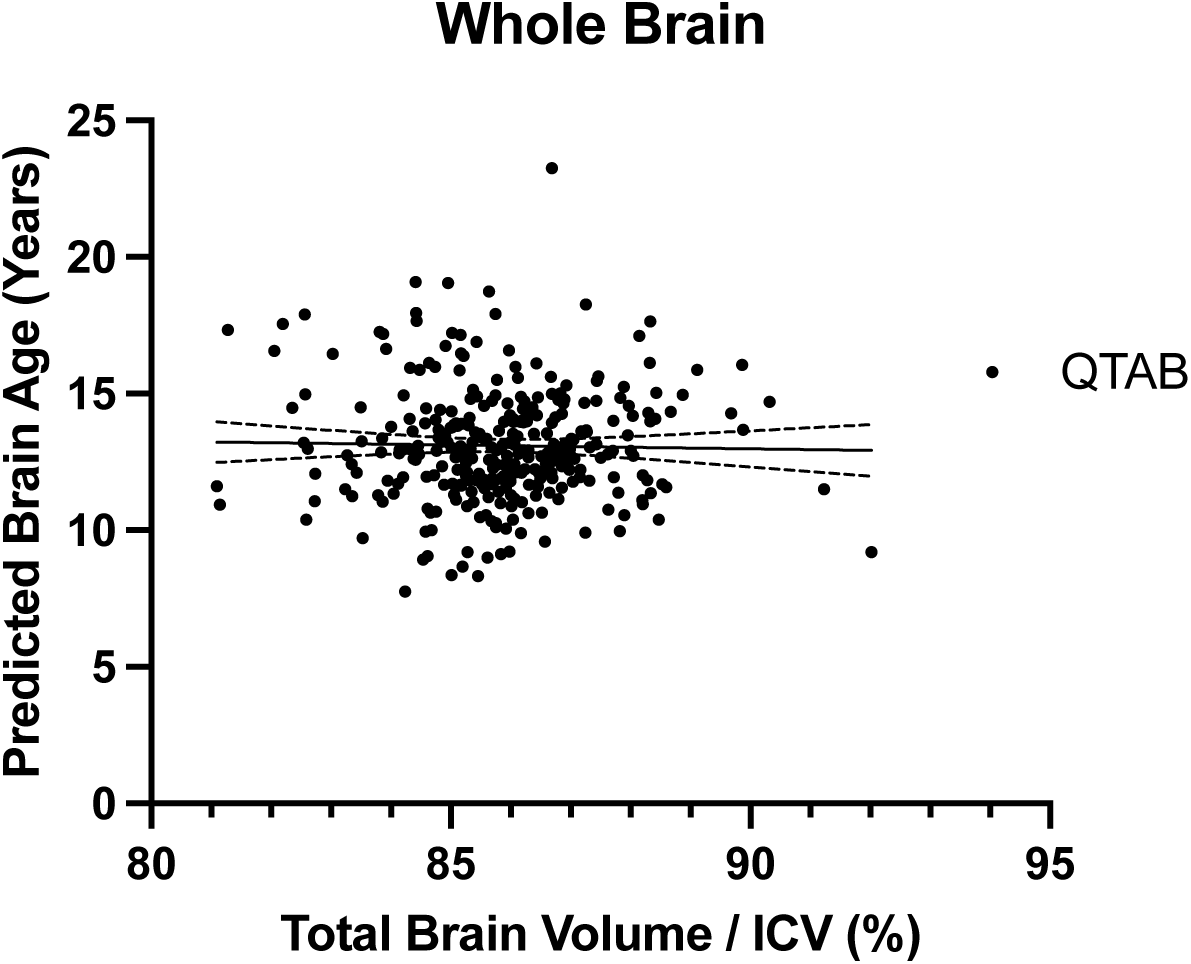
Correlation between whole brain volume controlled for total intracranial volume (ICV) and predicted whole brain age gap estimation in QTAB neurotypical controls. Total brain volume correlated with predicted whole brain age in the QTAB neurotypical controls (R^2^ < 0.01).

**Figure K15.**
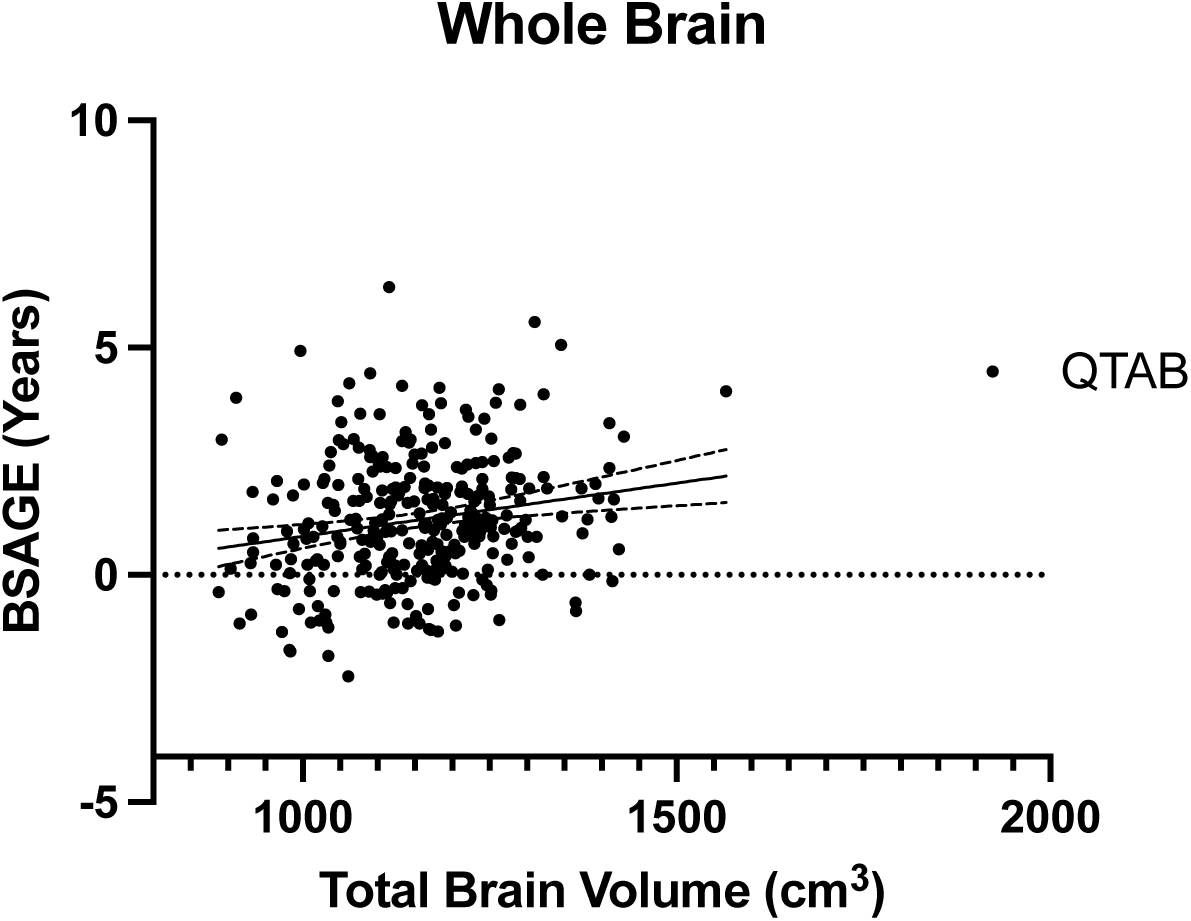
Correlation between whole brain volume (cm^3^) and predicted whole brain, brain structures age gap estimation (BSAGE) in the QTAB neurotypical controls. Total brain volume correlated with predicted whole brain age in the QTAB neurotypical controls (R^2^ = 0.04).

**Figure K16.**
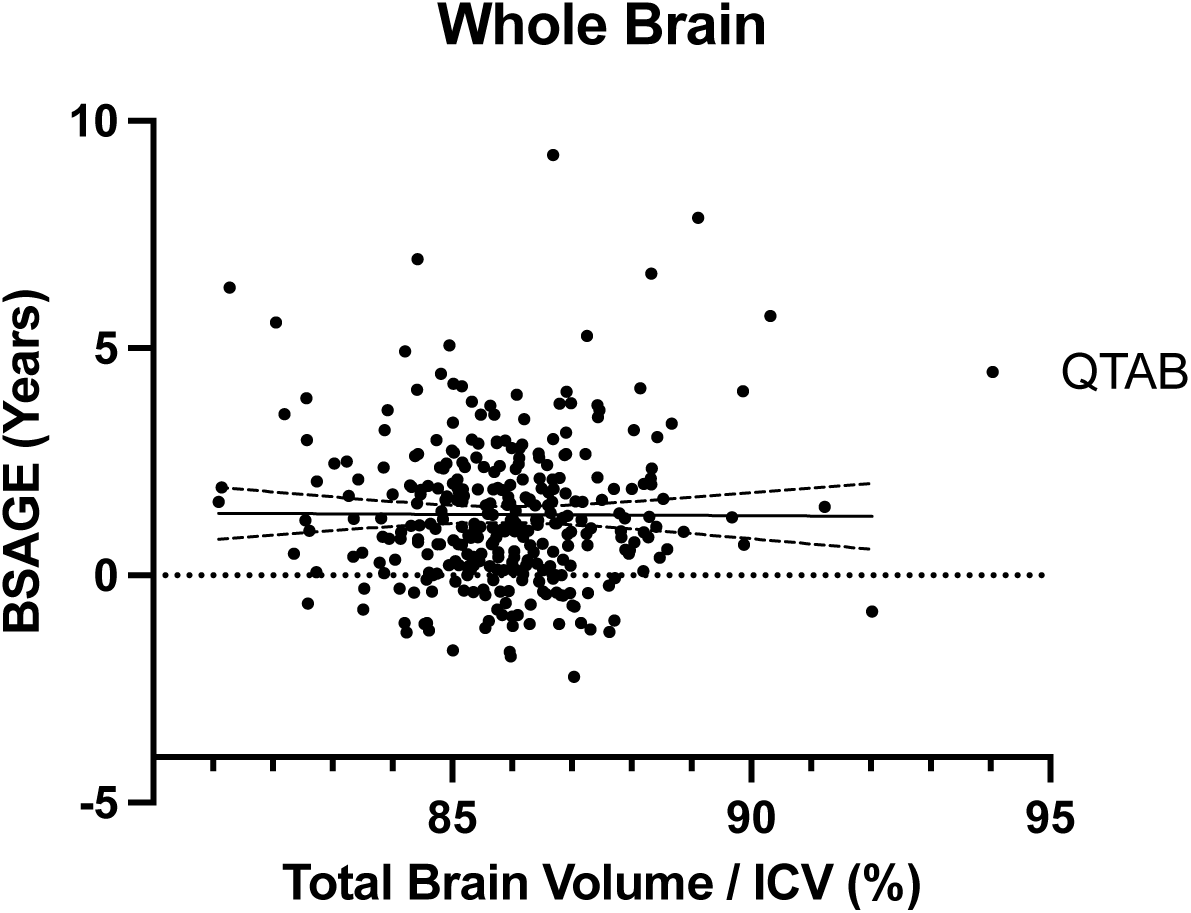
Correlation between whole brain volume controlled for total intracranial volume (ICV) and predicted whole brain, brain structures age gap estimation (BSAGE) in the QTAB neurotypical controls. Total brain volume did not correlate with predicted whole brain age in the QTAB neurotypical controls (R^2^ < 0.01).

**Figure K17.**
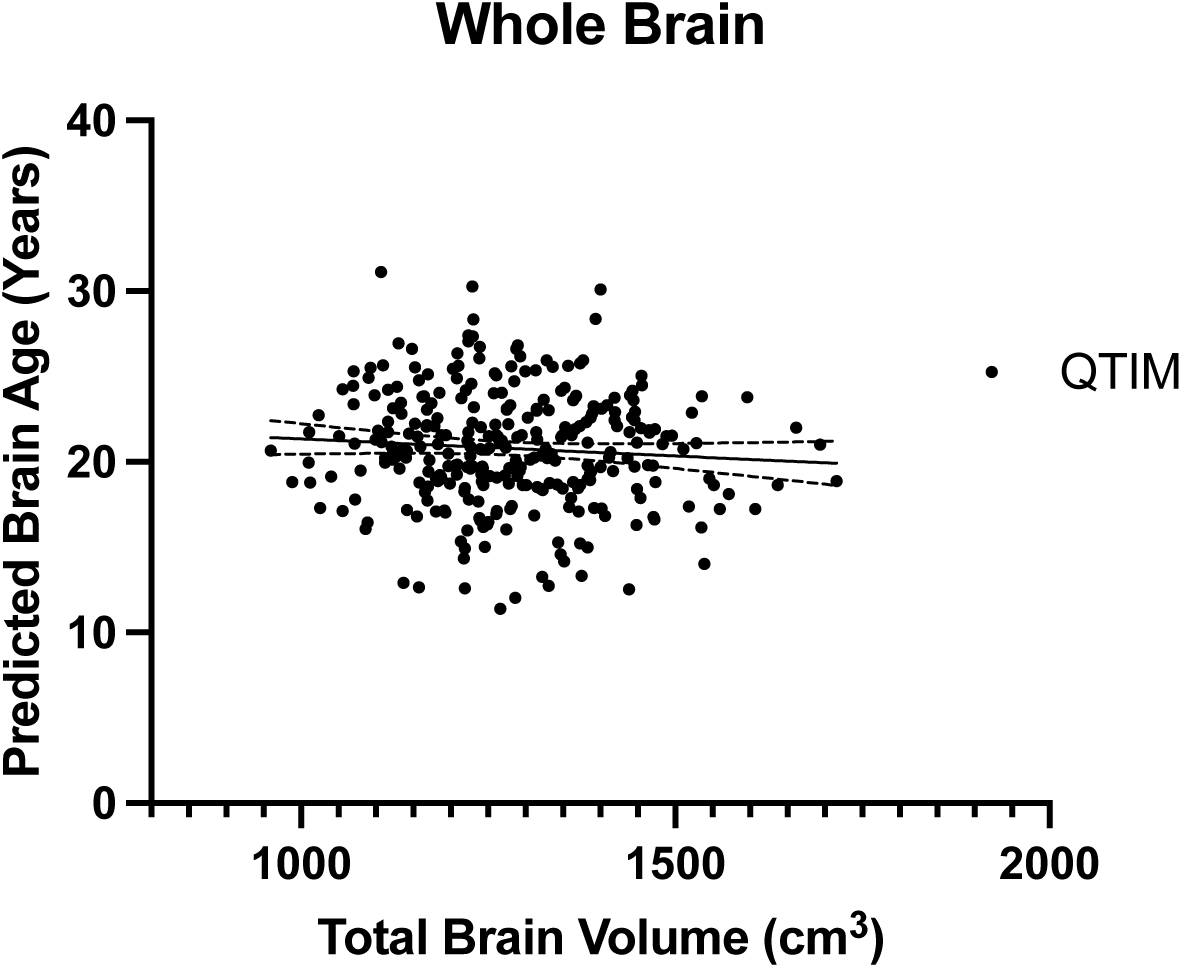
Correlation between whole brain volume and predicted whole brain age gap estimation in QTIM neurotypical controls. Total brain volume correlated with predicted whole brain age in the QTIM neurotypical controls (R^2^ = 0.01).

**Figure K18.**
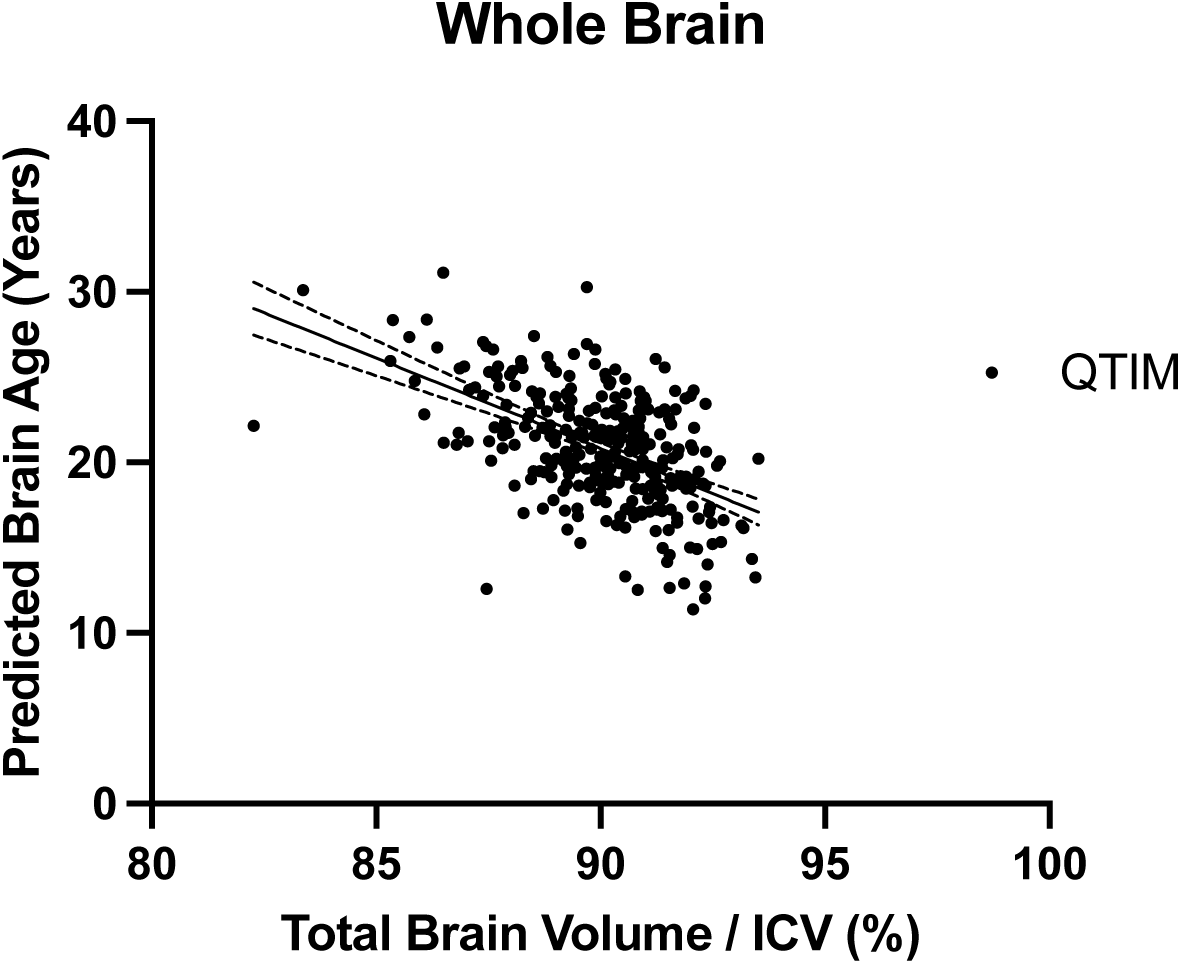
Correlation between whole brain volume controlled for ICV and predicted whole brain age gap estimation in QTIM neurotypical controls. Total brain volume correlated with predicted whole brain age in the QTIM neurotypical controls (R^2^ = 0.28).

**Figure K19.**
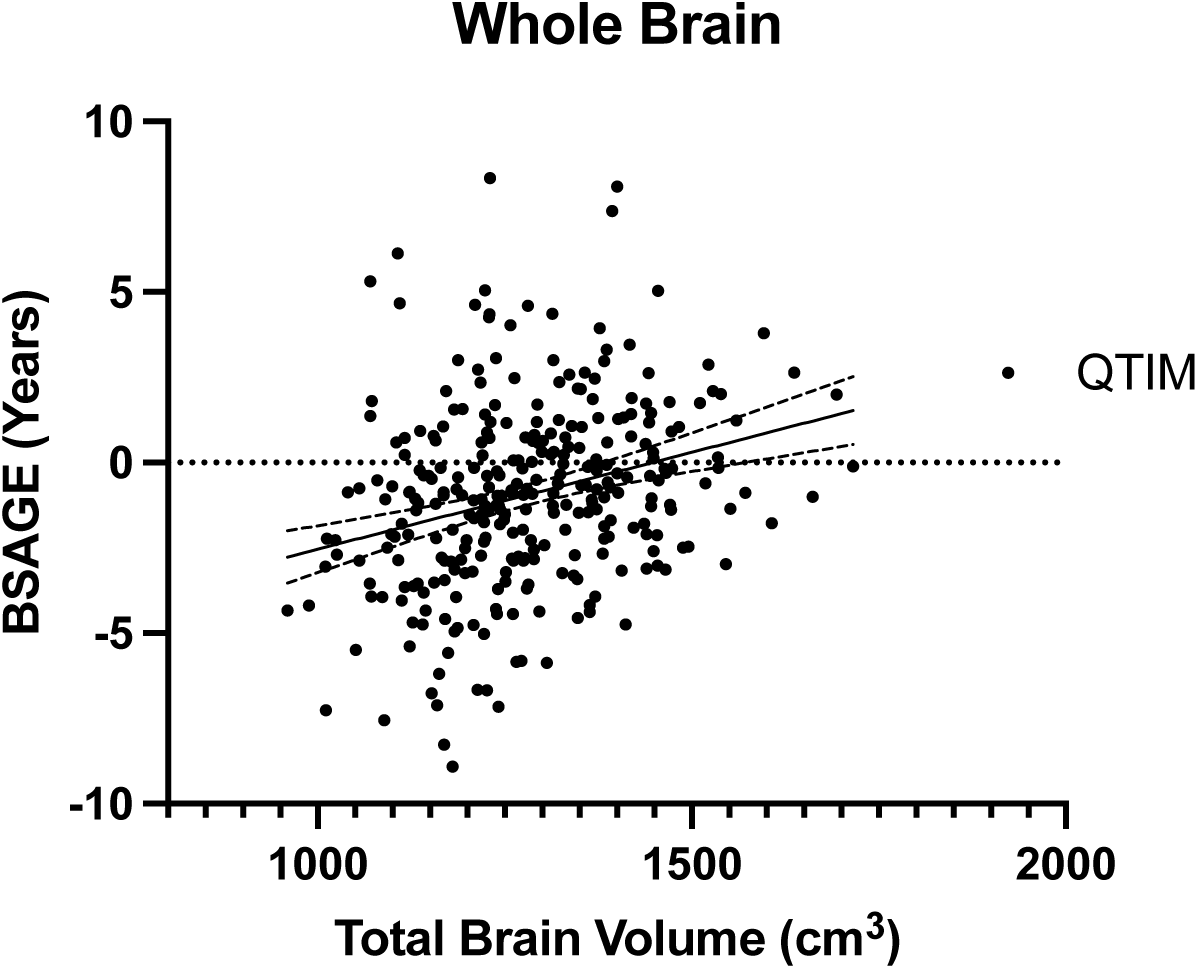
Correlation between whole brain volume (cm^3^) and predicted whole brain, brain structures age gap estimation (BSAGE) in the QTIM neurotypical controls. Total brain volume correlated with predicted whole brain age in the QTIM neurotypical controls (R^2^ = 0.08).

**Figure K20.**
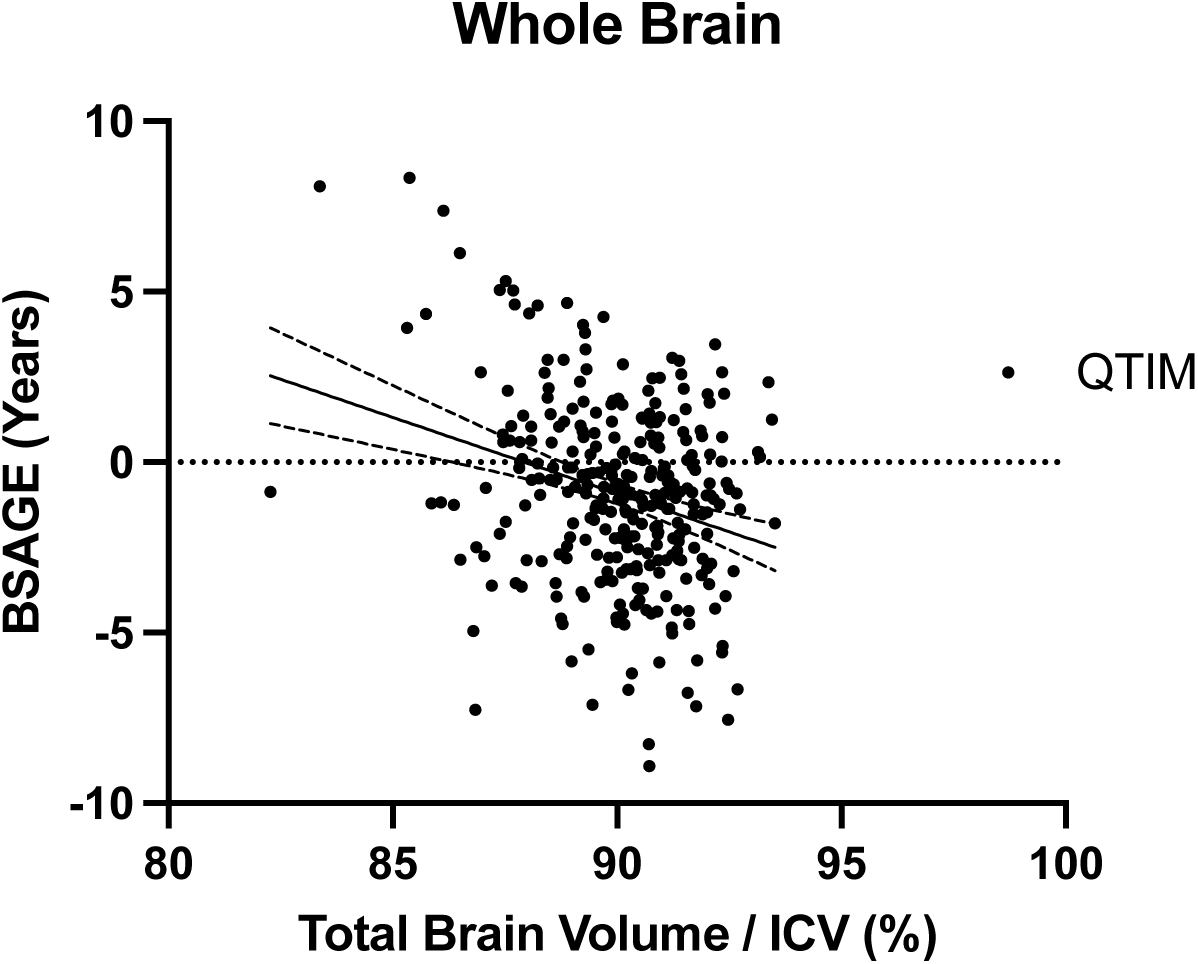
Correlation between whole brain volume controlled for total intracranial volume (ICV) and predicted whole brain, brain structures age gap estimation (BSAGE) in the QTIM neurotypical controls. Total brain volume correlated with predicted whole brain BSAGE in the QTIM neurotypical controls (R^2^ = 0.08).

